# Systematic Review and External Validation of Clinical Prediction Models for Adverse Pregnancy Outcomes Using Routinely Collected Pre-Conception and Early Pregnancy Data

**DOI:** 10.64898/2026.09.21.26363564

**Authors:** Yiran Zhang, Glen P Martin, Darren M Ashcroft, Tjeerd van Staa, Victoria Palin

**Author notes:** Corresponding Author: Yiran Zhang.

## Abstract

**Background:** Although numerous clinical prediction models (CPMs) have been developed to identify pregnancies at risk of serious adverse outcomes, most lack robust external validation, which restricts their reliability in clinical practice. This study aims to systematically review and externally validate existing clinical prediction models for Gestational Diabetes Mellitus (GDM), Pre-eclampsia (PE), Stillbirth, and Small-for-Gestational-Age (SGA), focusing on models for use in pre-conception or early pregnancy. The models were evaluated using a large, representative primary care cohort to assess their performance within a UK population.

**Methods:** Four two-stage systematic reviews identified CPMs for GDM, PE, Stillbirth, and SGA. Eligible models used routinely collected maternal characteristics available before conception or in early pregnancy and fully reported the model equation. Validation used the Clinical Practice Research Datalink (CPRD) Aurum, including over 1.58 million pregnancies among women aged 14-49 years between 2000 and 2019. Performance was assessed through discrimination (C-statistic), calibration-in-the-large (CITL), calibration slope and calibration plot, with pooled estimates across 20 imputations.

**Results:** Forty-eight models from 30 studies were included (47 binary and one continuous outcome), and 23 were developed in cohorts that included a UK population. Model development sample sizes ranged from 151 to 113,415 (median 5,013). Although 29 models had been externally validated, only one had been validated in a cohort including a UK population that met the required sample size and number of events. Across all outcomes, most models demonstrated limited generalisability in the UK population. C-statistics ranged from 0.57 to 0.77 for GDM, 0.50 to 0.72 for PE, 0.54 to 0.58 for stillbirth and 0.58 to 0.67 for SGA. Miscalibration was substantial across all outcomes, with CITL ranging from −6.14 to 9.64 (for binary-outcome models) and calibration slopes from 0.19 to 17.78.

**Conclusions:** Most existing CPMs that use pre-conception and early-pregnancy data to predict adverse pregnancy outcomes showed suboptimal discrimination and miscalibration in a large UK population, likely reflecting differences in population characteristics and outcome prevalence. Existing models are not suitable for direct implementation in UK practice and require further recalibration or updating using large, representative data before achieving clinical utility.

## Background

Pregnancy is a particularly vulnerable period for women and their babies, with approximately 15-25% of pregnancies reportedly ending in miscarriage [1]. In the UK, pregnancy complications such as pre-eclampsia (PE), gestational diabetes mellitus (GDM), and fetal growth restriction (FGR), affect approximately 5-8% of pregnancies, with a further 0.4% resulting in stillbirth [2–5]. Clinical prediction models (CPMs) have been increasingly developed for identifying pregnant women at higher risk of developing a complication or adverse outcome [6, 7]. For instance, the Fetal Medicine Foundation (FMF) developed a first trimester screening algorithm to identify pregnancies at high risk of preterm pre-eclampsia, which showed improved detection rates of PE compared to standard clinical screening and enhanced aspirin prescribing, improved risk stratification and saw a significant reduction in preterm and early pre-eclampsia rates [8, 9]. Although many CPMs to predict the risk of developing a complication or adverse pregnancy outcome exist, small sample size, short follow-up periods, as well as lack of robust external validation, diminish the performance in estimates, especially for prediction of rare outcomes [10–14]. Additionally, the majority of existing models were developed using data from selective populations in secondary or tertiary specialised care settings, which restricts their generalisability to routine antenatal populations. Many existing models have been developed for use during pregnancy, rather than at the pre-conception or early pregnancy stage, which limits their utility for early risk stratification and preventive intervention [7, 15].

External validation is essential before the implementation of a prediction model in clinical practice [16–18]. When the target-use population differs from the original development cohort, a reduction in model performance is common. Rigorous external validation therefore serves as a critical first step to determine if a model is ready for clinical implementation or if the model requires further refinement, such as re-calibration or subsequent updating, to ensure reliable performance in the intended population. To date, external validation studies of existing models have been restricted by small sample sizes, insufficient numbers of events, or non-representative cohorts, thereby limiting statistical performance and generalisability specifically for a national population [10–13, 19]. Furthermore, current CPMs for some outcomes have not been adequately validated in UK, non-specialised populations, limiting confidence in use for risk assessment within the National Health Service (NHS). Therefore, utilising large routinely collected and more accessible primary care data from early pregnancy is crucial for developing and validating timely and effective screening tools.

The UK offers a unique opportunity through its large-scale, routinely collected primary care datasets, containing electronic health records of clinical characteristics such as body mass index (BMI), blood pressure, smoking status, ethnicity, and comorbidity history, linked to secondary care data about the birth of babies. Furthermore, evidence from a recent systematic review demonstrated that pre-conception health data derived from these sources is strongly associated with adverse pregnancy outcomes [20]. This highlights that such data may be valuable for validating existing predictive models, which could then inform strategies to optimise pre-conception health, support early pregnancy risk stratification, and guide targeted interventions at a population level.

This study aimed to conduct a rigorous systematic review and external validation study of existing CPMs for the most common and serious adverse outcomes of high clinical and patient health importance in the UK: pre-eclampsia, gestational diabetes, fetal growth restriction, and stillbirth. During this validation, we focused on prediction models that use predictors available before conception and during early pregnancy, thus evaluating whether such routinely collected data in UK primary care records can be used for early prediction. Our focus was on maternal demographic and clinical history, rather than biomarkers, ultrasound findings, or screening results typically obtained later in pregnancy. To achieve this aim, we used the Clinical Practice Research Datalink (CPRD) Aurum, which is a large, nationally representative dataset of primary care records and linked secondary care data including birth outcomes.

## Methods

This study was conducted and reported in accordance with the Preferred Reporting Items for Systematic Reviews and Meta-Analyses (PRISMA) statement for the systematic review [21], and the Transparent Reporting of a multivariable prediction model for Individual Prognosis or Diagnosis + Artificial Intelligence (TRIPOD+AI) statement [22] for the external validation analysis. Validation results, including calibration, discrimination, and sensitivity analyses, are presented in line with TRIPOD recommendations to facilitate reproducibility, critical appraisal, and potential application of the findings in clinical settings. The overall study protocol was approved by the CPRD Independent Scientific Advisory Committee (protocol number: 22_002169, see Declarations), and the systematic review component was not registered separately.

### Literature search and selection

We conducted four outcome specific two-stage systematic reviews on PubMed to identify existing clinical prediction models (CPMs) for: gestational diabetes mellitus (GDM), stillbirth, pre-eclampsia (PE), and fetal growth restriction (FGR). Each review used the same broad approach but applied outcome-specific search terms and eligibility criteria. Eligible models were those developed to predict one of the specific pregnancy outcomes using routinely collected maternal characteristics available before conception or during early pregnancy. Routinely collected characteristics were defined as demographic factors, physical measurements, medical comorbidities, and maternal history. Existing literature used significant ambiguity in the definition and application of Small-for-Gestational-Age (SGA) and Fetal Growth Restriction (FGR), and often used interchangeably. Most definitions also failed to adhere to the international Delphi consensus definition, which incorporated expert agreement on fetal biometric, Doppler, and clinical predictors [23]. Consequently, we searched for all relevant publications that employed the terms FGR or SGA and used their original definition as outcome in subsequent model validation. Given that most model development studies utilised an SGA-based outcome (birthweight < 10^th^ centile), we henceforth use “SGA” to denote a growth restricted outcome in this study. Full details on the search strategies and screening criteria are illustrated in Additional file 1: Table S1.

Stage one identified existing systematic reviews, meta-analyses, or external validation studies for each outcome. Titles and abstracts were screened first, followed by full-text review of potentially eligible articles. Where reviews or validation studies reported potentially relevant prediction models, the original model development studies were retrieved and assessed to confirm eligibility, reproducibility, and applicability. In stage two, we conducted updated outcome-specific searches from the search end date of the most recent relevant review for each outcome to 1 June 2025. Titles and abstracts from the updated searches were screened before full texts were assessed for eligibility.

The original development studies were examined to ensure strict model reproducibility and applicability. Models were eligible for inclusion if they predicted a specific binary outcome, or a continuous outcome that could be dichotomised into the outcome of interest, e.g., as SGA is defined based on a fetal weight centile threshold, models predicting birthweight were also eligible for inclusion. In cases where a model was developed based on routinely collected maternal characteristics (prior-risk model), which was then updated with other biomarker or genetic data, the version of prior-risk model was included. Models were excluded if they used a survival or competing-risks framework, because the outcome was defined as time-to-event (which can be unreliable in routine maternity data where care is delivered across care sites) rather than the occurrence of the complication itself. In addition, models including fewer than three predictors were excluded to distinguish comprehensive risk algorithms from simple association or biomarker-screening tests. Models were also excluded if their individual predictors could not be replicated within our validation database or required predictors that are not routinely collected data such as biomarkers, ultrasound measures, or genetic information. Furthermore, models were excluded if they did not report sufficient details for full reproduction of the prediction equation, such as the intercept and/or predictor coefficients for each predictor in a logistic regression model. Models that reported all predictor coefficients but omitted the intercept were excluded from the primary analysis (as calibration cannot be evaluated without absolute risk predictions) and were instead retained for a sensitivity analysis evaluating discrimination alone.

### Data Source and Study cohort

This study utilised the Clinical Practice Research Datalink Aurum (CPRD Aurum, June 2021 release) and its linked datasets, including the Pregnancy Register and Hospital Episode Statistics Admitted Patient Care (HES-APC) [24]. CPRD Aurum is a representative database of anonymised patient electronic health records from primary care settings in the UK, which includes comprehensive longitudinal records, covering 1,373 contributing primary care practices and approximately 20% of the population in the UK [25]. The study cohort included women aged 14 to 49 who were pregnant between 1 January 2000 and 31 December 2019. Follow-up for each woman began at the latest of the study start, or the patient registration date. Follow-up ended at the earliest of study end date, or date of deregistration (where available), or date of death (if applicable). Only singleton pregnancies occurring entirely within the follow-up period were eligible for inclusion. To ensure clinical plausibility and outcome relevance, pregnancies were included in the validation cohort only if their gestational length was 20-42 weeks. Separate validation datasets were constructed for each outcome due to the variation in definitions among outcomes and predictors. Additional file 2: Figure S1 shows the process of constructing the cohort for primary analysis.

### Definition of outcomes and predictors

Outcomes for GDM, PE, and stillbirth, as well as predictors were defined using medical codes from CPRD Aurum and ICD-10 codes from HES-APC. Relevant medical codes were identified using keyword searches in the CPRD Code Browser and reviewed for comprehensiveness. For HES-APC, ICD-10 codes were selected from the relevant diagnostic categories of the ICD-10 dictionary, retaining only the first five characters to align with HES-APC coding conventions and ensure consistency. In addition, published code lists, including SNOMED CT and Read codes, were reviewed and mapped to relevant formats where appropriate, before our final review. For outcomes of SGA, birthweight data were extracted from HES-APC and then were standardised into Z-scores using the World Health Organization (WHO) calculation algorithm [26, 27]. These Z-scores were subsequently mapped to percentiles based on the standard normal distribution. The binary outcome was then defined by applying the specific percentile threshold as defined in each original development study.

To account for potential delays in the recording of events across different care settings, outcome time windows were uniformly extended by two weeks. For example, stillbirth, originally defined as occurring from 24 weeks in gestation until the end of pregnancy, was redefined to include any record from 24 weeks in gestation up to two weeks after the documented pregnancy end date. In terms of predictors, due to the inherent limitations of retrospective data, information on maternal characteristics and health conditions needed to be retrieved from historical patients records. This included physical measures (e.g., height, weight, and blood pressure), medical history, and chronic conditions (e.g., chronic hypertension, diabetes, autoimmune diseases). Therefore, the observation window for these predictors was extended up to five years before pregnancy start for long-term conditions, or up to one year prior for more recent clinical indicators, depending on the nature of the predictor. If a patient had multiple medical records during the time window, the latest was used. To ensure clinical plausibility and minimise bias from data entry errors, values were treated as missing if they fell outside the following ranges: maternal weight (40 - 200 kg), height (100 - 200 cm), BMI (10 - 100 kg/m²), systolic blood pressure (70 - 250 mmHg), diastolic blood pressure (40 - 150 mmHg). See Availability of Data and Materials section for code lists and definitions of outcomes and predictors.

### Required Sample Size Calculation

Sample size requirements were calculated following the recommendations of Riley et al. [28]. Specifically, sample sizes were determined to achieve prespecified precision across three criteria: the observed-to-expected (O/E) ratio (target 95% confidence interval [CI] width = 0.2), calibration slope (target 95% CI width = 0.2), and C-statistic (target 95% CI width = 0.1), taking the largest requirement across evaluable criteria [28]. For each eligible model, we used the outcome prevalence in our validation cohort and the reported C-statistic from the development study, assuming normally distributed linear predictors with common variance in event and non-event groups (Additional file 1: Table S2). Using the same approach, we also calculated the required sample size for each existing external validation study based on its own cohort outcome prevalence (evaluating the largest cohort in multi-cohort studies) to assess whether its sample size was sufficient.

### Missing data and multiple imputation

To address missingness in predictors (height, weight, systolic and diastolic blood pressure) we performed multiple imputation by chained equations (MICE), generating 20 imputed datasets, separately for each outcome of interest [29]. Missing body mass index (BMI) and mean arterial pressure (MAP) were derived from imputed weight, height, and systolic/diastolic blood pressure values. Missing ethnicity was retained as an “Unknown” category. For the imputation process of each outcome-specific validation dataset, all predictors from the models associated with that outcome, along with their outcomes, were included. As all predictors subject to imputation were continuous, linear regression imputation with added normally distributed residuals was employed. For each imputed predictor, values were constrained within clinically and statistically plausible ranges as observed in the original data, and convergence was assessed using diagnostic plots to ensure stability and consistency of the imputed distributions.

### Validation of existing models

The models selected for validation included linear regression, logistic regression, and Poisson regression models. To validate each model, the linear predictor (LP, i.e., the sum of each covariate multiplied by the corresponding model coefficient as reported in the original development study of each CPM) was calculated for every pregnancy episode using the published model equation. For logistic models, the predicted risk probability of the outcome was then obtained by applying the inverse logit transformation [30]. The predicted probability of Poisson models was calculated by applying an exponential transformation to the linear predictor [31].

Calibration was assessed using three measures: calibration slope, calibration-in-the-large (CITL), and calibration plots. The calibration slope assesses the overall calibration by fitting a logistic regression model (for binary outcome) or linear regression model (for continuous outcome) of the outcomes on the linear predictor, where a slope of one indicates perfect calibration. CITL evaluated whether the mean predicted risk across all individuals matched the overall observed event rate in the population, with an ideal value of zero indicating perfect agreement on average. A calibration plot visually assessed model calibration by plotting observed event frequencies against predicted risk probabilities. Perfect calibration occurred when the plotted curve aligns with the 45-degree diagonal line. Discrimination was assessed using the C-statistic (area under the receiver operating characteristic curve, AUC), where a value of one denotes perfect discrimination [16, 30].

All performance metrics (calibration slope, CITL, and C-statistic) were estimated for each imputation cohort and pooled across the 20 datasets using Rubin’s rules along with corresponding 95% confidence intervals (CIs). For calibration plots, a single pooled plot was generated for each model by first averaging the LP across all imputed datasets and then transforming the pooled LP into predicted probabilities.

### Sensitivity Analysis

As CPRD pregnancy episodes are algorithm derived, some start and end dates of pregnancy episodes for individuals may overlap. To mitigate potential bias, a validation sub-cohort retaining only the first chronological pregnancy for overlapping episodes was used to evaluate performances using the same discrimination and calibration metrics. A complete-case sensitivity analysis was also conducted to assess the robustness of the external validation regarding the approach to missing data. For this, the validation cohort was restricted to pregnancy episodes with complete records for both outcomes and predictors to re-estimate discrimination and calibration metrics. In addition, for models reporting complete predictors and coefficients but lacking the intercept, we further assessed discrimination (C-statistic) alone using the linear predictor excluding the intercept. By applying the same validation cohorts, 20 imputed datasets, and Rubin’s rules as in the primary analysis, pooled C-statistics and 95% CIs were estimated for each model.

### Fairness

Assessment of fairness was not a primary objective of this study. The primary aim was to externally validate previously developed prediction models. The original model development studies were conducted in populations with substantially different demographic and ethnic compositions. Therefore, fairness-related analyses (e.g., evaluation of model performance across ethnic subgroups) were not applicable within the scope of this work. Patients and the public were not involved in the design, conduct, reporting, or dissemination plans of this research.

## Results

### Eligible Models for External Validation

Stage one of the four two-stage systematic reviews identified a total of 664 articles from systematic reviews, meta-analyses, or external validation studies across the four specified outcomes, following full text screening of 88 eligible studies and subsequent review of original development studies, 43 clinical prediction models (CPMs) were included meeting the criteria for external validation. Within this first stage, the literature search dates for the most recent systematic reviews concluded on 06 June 2024 for GDM, PE, and SGA [7], and on 18 October 2022 for stillbirth [32]. The second stage, which focused on models published after the most recent systematic review for each outcome yielded an additional 1,269 articles. From these, 172 articles were eligible for full text screening, resulting in five additional models from three articles meeting the inclusion criteria (Figure 1). In total, 48 eligible models from 30 original development studies were included (47 binary-outcome models and one continuous-outcome model). For a comprehensive list of all models that were reviewed in full text but subsequently excluded, along with exclusion reasons, see Additional file 3: Table S11.

**Figure 1.**
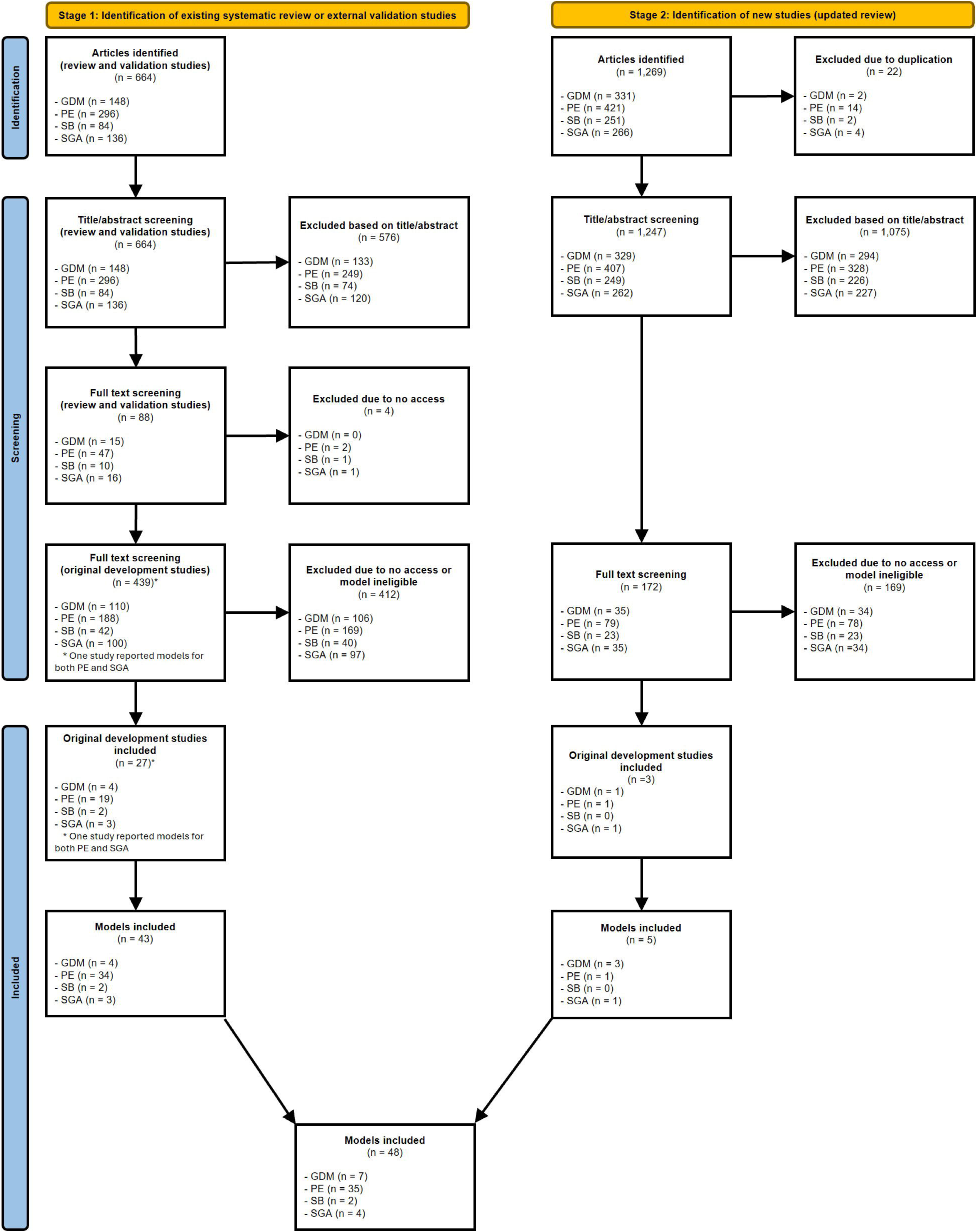
Flow diagram of the two-stage systematic review and model selection process. Stage 1 identifies models from existing systematic reviews and external validation studies, while Stage 2 involves an updated search for newly published models. Abbreviations: GDM, gestational diabetes mellitus; PE, pre-eclampsia; SB, stillbirth; SGA, small-for-gestational-age.

Table 1 summarises the characteristics of the 48 included models by outcome. Outcome definitions varied across models. For stillbirth, one model defined the outcome as occurring at ≥24 weeks of gestation, whereas the other used ≥32 weeks. Among the 35 PE models, 12 predicted any PE regardless of onset, eight each predicted early-onset PE (EOPE, <34 gestational weeks) and late-onset PE (LOPE, ≥34 gestational weeks), four predicted preterm PE (<37 gestational weeks), one predicted term PE (≥37 gestational weeks), one predicted intermediate PE (34-37 gestational weeks), and one predicted late PE (≥38 gestational weeks). For SGA, three models predicted binary SGA and one linear regression model predicted log-transformed birthweight. Across all 48 models, 23 were developed using cohorts that included a UK population. The number of predictors per model ranged from 3 to 17 (median 6). Substantial variation was observed in original development sample sizes, ranging from 151 to 113,415 (median 5,013), as well as in outcome prevalence across the development cohorts (GDM: 2.41%-55.35%; Stillbirth: 0.35%-0.58%; Any PE: 1.78%-66.89%; EOPE: 0.50%-11.97%; LOPE: 1.53%-16.53%; Preterm PE: 1.65%-20.00%; Term PE: 4.64%; Intermediate PE: 0.76%; Late PE: 2.22%; SGA: 9.50%-19.53%). Details of the model developed with the largest sample size for each outcome are presented in Table 2, with full specifications of all 48 models provided in Additional file 1: Table S2.

**Table 1.** Overview of the selected models for each outcome.

| **Outcome** |  | **Models, N** | **Models developed with a UK population, N<sup>#</sup>** | **Predictor count (median), N** | **Sample size (median), N** | **Reported C-statistics in original development study** |
|---|---|---|---|---|---|---|
| **Gestational diabetes (GDM)** | GDM | 7 | 3 | 4 - 8 (7) | 253 - 20,549 (1,205) | 0.72 - 0.88 |
| **Stillbirth** | Stillbirth (≥ 24 weeks) | 1 | 1 | 8 (8) | 113,415 (113,415) | 0.64 |
|  | Stillbirth (≥32 weeks) | 1 | 0 | 12 (12) | 57,326 (57,326) | 0.66 |
| **Pre-eclampsia (PE)** | Any PE | 12 | 6 | 3 - 12 (6) | 151 - 29,187 (3,840) | 0.67 - 0.85 |
|  | Early-onset PE | 8 | 3 | 3 - 8 (5) | 568 - 29,187 (5,593) | 0.72 - 0.83 |
|  | Late-onset PE | 8 | 4 | 3 - 9 (7) | 599 - 29,187 (7,191) | 0.68 - 0.83 |
|  | Preterm PE | 4 | 1 | 3 - 5 (3) | 235 - 1,756 (733) | 0.76 - 0.84 |
|  | Term PE | 1 | 0 | 3 (3) | 733 (733) | 0.75 |
|  | Intermediate PE | 1 | 1 | 6 (6) | 4,855 (4,855) | 0.77 |
|  | Late PE | 1 | 1 | 5 (5) | 4,855 (4,855) | 0.76 |
| **Small-for-gestational-age (SGA)** | SGA | 3 | 2 | 3 - 13 (10) | 988 - 107,875 (12,679) | 0.62 - 0.72 |
|  | Birthweight | 1 | 1 | 17 (17) | 33,602 (33,602) | Not applicable |

**Note:**
\# Models developed in multinational cohorts that included a UK population were counted.

**Table 2.** Validation results for the models with largest sample sizes for each outcome.

| **Outcome** | **Model** | **Development sample size (cases), N** | **Data type** | **Validation result in primary analysis** |  |  |
|---|---|---|---|---|---|---|
|  |  |  |  | **C-statistic** | **Calibration-in-the-large** | **Calibration slope** |
| **Gestational diabetes** | Syngelaki 2025 (for nulliparous) [34] | 20,549 (1,787) | Prospective | 0.75 (0.75 - 0.76) | -0.78 (-0.79 - -0.76) | 1.07 (1.05 - 1.08) |
| **Any pre-eclampsia** | Allotey 2020 (a) [11] | 29,187 (1,187) | Retrospective | 0.72 (0.70 - 0.74) | -0.19 (-0.20 - -0.18) | 1.10 (1.01 - 1.20) |
| **Early-onset pre-eclampsia** | Allotey 2020 (a) [11] | 29,187 (149) | Retrospective | 0.67 (0.66 - 0.68) | -0.74 (-0.77 - -0.70) | 0.82 (0.79 - 0.86) |
| **Late-onset pre-eclampsia** | Allotey 2020 (a) [11] | 29,187 (1,039) | Retrospective | 0.71 (0.69 - 0.73) | -0.09 (-0.10 - -0.08) | 1.04 (0.94 - 1.14) |
| **Preterm PE** | Sepúlveda-Martínez 2019 [41] | 1,756 (29) | Prospective | 0.62 (0.61 - 0.62) | -0.71 (-0.74 - -0.69) | 0.81 (0.78 - 0.83) |
| **Term PE** | Rocha 2017 [42] | 733 (34) | Prospective | 0.66 (0.59 - 0.73) | -4.10 (-4.12 - -4.09) | 10.58 (6.42 - 14.73) |
| **Intermediate PE** | Lai 2013 [43] | 4,855 (37) | Prospective | 0.68 (0.65 - 0.70) | -0.01 (-0.03 - 0.02) | 0.56 (0.49 - 0.63) |
| **Late PE** | Lai 2013 [43] | 4,855 (108) | Prospective | 0.68 (0.68 - 0.68) | -0.03 (-0.05 - -0.02) | 0.70 (0.69 - 0.72) |
| **Stillbirth (≥ 24 weeks)** | Yerlikaya 2016 [33] | 113,415 (396) | Prospective | 0.58 (0.57 - 0.58) | 0.49 (0.46 - 0.51) | 0.87 (0.83 - 0.91) |
| **Stillbirth (≥ 32 weeks)** | Trudell 2017 [44] | 57,326 (330) | Retrospective | 0.54 (0.53 - 0.55) | 0.89 (0.86 - 0.92) | 0.42 (0.36 - 0.48) |
| **Small-for-gestational-age (SGA)** | Adjahou 2025 [45] | 107,875 (12,420) | Retrospective | 0.67 (0.67 - 0.67) | -0.16 (-0.16 - -0.15) | 0.73 (0.72 - 0.73) |
| **Birthweight** | Poon 2011 (log(birthweight)) [35] | 33,602 | Prospective | Not applicable | 104.23 g (103.46 - 105.01) | 0.94 (0.94 - 0.95) |

**Note:**
See Additional file 1: Table S2 for information on all models and Additional file 1: Table S5 for validation results of all models.

Regarding previous external validations of the 48 included models, 29 models (28 binary-outcome models and one continuous-outcome model) had been externally validated across 13 published studies, including 21 models validated in cohorts containing a UK population (Additional file 1: Table S3). Cohort sample sizes across these validation studies ranged from 230 to 406,286. All existing validation studies of binary-outcome models reported discrimination (C-statistics), whereas three model validations (across three studies) did not report calibration metrics.

### Sample Size Calculation

For our external validation in the CPRD cohort, the minimum required sample size could be calculated for 42 of the 48 included models (excluding five PE models that did not report a C-statistic in their original development studies and one continuous birthweight model). Based on the outcome prevalence in our validation cohort, the estimated minimum sample size required for our external validation ranged from 5,905 pregnancies (795 events) for SGA to 307,424 pregnancies (1,138 events) for stillbirth ≥32 weeks (with required events ranging from 369 to 2,711 across all evaluable models; Additional file 1: Table S2). All required sample sizes and event counts were substantially smaller than our available CPRD validation cohorts. When evaluated against the sample size requirements calculated for each existing validation cohort, only one model (Yerlikaya 2016 for stillbirth ≥24 weeks [**33**]) had previously been validated in a cohort including a UK population that satisfied both the required sample size and number of events (Additional file 1: Table S3).

### Study Cohort Characteristics

In the primary analysis, the validation dataset for GDM, PE, and stillbirth included 1,063,974 women with 1,581,161 unique pregnancy episodes. Routine maternal characteristics such as maternal age, parity, ethnicity, and body mass index (BMI) were available, although some predictors had notable levels of missingness (weight, height, systolic and diastolic blood pressure, see Table 3 and Additional file 1: Table S4), consistent with the nature of routinely collected clinical data. The mean maternal age was 29.86 years, and 43.96% of pregnancies were nulliparous. In terms of ethnicity, 79.55% of the cohort were White, followed by Asian (10.21%) and Black (4.85%) women. Filtering to observations with a recorded birthweight to validate models predicting SGA reduced the validation cohort to 957,496 pregnancies, resulting in minor changes in the prevalence of predictors (Table 3). The prevalence of the outcomes of interest, including GDM, SGA, PE, and stillbirth, was consistent with national statistics [2–4], ensuring the representativeness and external validity of the study cohort. The main characteristics of the cohort for primary analysis are described in Table 3, and that of each predictor used in validating each model is detailed in Additional file 1: Table S4.

**Table 3.** Characteristics of the pregnancy cohort for primary variables and outcomes.

|  |  | **Full cohort<br>(Pregnancies, N = 1,581,161)** |  | **Sub-cohort for small-for-gestational-age<br>(Pregnancies, N = 957,496)** |  |
|---|---|---|---|---|---|
| **Variable** |  | Mean (SD) / Count (%)<sup>#</sup> | Missing, N (%)<sup>#</sup> | Mean (SD) / Count (%)<sup>#</sup> | Missing, N (%)<sup>#</sup> |
| **Maternal age (years)** |  | 29.86 (5.68) | 0 | 29.77 (5.63) | 0 |
| **Gravidity (total pregnancies)** |  | 2.42 (1.60) | 0 | 2.37 (1.54) | 0 |
| **Parity** | Nulliparous | 695,066 (43.96%) | 0 | 428,502 (44.75%) | 0 |
|  | Parous | 886,095 (56.04%) | 0 | 528,994 (55.25%) | 0 |
| **Gestational length (weeks)** |  | 39.03 (2.67) | 0 | 39.19 (1.89) | 0 |
| **Height (cm)** |  | 163.94 (6.94) | 505,291 (31.96%) | 163.90 (6.91) | 287,167 (29.99%) |
| **Weight (kg)** |  | 68.81 (16.04) | 326,710 (20.66%) | 68.95 (16.10) | 177,802 (18.57%) |
| **Body Mass Index (kg/m<sup>2</sup>)** |  | 25.55 (5.74) | 442,627 (27.99%) | 25.62 (5.76) | 243,526 (25.43%) |
| **Ethnicity** | **Bangladeshi** | 21,848 (1.38%) | 23,039 (1.46%) | 15,198 (1.59%) | 10,831 (1.13%) |
|  | **Black African** | 48,205 (3.05%) |  | 29,668 (3.10%) |  |
|  | **Black Caribbean** | 15,414 (0.97%) |  | 9,241 (0.97%) |  |
|  | **Black Other** | 13,071 (0.83%) |  | 8,103 (0.85%) |  |
|  | **Chinese** | 9,399 (0.59%) |  | 5,978 (0.62%) |  |
|  | **Indian** | 46,372 (2.93%) |  | 28,660 (2.99%) |  |
|  | **Mixed** | 22,507 (1.42%) |  | 14,184 (1.48%) |  |
|  | **Other** | 39,763 (2.51%) |  | 24,755 (2.59%) |  |
|  | **Other Asian** | 31,908 (2.02%) |  | 20,043 (2.09%) |  |
|  | **Pakistani** | 51,834 (3.28%) |  | 33,768 (3.53%) |  |
|  | **White** | 1,257,801 (79.55%) |  | 757,067 (79.07%) |  |
| **Gestational diabetes** |  | 64,522 (4.08%) | 0 | 44,623 (4.66%) | 0 |
| **Pre-eclampsia (Any)** |  | 34,856 (2.20%) | 0 | 23,304 (2.43%) | 0 |
| **Pre-eclampsia (early-onset, < 34 weeks)** |  | 3,281 (0.21%) | 0 | 2,415 (0.25%) | 0 |
| **Pre-eclampsia (late-onset, ≥ 34 weeks)** |  | 31,575 (2.00%) | 0 | 20,889 (2.18%) | 0 |
| **Pre-eclampsia (preterm, < 37 weeks)** |  | 8,106 (0.51%) | 0 | 6,237 (0.65%) | 0 |
| **Pre-eclampsia (term, ≥ 37 weeks)** |  | 26,750 (1.69%) | 0 | 17,067 (1.78%) | 0 |
| **Pre-eclampsia (intermediate, 34 - 37 weeks)** |  | 8,479 (0.54%) | 0 | 6,968 (0.73%) | 0 |
| **Pre-eclampsia (late, ≥ 38 weeks)** |  | 23,096 (1.46%) | 0 | 13,921 (1.45%) | 0 |
| **Stillbirth (≥24 weeks)** |  | 7,748 (0.49%) | 0 | 3,739 (0.39%) | 0 |
| **Stillbirth (≥32 weeks)** |  | 5,922 (0.37%) | 0 | 2,482 (0.26%) | 0 |
| **Birthweight (g)** |  | 3,377.44 (542.07) | 623,665 (39.44%) | 3,377.44 (542.07) | 0 |
| **Small-for-gestational-age** |  | / | / | 128,748 (13.45%) | 0 |

**Note:**
**SD:** standard deviation
\# Percentages may not sum to 100% due to rounding.

### External Validation of Models

The performance metrics of all models displayed significant heterogeneity, with a summary of distribution for each outcome shown in Figure 2. Detailed performance metrics and calibration plots for individual models are presented in Additional file 1: Table S5 and Additional file 2: Figure S2. The validation results for the model possessing the largest development dataset within each outcome are presented in Table 2.

**Figure 2.**
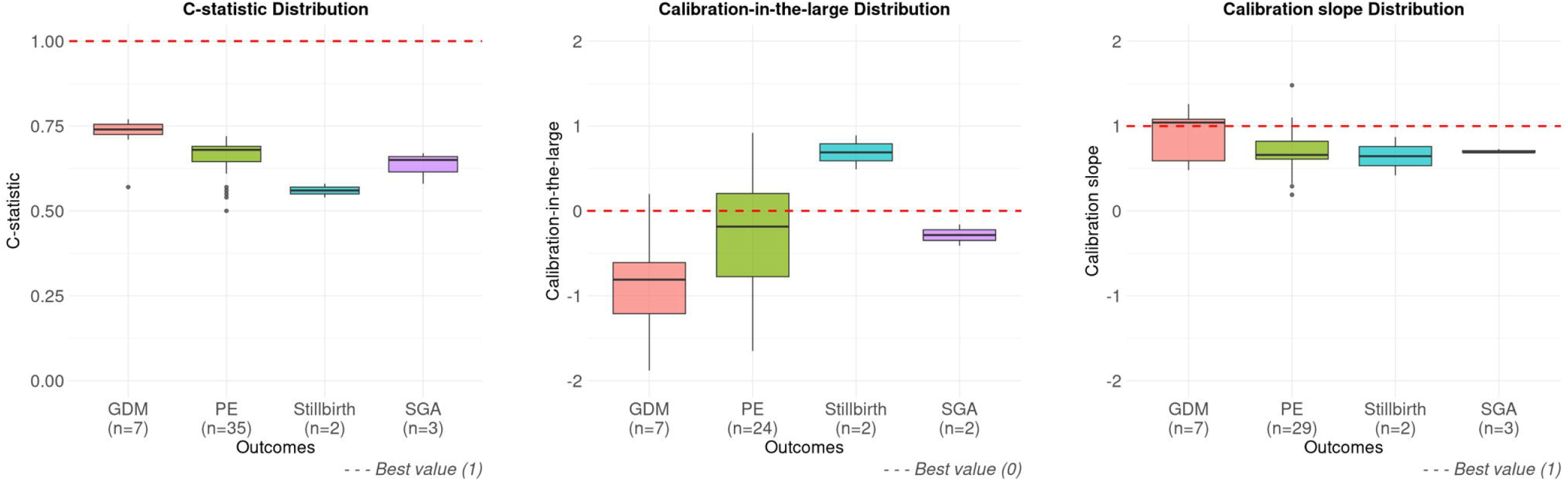
Distribution of external validation performance by outcome in the primary analysis. Boxplots display the C-statistic, calibration-in-the-large, and calibration slope. The red dashed lines indicate the ideal performance values (1.00 for C-statistic and calibration slope; 0 for calibration-in-the-large). Abbreviations: GDM, gestational diabetes mellitus; PE, pre-eclampsia; SGA, small-for-gestational-age. Note: The continuous linear regression model was excluded from all panels because C-statistic is not applicable and its calibration-in-the-large is measured on a continuous scale (grams) rather than log-odds. To preserve visual interpretability and prevent axis distortion caused by extreme outliers, binary-outcome models with calibration-in-the-large or calibration slope values outside the range of −2 to 2 were excluded from the respective calibration panels.

Compared with models for the other outcomes, models for gestational diabetes (GDM) demonstrated higher discriminative ability and less extreme calibration instability. Across all GDM models, C-statistics ranged from 0.57 to 0.77, calibration-in-the-large (CITL) from −1.88 to 0.20, and calibration slopes from 0.48 to 1.26. Six of the seven models achieved C-statistics above 0.70 (0.71 - 0.77), with only one model [34] tailored to high-risk parous women exhibiting a lower C-statistic (0.57). Most models yielded negative CITL (−1.88 to −0.44), indicating a consistent tendency to systematically overestimate baseline risk in the UK population. Regarding calibration slopes, four models showed slight underfitting (1.04 to 1.26), whereas the remaining three models suffered from overfitting with slopes (0.48 to 0.69).

Across all 35 pre-eclampsia (PE) models, C-statistics ranged from 0.50 to 0.72, CITL from −6.14 to 0.92, and calibration slopes from 0.19 to 17.78. Discrimination varied considerably across outcome definitions, ranging from 0.55 to 0.72 for any PE, 0.50 to 0.69 for early-onset PE (EOPE), 0.54 to 0.71 for late-onset PE (LOPE), and 0.62 to 0.68 for preterm, term, intermediate, and late PE. Most models (29 of 35) yielded negative CITL (−6.14 to −0.01), indicating a consistent tendency to overestimate baseline risk, including five models with severe overestimation beyond −5.00 (−6.14 to −5.14) and 24 models ranging from −4.10 to −0.01. Regarding calibration slopes, the majority of models (26 of 35) suffered from overfitting with slopes below 1.0 (0.19 to 0.92), whereas four models showed calibration slopes from 1.04 to 2.94, and five models showing extreme underfitting with slopes above 5.00 (5.54 to 17.78).

The two stillbirth models showed relatively poor discrimination in this validation cohort, with C-statistics of 0.58 and 0.54. In terms of calibration, both models yielded positive CITL (0.49 and 0.89), indicating systematic underestimation of risk. Regarding calibration slopes, both models suffered from overfitting with slopes below 1.0 (0.42 and 0.87). Calibration plots showed little separation of risks and no clinically useful stratification (Additional file 2: Figure S2).

Across the three binary outcome models for SGA, C-statistics ranged from 0.58 to 0.67. CITL varied widely from −0.41 to 9.64, reflecting slight baseline risk overestimation in two models (−0.41 and −0.16) but severe underestimation in one model (9.64). Regarding calibration slopes, all three binary models suffered from overfitting with slopes below 1.0 (0.69 to 0.73). In addition, the continuous linear regression model [35] predicting birthweight had a calibration slope close to ideal (0.94) and a CITL of 104.23 g.

### Sensitivity analysis

Excluding overlapping pregnancies reduced the cohorts to 1,527,385 (GDM, stillbirth and PE) and 936,994 (SGA), while complete-case analysis further restricted the sample sizes to 793,031 (GDM), 792,401 (PE), 1,178,044 (stillbirth) and 502,967 (SGA). Both sensitivity analyses showed minimal changes in characteristic distributions and yielded validation results consistent with the primary analysis (Additional file 1: Tables S6–S9; Additional file 2: Figure S3). For models without reported intercepts, 12 models from seven studies were evaluated for discrimination alone (eight for GDM and four for PE). Validated C-statistics ranged from 0.67 to 0.77 for GDM, and from 0.69 to 0.73 for PE (any PE: 0.69; early-onset PE: 0.73; intermediate PE: 0.71; late PE: 0.69). Model information and validation results are presented in Additional file 1: Table S10.

## Discussion

### Main findings and Interpretation

This study represents a comprehensive external validation of clinical prediction models (CPMs) for adverse pregnancy outcomes using a large, nationally representative UK linked primary and hospital care dataset. Most existing models, when applied to the UK population data, demonstrated suboptimal generalisability. Specifically, most models showed suboptimal discrimination and/or calibration. For low prevalence outcomes, such as stillbirth, only a limited number of models were available, and their performance was consistently poor. The observed decline in performance is likely attributable to several factors. Firstly, significant heterogeneity exists between our validation cohort and the original model development populations. Many of the validated models were developed in small, hospital-based, or prospectively recruited cohorts with different demographic features, outcome prevalences, and ethnic compositions. Secondly, some models used a limited number of predictors, with as few as three predictors. This may be insufficient to capture all useful information leading to adverse outcomes, thus reducing predictive power when applied to a new, broader population.

### Strengths and Limitations

This study used a large, nationally representative electronic health record dataset of the UK population, providing a realistic sample and ample statistical power for certainty in our performance estimates. This enhances the generalisability of the findings to UK clinical settings and supports their relevance (or lack of for some models) for clinical application. The systematic approach to identifying relevant models and the comprehensive sensitivity analysis conducted to explore potential reasons for performance degradation add to the overall robustness of the study. However, some limitations inherent to the use of retrospective, routinely collected data are evident. The inaccuracies in clinical coding and misclassification bias cannot be entirely ruled out. Although missing data was addressed with multiple imputation, high levels of missingness for certain predictors (such as height and weight) remain a concern, and imputation cannot fully compensate for the information lost due to missingness. The results from sensitivity analyses were, however, largely consistent with the primary analyses, suggesting the strategies for handling missing observations and algorithm derived overlapping pregnancies did not introduce significant bias. Finally, whilst our findings are highly relevant to the UK population, their generalisability to populations in other countries may be limited.

### Implications for Clinical Practice and Future Research

This extensive external validation work has several implications for clinical prediction models if they are to be used to improve antenatal risk stratification. Many existing models were identified with several having acceptable discrimination in our validation, indicating that routinely collected, early maternal information contains meaningful predictive signal associated with developing complications or poor outcomes. This aligns with previous evidence showing that the routinely collected primary care data has strong association with pregnancy complications [20], and models based solely on clinical characteristics can achieve performance comparable to those incorporating biomarkers or ultrasound [11]. However, despite their discriminative potential, most models showed limited generalisability, failing to achieve adequate discrimination and calibration simultaneously, with pronounced overestimation of risk. Similar limitations have been reported across existing validation using international cohorts [11, 12, 19, 36], suggesting overfitting in original development studies and highlighting that most models cannot be directly applied to a new, larger population without adjustment. Applying such poorly performing tools in the pre-conception or early pregnancy window could lead to unnecessary investigation and intervention for low-risk pregnancies and misunderstanding of the true at-risk group.

Model performance inherently varies across different time periods, geographical locations, and demographic structures. When model performance is optimal and implemented in practice, monitoring of model performance is required to detect any future shifts in these factors, ensuring models remain clinically relevant and reliable in evolving healthcare settings [37]. Given the suboptimal generalisability of models within the UK population, our future work will explore the potential for performance improvement by prioritising re-calibration or where necessary, model updating, to prevent research waste. The development of entirely new models should only be considered if updating yields insufficient improvements or if substantial structural modifications are required [38].

Furthermore, many pregnancy complications and adverse outcomes are complex and interrelated. For example, both gestational diabetes and hypertensive disorders of pregnancy can increase the risk of FGR, and FGR is strongly associated with an elevated risk of stillbirth [39]. Therefore, methods to predict the risk of individuals developing multiple outcomes by capturing these associations may enhance the predictive abilities and provide a more holistic assessment of pre- or early pregnancy risks. Importantly, studies should also examine whether these models retain predictive accuracy across diverse populations, healthcare settings, and time periods, whilst also assessing the accuracy of key outcome components and identifying potential inconsistencies among predictors [40]. Such work will be essential to ensure that prediction models are both clinically applicable and reliable for improving maternal and perinatal outcomes.

## Conclusions

In this large-scale external validation of clinical prediction models for adverse pregnancy outcomes using pre-conception or early pregnancy routine health data, most existing models demonstrated suboptimal discrimination and/or calibration, particularly for rare outcomes such as stillbirth. These findings indicate that currently available models lack generalisability to real-world clinical practice in the UK and should not be implemented without consideration of model re-calibration, or model updating, before consideration of re-development.

## Supporting information

PRISMA checklist

TRIPOD checklist

## List of abbreviations

AUC: area under the receiver operating characteristic curve
BMI: body mass index
CI: confidence interval
CITL: calibration-in-the-large
CPM: clinical prediction model
CPRD: Clinical Practice Research Datalink
EOPE: early-onset pre-eclampsia
FGR: fetal growth restriction
FMF: Fetal Medicine Foundation
GDM: gestational diabetes mellitus
HES-APC: Hospital Episode Statistics Admitted Patient Care
ICD-10: International Classification of Diseases, 10th Revision
LOPE: late-onset pre-eclampsia
LP: linear predictor
MICE: multiple imputation by chained equations
NHS: National Health Service
O/E: observed-to-expected
PE: pre-eclampsia
PRISMA: Preferred Reporting Items for Systematic Reviews and Meta-Analyses
SD: standard deviation
SGA: small-for-gestational-age
SNOMED CT: Systematized Nomenclature of Medicine Clinical Terms
TRIPOD+AI: Transparent Reporting of a multivariable prediction model for Individual Prognosis or Diagnosis + Artificial Intelligence
UK: United Kingdom
WHO: World Health Organization

## Declarations

### Ethics approval and consent to participate

This study used anonymised electronic health records from the Clinical Practice Research Datalink (CPRD). In accordance with CPRD research approval, the study protocol was approved by the Independent Scientific Advisory Committee (ISAC) for CPRD research (protocol number: 22_002169).

### Consent for publication

Not applicable.

### Availability of data and materials

The data that support the findings of this study are available from Clinical Practice Research Datalink (CPRD), but restrictions apply to the availability of these data, which were used under license for the current study, and so are not publicly available. For more information see https://cprd.com/data-access. Linked secondary care data from Hospital Episode Statistics, mortality data from the Office for National Statistics, and index of multiple deprivation data can also be requested from CPRD.

To facilitate transparency and reproducibility, all clinical code lists and statistical analysis scripts used in this study have been made openly accessible and can be found at the following GitHub repositories:

- Code list to build outcomes and predictors: https://github.com/YiranZhang1014/cprd-aurum-pregnancy-codelist
- Analysis codes: https://github.com/YiranZhang1014/pregnancy-model-validation

## Competing interests

The authors declare that they have no competing interests.

## Funding

Darren M Ashcroft is funded by the National Institute for Health and Care Research (NIHR) Greater Manchester Patient Safety Research Collaboration (NIHR204295).

## Authors’ contributions

All authors contributed to the study conception and design. Approvals were obtained by Victoria Palin (VP), Glen Philip Martin (GPM), Tjeerd van Staa (TvS), Darren M Ashcroft (DMA). Intellectual input on design and analysis from VP, GPM, TvS, DMA, Yiran Zhang (YZ). Predefined eligibility criteria and a structured search strategy were defined and reviewed by all authors. With oversight from all authors, YZ conducted the systematic review and primary data analysis. YZ prepared the first version of the manuscript and all authors contributed to revisions and approved the final manuscript.

## Data Availability

No data are available. Electronic health records are, by definition, considered sensitive data in the UK by the Data Protection Act and cannot be shared by public deposition because of information governance restriction in place to protect patient confidentiality. Access for other research data from Clinical Practice Research Datalink (CPRD) is subject to protocol approval via research data governance process of CPRD. For more information see https://cprd.com/data-access. Linked secondary care data from Hospital Episodes Statistics, mortality data from the Office for National Statistics, and index of multiple deprivation data can also be requested from CPRD.

## Acknowledgements

This study is based on data from the Clinical Practice Research Datalink obtained under licence from the UK Medicines and Healthcare Products Regulatory Agency (MHRA). The data are provided by patients and collected by the NHS as part of their care and support. Hospital Episode Statistics and Office for National Statistics mortality data are subject to Crown copyright (2025) protection, reused with the permission of the Health and Social Care Information Centre, all rights reserved. The interpretation and conclusions in this study are those of the authors alone, and not necessarily those of the MHRA, National Institute for Health and Care Research, NHS, or Department of Health and Social Care. The study protocol was approved by Clinical Practice Research Datalink’s independent scientific advisory committee (reference: 22_002169). We acknowledge all of the data providers and general practices who make anonymised data available for research.

## Additional files

**Additional file 1** (.docx): Supplementary Tables S1-S10. Literature search strategy, characteristics and equations of eligible models, existing external validation studies, cohort and predictor characteristics, validation results for the primary, non-overlapping and complete-case analyses.

**Additional file 2** (.docx): Supplementary Figures S1-S3. Cohort derivation flow chart; calibration plots for all models in the primary analysis; distribution of validation metrics in sensitivity analyses.

**Additional file 3** (.docx): Supplementary Table S11. Excluded models and reasons.

## Notes

### Competing Interest Statement

The authors have declared no competing interest.

### Author Declarations

The study protocol was reviewed and approved by the Independent Scientific Advisory Committee (ISAC) for CPRD research (protocol number: 22_002169) before commencement.

### Summary of Updates

The manuscript and supplementary files have been restructured and reformatted to align with BMC Medicine submission requirements (including updated PRISMA 2020 and TRIPOD+AI checklists). We added a new discrimination-only sensitivity analysis evaluating 12 models that reported complete predictor coefficients without an intercept (Additional file 1: Table S10). We also updated the minimum required sample size and event calculations for both our external validation cohort and previously published external validation studies using the corrected Riley et al. Finally, we corrected minor inaccuracies in model specifications and ensured consistency across the main text, tables, figures, and Additional files 1 to 3.

## References

1. Haley GG, Dana BM: The prevalence of sporadic and recurrent pregnancy loss. Fertility and Sterility 2023, 120(5):934–936.

2. Hypertension in pregnancy: Background information - Prevalence [https://cks.nice.org.uk/topics/hypertension-in-pregnancy/background-information/prevalence/]

3. National Institute for Health and Care Excellence: Diabetes in pregnancy: management from preconception to the postnatal period. In. London; 2020.

4. Office for National Statistics: Births in England and Wales: 2024. In. Newport, UK; 2025.

5. Schlembach D: Fetal Growth Restriction - Diagnostic Work-up, Management and Delivery. Geburtshilfe Frauenheilkd 2020, 80(10):1016–1025.

6. Islam MN, Mustafina SN, Mahmud T, Khan NI: Machine learning to predict pregnancy outcomes: a systematic review, synthesizing framework and future research agenda. BMC Pregnancy Childbirth 2022, 22(1):348.

7. van Eekhout JCA, Becking EC, Scheffer PG, Koutsoliakos I, Bax CJ, Henneman L, Bekker MN, Schuit E: First-Trimester Prediction Models Based on Maternal Characteristics for Adverse Pregnancy Outcomes: A Systematic Review and Meta-Analysis. BJOG 2025, 132(3):243–265.

8. Guy GP, Leslie K, Diaz Gomez D, Forenc K, Buck E, Khalil A, Thilaganathan B: Implementation of routine first trimester combined screening for pre-eclampsia: a clinical effectiveness study. BJOG 2021, 128(2):149–156.

9. Tan MY, Wright D, Syngelaki A, Akolekar R, Cicero S, Janga D, Singh M, Greco E, Wright A, Maclagan K et al: Comparison of diagnostic accuracy of early screening for pre-eclampsia by NICE guidelines and a method combining maternal factors and biomarkers: results of SPREE. Ultrasound Obstet Gynecol 2018, 51(6):743–750.

10. Allotey J, Archer L, Coomar D, Snell KI, Smuk M, Oakey L, Haqnawaz S, Betrán AP, Chappell LC, Ganzevoort W et al: Development and validation of prediction models for fetal growth restriction and birthweight: an individual participant data meta-analysis. Health Technol Assess 2024, 28(47):1–119.

11. Allotey J, Snell KI, Smuk M, Hooper R, Chan CL, Ahmed A, Chappell LC, von Dadelszen P, Dodds J, Green M et al: Validation and development of models using clinical, biochemical and ultrasound markers for predicting pre-eclampsia: an individual participant data meta-analysis. Health Technol Assess 2020, 24(72):1–252.

12. Allotey J, Whittle R, Snell KIE, Smuk M, Townsend R, von Dadelszen P, Heazell AEP, Magee L, Smith GCS, Sandall J et al: External validation of prognostic models to predict stillbirth using International Prediction of Pregnancy Complications (IPPIC) Network database: individual participant data meta-analysis. Ultrasound Obstet Gynecol 2022, 59(2):209–219.

13. Snell KIE, Allotey J, Smuk M, Hooper R, Chan C, Ahmed A, Chappell LC, Von Dadelszen P, Green M, Kenny L et al: External validation of prognostic models predicting pre-eclampsia: individual participant data meta-analysis. BMC Med 2020, 18(1):302.

14. Townsend R, Manji A, Allotey J, Heazell A, Jorgensen L, Magee LA, Mol BW, Snell K, Riley RD, Sandall J et al: Can risk prediction models help us individualise stillbirth prevention? A systematic review and critical appraisal of published risk models. BJOG 2021, 128(2):214–224.

15. Lamain-de Ruiter M, Kwee A, Naaktgeboren CA, Franx A, Moons KGM, Koster MPH: Prediction models for the risk of gestational diabetes: a systematic review. Diagn Progn Res 2017, 1:3.

16. Binuya MAE, Engelhardt EG, Schats W, Schmidt MK, Steyerberg EW: Methodological guidance for the evaluation and updating of clinical prediction models: a systematic review. BMC Med Res Methodol 2022, 22(1):316.

17. Zhou ZR, Wang WW, Li Y, Jin KR, Wang XY, Wang ZW, Chen YS, Wang SJ, Hu J, Zhang HN et al: In-depth mining of clinical data: the construction of clinical prediction model with R. Ann Transl Med 2019, 7(23):796.

18. Moons KG, Altman DG, Reitsma JB, Ioannidis JP, Macaskill P, Steyerberg EW, Vickers AJ, Ransohoff DF, Collins GS: Transparent Reporting of a multivariable prediction model for Individual Prognosis or Diagnosis (TRIPOD): explanation and elaboration. Ann Intern Med 2015, 162(1):W1–73.

19. Meertens L, Smits L, van Kuijk S, Aardenburg R, van Dooren I, Langenveld J, Zwaan IM, Spaanderman M, Scheepers H: External validation and clinical usefulness of first-trimester prediction models for small- and large-for-gestational-age infants: a prospective cohort study. BJOG 2019, 126(4):472–484.

20. Schoenaker D, Lovegrove EM, Cassinelli EH, Hall J, McGranahan M, McGowan L, Carr H, Alwan NA, Stephenson J, Godfrey KM: Preconception indicators and associations with health outcomes reported in UK routine primary care data: a systematic review. Br J Gen Pract 2025, 75(751):e129–e136.

21. Page MJ, McKenzie JE, Bossuyt PM, Boutron I, Hoffmann TC, Mulrow CD, Shamseer L, Tetzlaff JM, Akl EA, Brennan SE et al: The PRISMA 2020 statement: an updated guideline for reporting systematic reviews. Bmj 2021, 372:n71.

22. Collins GS, Moons KGM, Dhiman P, Riley RD, Beam AL, Van Calster B, Ghassemi M, Liu X, Reitsma JB, van Smeden M et al: TRIPOD+AI statement: updated guidance for reporting clinical prediction models that use regression or machine learning methods. Bmj 2024, 385:e078378.

23. Gordijn SJ, Beune IM, Thilaganathan B, Papageorghiou A, Baschat AA, Baker PN, Silver RM, Wynia K, Ganzevoort W: Consensus definition of fetal growth restriction: a Delphi procedure. Ultrasound Obstet Gynecol 2016, 48(3):333–339.

24. CPRD Linked Data [https://www.cprd.com/data/linked-data]

25. CPRD Aurum June 2021 dataset [https://www.cprd.com/cprd-aurum-june-2021]

26 . Kiserud T, Piaggio G, Carroli G, Widmer M, Carvalho J, Neerup Jensen L, Giordano D, Cecatti JG, Abdel Aleem H, Talegawkar SA et al: The World Health Organization Fetal Growth Charts: A Multinational Longitudinal Study of Ultrasound Biometric Measurements and Estimated Fetal Weight. PLoS Med 2017, 14(1):e1002220.

27. Fetal Growth Calculator [https://srhr.org/fetalgrowthcalculator/]

28. Riley RD, Debray TPA, Collins GS, Archer L, Ensor J, van Smeden M, Snell KIE: Minimum sample size for external validation of a clinical prediction model with a binary outcome. Statistics in Medicine 2021, 40(19):4230–4251.

29. Sterne JA, White IR, Carlin JB, Spratt M, Royston P, Kenward MG, Wood AM, Carpenter JR: Multiple imputation for missing data in epidemiological and clinical research: potential and pitfalls. Bmj 2009, 338:b2393.

30. Gehringer CK, Martin GP, Van Calster B, Hyrich KL, Verstappen SMM, Sergeant JC: How to develop, validate, and update clinical prediction models using multinomial logistic regression. J Clin Epidemiol 2024, 174:111481.

31. Zou G: A modified poisson regression approach to prospective studies with binary data. Am J Epidemiol 2004, 159(7):702–706.

32. Li Q, Li P, Chen J, Ren R, Ren N, Xia Y: Machine Learning for Predicting Stillbirth: A Systematic Review. Reprod Sci 2025, 32(5):1388–1398.

33. Yerlikaya G, Akolekar R, McPherson K, Syngelaki A, Nicolaides KH: Prediction of stillbirth from maternal demographic and pregnancy characteristics. Ultrasound Obstet Gynecol 2016, 48(5):607–612.

34. Syngelaki A, Wright A, Gomez Fernandez C, Mitsigiorgi R, Nicolaides KH: First-Trimester Prediction of Gestational Diabetes Mellitus Based on Maternal Risk Factors. Bjog 2025, 132(7):972–982.

35. Poon LC, Karagiannis G, Staboulidou I, Shafiei A, Nicolaides KH: Reference range of birth weight with gestation and first-trimester prediction of small-for-gestation neonates. Prenat Diagn 2011, 31(1):58–65.

36. Meertens LJE, Scheepers HCJ, van Kuijk SMJ, Roeleveld N, Aardenburg R, van Dooren IMA, Langenveld J, Zwaan IM, Spaanderman MEA, van Gelder M, Smits LJM: External validation and clinical utility of prognostic prediction models for gestational diabetes mellitus: A prospective cohort study. Acta Obstet Gynecol Scand 2020, 99(7):891–900.

37. Finlayson SG, Subbaswamy A, Singh K, Bowers J, Kupke A, Zittrain J, Kohane IS, Saria S: The Clinician and Dataset Shift in Artificial Intelligence. N Engl J Med 2021, 385(3):283–286.

38. Steyerberg EW, Moons KG, van der Windt DA, Hayden JA, Perel P, Schroter S, Riley RD, Hemingway H, Altman DG: Prognosis Research Strategy (PROGRESS) 3: prognostic model research. PLoS Med 2013, 10(2):e1001381.

39. Flenady V, Koopmans L, Middleton P, Frøen JF, Smith GC, Gibbons K, Coory M, Gordon A, Ellwood D, McIntyre HD et al: Major risk factors for stillbirth in high-income countries: a systematic review and meta-analysis. Lancet 2011, 377(9774):1331–1340.

40. Efthimiou O, Seo M, Chalkou K, Debray T, Egger M, Salanti G: Developing clinical prediction models: a step-by-step guide. Bmj 2024, 386:e078276.

41. Sepúlveda-Martínez A, Rencoret G, Silva MC, Ahumada P, Pedraza D, Muñoz H, Valdés E, Parra-Cordero M: First trimester screening for preterm and term pre-eclampsia by maternal characteristics and biophysical markers in a low-risk population. J Obstet Gynaecol Res 2019, 45(1):104–112.

42. Rocha RS, Alves JAG, Maia EHMSB, Araujo Júnior E, Peixoto AB, Santana EFM, Martins WP, Vasconcelos CTM, Da Silva Costa F, Oriá MOB: Simple approach based on maternal characteristics and mean arterial pressure for the prediction of preeclampsia in the first trimester of pregnancy. J Perinat Med 2017, 45(7):843–849.

43. Lai J, Poon LC, Pinas A, Bakalis S, Nicolaides KH: Uterine artery Doppler at 30-33 weeks’ gestation in the prediction of preeclampsia. Fetal Diagn Ther 2013, 33(3):156–163.

44. Trudell AS, Tuuli MG, Colditz GA, Macones GA, Odibo AO: A stillbirth calculator: Development and internal validation of a clinical prediction model to quantify stillbirth risk. PLoS One 2017, 12(3):e0173461.

45. Adjahou S, Syngelaki A, Nanda M, Papavasileiou D, Akolekar R, Nicolaides KH: Routine 36-week scan: prediction of small-for-gestational-age neonate. Ultrasound Obstet Gynecol 2025, 65(1):20–29.

