## Additional file 1 for "Systematic Review and External Validation of Clinical Prediction Models for Adverse Pregnancy Outcomes Using Routinely Collected Pre-Conception and Early Pregnancy Data"

**Note: References in this document correspond to the reference list at the end of this file.**

Additional file 1: Table S1 (a): Literature Search Keywords

| Strategy | Keywords |
|---|---|
| [Systematic review, meta-analysis, or external validation] | ("systematic review"[tiab]<br>OR "external validation"[tiab]<br>OR "externally validat*"[tiab]<br>OR "external evaluation"[tiab]<br>OR "externally evaluat*"[tiab]<br>OR "meta analysis"[tiab]<br>OR "meta-analysis"[tiab]) |
| [Prediction] | ("predictive model"[tiab]<br>OR "predictive models"[tiab]<br>OR "prediction"[tiab]<br>OR "predictions"[tiab]<br>OR "risk calculator"[tiab]<br>OR "risk calculators"[tiab]<br>OR "risk model"[tiab]<br>OR "risk models"[tiab]<br>OR "risk score"[tiab]<br>OR "risk scores"[tiab]<br>OR "algorithm"[tiab]<br>OR "algorithms"[tiab]<br>OR "risk assessment"[tiab]<br>OR "risk assessments"[tiab]<br>OR "nomogram"[tiab]<br>OR "nomograms"[tiab]<br>OR "prognostic model"[tiab]<br>OR "prognostic models"[tiab]<br>OR "scoring system"[tiab]<br>OR "scoring systems"[tiab]<br>OR "screening model"[tiab]<br>OR "screening models"[tiab]<br>OR "decision rule"[tiab]<br>OR "decision rules"[tiab]) |
| [Gestational diabetes] | ("gestational diabetes"[tiab]<br>OR "pregnancy induced diabetes"[tiab]<br>OR "pregnancy-induced diabetes"[tiab]<br>OR "Diabetes, Gestational"[Mesh]) |
| [Pre-eclampsia] | ("preeclamp*"[tiab]<br>OR "pre-eclamp*"[tiab]<br>OR "Pre-Eclampsia"[Mesh]) |

| | |
|---|---|
| [Fetal growth restriction or small for gestational age] | ("fetal growth restriction"[tiab]<br>OR "foetal growth restriction"[tiab]<br>OR "fetal Growth Retardation"[tiab]<br>OR "foetal Growth Retardation"[tiab]<br>OR "FGR"[tiab]<br>OR "Intrauterine Growth Restriction"[tiab]<br>OR "Intrauterine Growth Retardation"[tiab]<br>OR "IUGR"[tiab]<br>OR "Small for gestational age"[tiab]<br>OR "Small-for-gestational-age"[tiab]<br>OR "SGA"[tiab]<br>OR "Fetal Growth Retardation"[MeSH]<br>OR "Infant, Small for Gestational Age"[MeSH]) |
| [Stillbirth] | ("stillbirth*"[tiab]<br>OR "fetal death"[tiab]<br>OR "foetal death"[tiab]<br>OR "perinatal death"[tiab]<br>OR "intrauterine death"[tiab]<br>OR "intrauterine fetal death"[tiab]<br>OR "intrauterine foetal death"[tiab]<br>OR "perinatal fetal death"[tiab]<br>OR "fetal mortality"[tiab]<br>OR "foetal mortality"[tiab]<br>OR "intrauterine fetal mortality"[tiab]<br>OR "perinatal fetal mortality"[tiab]<br>OR "perinatal foetal mortality"[tiab]<br>OR "Stillbirth"[Mesh]) |

Additional file 1: Table S1 (b): Literature Search Strategy

| | | Gestational diabetes (GDM) | Stillbirth | Pre-eclampsia (PE) | Fetal growth restriction (FGR) or small for gestational age (SGA) |
|---|---|---|---|---|---|
| **Stage 1** | **Search time** | Until 2025-06-01 | Until 2025-06-01 | Until 2025-06-01 | Until 2025-06-01 |
| | **Search keywords** | [Systematic review, meta-analysis, or external validation]<br>AND<br>[Prediction]<br>AND<br>[Gestational diabetes] | [Systematic review, meta-analysis, or external validation]<br>AND<br>[Prediction]<br>AND<br>[Stillbirth] | [Systematic review, meta-analysis, or external validation]<br>AND<br>[Prediction]<br>AND<br>[Pre-eclampsia] | [Systematic review, meta-analysis, or external validation]<br>AND<br>[Prediction]<br>AND<br>[Fetal growth restriction or small for gestational age] |
| **Stage 2** | **Search time** | 2024-06-06 - 2025-06-01 | 2022-10-18 - 2025-06-01 | 2024-06-06 - 2025-06-01 | 2024-06-06 - 2025-06-01 |
| | **Search keywords** | [Prediction]<br>AND<br>[Gestational diabetes] | [Prediction]<br>AND<br>[Stillbirth] | [Prediction]<br>AND<br>[Pre-eclampsia] | [Prediction]<br>AND<br>[Fetal growth restriction or small for gestational age] |

Additional file 1: Table S2 Information of eligible models in external validation

| Outcome | Model | Model type | Equation | Number of predictors, N | Population Source | Original data type | Sample size (cases), N | Reported C-Statistics | Estimated sample size (cases) required for primary analysis, N |
|---|---|---|---|---|---|---|---|---|---|
| Gestational diabetes (GDM) | van Leeuwen 2010 [1] | Logistic regression | logit(p) = -6.1 + 0.83*(non-Caucasian ethnicity) + 0.57*(family history of diabetes) - 0.67*(parous without previous gestational diabetes) + 0.5*(parous with previous gestational diabetes) + 0.13*(BMI) | 5 | Netherlands | Prospective | 995 (24) | 0.77 | 10,309 (421) |
| | Sweeting 2017 [2] | Logistic regression | logit(p) = -5.9515 + 2.8624*(previous GDM) + 1.55*(east Asian ethnicity) + 1.7033*(south Asian) + 3.0678*(family history of diabetes) - 0.5836*(parity) + 0.0354*(maternal age) + 0.08462*(BMI) | 7 | Australia | Retrospective | 980 (248) | 0.88 | 9,039 (369) |
| | Benhalima 2020 [3] | Logistic regression | logit(p) = -1.39 + 0.08*(maternal age) + 0.41*(family history of diabetes) + 1.19*(Asian ethnicity) + 0.35*(smoke) + 2.05*(previous gestational diabetes) + 0.07*(BMI) - 3.40*(height in m) | 7 | Belgium | Prospective | 1,843 (231) | 0.72 | 15,547 (635) |
| | Basil 2024 [4] | Logistic regression | logit(p) = - 6.358 + 0.066*(maternal age) + 0.075*(BMI) + 1.879*(first degree family history of diabetes) + 0.522*(previous fetal macrosomia) | 4 | Nigeria | Prospective | 253 (52) | 0.814 | 9,039 (369) |
| | Syngelaki 2025 (for nulliparous) [5] | Logistic regression | logit(p) = -2.4928 + 0.0359*(weight - 69 kg) - 0.0602*(height - 164 cm) + 0.0599*(maternal age - 35 years) + 0.3660*(Black ethnicity) + 1.2375*(east Asian ethnicity) + 0.9245*(south Asian ethnicity) + 0.4455*(family history of diabetes) | 7 | UK | Prospective | 20,549 (1,787) | 0.757 | 11,366 (464) |
| | Syngelaki 2025 (for parous without previous GDM) [5] | Logistic regression | logit(p) = -2.6949 + 0.2162*(previous birthweight z-score) + 0.0359*(weight - 69 kg) - 0.0602*(height - 164 cm) + 0.0599*(maternal age - 35 years) + 0.3660*(Black ethnicity) + 1.2375*(east Asian ethnicity) + 0.9245*(south Asian ethnicity) + 0.4455*(family history of diabetes) | 8 | UK | Prospective | 20,099 (1,830) | 0.757 | 11,366 (464) |

| | | | | | | | | | |
|---|---|---|---|---|---|---|---|---|---|
| | Syngelaki 2025 (For parous with previous GDM) [5] | Logistic regression | logit(p) = -0.2608 + 0.0176*(weight - 69 kg) - 0.0199*(height - 164 cm) + 0.0496*(maternal age - 35 years) + 0.3277*(Black ethnicity) + 0.6754*(east Asian ethnicity) + 0.5292*(south Asian ethnicity) + 0.3942*(family history of diabetes) | 7 | UK | Prospective | 1,205 (667) | 0.757 | 11,366 (464) |
| Pre-eclampsia (PE) | Plasencia 2007 for Any PE [6] | Logistic regression | logit(p) =-6.253 + 1.432*(Afro-Caribbean ethnicity) + 1.465*(mixed ethnicity) + 0.084*(BMI) + 0.81*(family history of pre-eclampsia) + 1.049*(parous with a history of pre-eclampsia) - 1.539*(parous with no history of pre-eclampsia) | 6 | UK | Prospective | 6,015 (107) | 0.806 | 17,092 (377) |
| | Emonts 2008 for Any PE [7] | Logistic regression | logit(p) = -3.72 + 0.030*(maternal age) - 0.50*(parity) + 0.15*(gestation) + 1.89*(family history of hypertension) + 0.14*(BMI) + 0.079*(systolic blood pressure) - 0.13*(diastolic blood pressure) | 7 | Belgium | Retrospective | 151 (101) | Not reported | Not available |
| | Poon 2008 for Any PE (1) [8] | Logistic regression | logit(p) = -8.0469 + 0.0552*(maternal age) + 0.0738*(BMI) - 1.9074*(parous with no history of pre-eclampsia) + 1.2328*(parous with history of pre-eclampsia) + 1.164*(Family history of pre-eclampsia) + 2.2196*(Afro-Caribbean ethnicity) + 1.506*(Indian or Pakistani or Bangladeshi ethnicity) + 1.7842*(mixed ethnicity) | 8 | UK | Prospective | 2,486 (51) | 0.852 | 17,092 (377) |
| | Poon 2008 for Any PE (2) [9] | Logistic regression | logit(p) = -6.311 + 1.299*(Afro-Caribbean ethnicity) + 0.092*(BMI) + 0.855*(family history of pre-eclampsia) - 1.481*(parous with no history of pre-eclampsia) + 0.933*(parous with a history of pre-eclampsia) | 5 | UK | Prospective | 5,193 (104) | 0.801 | 17,092 (377) |
| | Direkvand-Moghadam 2013 for Any PE [10] | Logistic regression | logit(p) = -0.74 + 1.016*(infertility) + 0.72*(chronic hypertension) + 1.69*(previous pre-eclampsia) | 3 | Iran | Retrospective | 610 (58) | 0.67 | 47,128 (1,037) |
| | Macdonald-Wallis 2015 for Any PE [11] | Logistic regression | logit(p) = -3.46 + 0.046*(mean arterial pressure) + 0.057*(BMI) - 0.024*(height) + 0.384*(maternal age ≥35) - 1.590*(parity = 1) - 1.546*(parity ≥ 2) - 0.479*(smoke) + 0.793*(hypertension) + 1.346*(pre-gestational hypertension) + 2.301*(diabetes) + 0.941*(pre-gestational diabetes) + 0.429*(non-white ethnicity) | 12 | UK | Prospective | 12,996 (317) | 0.77 | 17,812 (392) |

| | | | | | | | | | |
|---|---|---|---|---|---|---|---|---|---|
| | Rocha 2017 for Any PE (a) [12] | Logistic regression | logit(p) = -0.189 + 0.011*(BMI) + 0.127*(family history of pre-eclampsia) - 0.062*(no history of pre-eclampsia) | 3 | Brazil | Prospective | 733 (55) | 0.747 | 21,492 (473) |
| | Rocha 2017 for Any PE (b) [12] | Logistic regression | logit(p) = -0.426 + 0.006*(BMI) + 0.129*(family history of pre-eclampsia) - 0.059*(no history of pre-eclampsia) + 0.004*(mean arterial pressure) | 4 | Brazil | Prospective | 733 (55) | 0.787 | 17,092 (377) |
| | Allotey 2020 for Any PE (a) [13] # | Logistic regression | logit(p) = -6.5297 - 0.0151*(maternal age) + 0.0156*(systolic blood pressure) + 0.0370*(BMI) + 0.9287*(nulliparous) + 1.2574*(previous pre-eclampsia) + 1.5019*(renal disease) + 1.5689*(chronic hypertension) + 0.3578*(pre-existing diabetes) | 8 | Multiple countries (1) | Retrospective | 29,187 (1,187) | 0.677 | 43,329 (954) |
| | Allotey 2020 for Any PE (b) [13] $ | Logistic regression | logit(p) = -8.0909 - 0.0165*(maternal age) + 0.0108*(systolic blood pressure) + 0.0198*(diastolic blood pressure) + 0.0614*(BMI) + 0.9616*(nulliparous) + 1.4068*(previous pre-eclampsia) + 0.9700*(renal disease) + 1.7763*(chronic hypertension) | 8 | Multiple countries (1) | Retrospective | 29,153 (1,144) | 0.703 | 32,518 (716) |
| | Tang 2022 for Any PE [14] | Logistic regression | logit(p) = -13.40 + 0.102*(BMI) - 0.920*(parous) + 0.0445*(maternal age) + 0.433*(diabetes) + 0.320*(family history of hypertension) + 0.0551*(mean arterial pressure) | 6 | China | Retrospective | 20,582 (717) | 0.71 | 30,279 (667) |
| | Tarca 2022 for Any PE [15] | Poisson regression | ln(p) = -3.3940 + 1.6034*(chronic hypertension) + 0.0049*(weight - 75 kg) + 0.5007*(nulliparity) + 0.8986*(history of pre-eclampsia) | 4 | USA | Retrospective | 1,150 (230) | 0.70 | 33,552 (739) |
| | Plasencia 2007 for EOPE [6] | Logistic regression | logit(p) = -6.431 + 1.680*(Afro-Caribbean ethnicity) + 1.889*(mixed ethnicity) + 2.822*(parous with a history of pre-eclampsia) | 3 | UK | Prospective | 6,015 (not reported) | 0.784 | 182,696 (384) |
| | Kuc 2013 for EOPE [16] | Logistic regression | logit(p) = -6.790 - 0.119*(height) + 4.8565*(ln(weight)) + 1.845*(nulliparous) + 0.086*(maternal age) + 1.353*(smoke) | 5 | Netherlands | Retrospective | 568 (68) | Not reported | Not available |
| | Scazzocchio 2013 for EOPE [17] | Logistic regression | logit(p) = -7.703 + 0.086*(BMI) + 1.708*(chronic hypertension) + 4.033*(renal disease) + 1.931*(parous with a history of pre-eclampsia) + 0.005*(parous with no history of pre-eclampsia) | 5 | Spain | Prospective | 5,170 (26) | Not reported | Not available |

| | | | | | | | | | |
|---|---|---|---|---|---|---|---|---|---|
| | Baschat 2014 for EOPE [18] | Logistic regression | logit(p) = -5.803 + 0.302*(diabetes) + 0.767*(chronic hypertension) + 0.00948*(mean arterial pressure) | 3 | USA | Prospective | 2,441 (18) | 0.83 | 182,696 (384) |
| | Teixeira 2014 for EOPE [19] | Logistic regression | logit(p) = -4.951 + 1.519*(Chronic hypertension) - 1.201*(multiparous) + 3.201*(History of PE) + 7.108*(log10(Weight MoM)) | 4 | Portugal | Retrospective | 4,799 (35) | 0.754 | 195,361 (411) |
| | Crovetto 2015 for EOPE [20] | Logistic regression | logit(p) = −5.177 + 2.383*(Black) - 1.105*(nulliparous) + 3.543*(parous pre-eclampsia) + 2.229*(chronic hypertension) + 2.201*(renal disease) | 5 | Spain | Prospective | 9,462 (57) | 0.757 | 190,099 (400) |
| | Allotey 2020 for EOPE (a) [13] [#] | Logistic regression | logit(p) = -7.9161 - 0.0360*(maternal age) - 0.0176*(systolic blood pressure) + 0.0753*(diastolic blood pressure) - 0.5937*((BMI/10)$^{-2}$) + 1.7440*(previous pre-eclampsia) + 2.2795*(renal disease) + 0.5486*(chronic hypertension) + 1.7185*(diabetes) | 8 | Multiple countries (1) | Retrospective | 29,187 (149) | 0.719 | 262,777 (551) |
| | Allotey 2020 for EOPE (b) [13] [$] | Logistic regression | logit(p) = -10.1805 + 0.0647*(diastolic blood pressure) + 2.3643*(previous pre-eclampsia) + 1.4929*(renal disease) + 1.1445*(autoimmune disease) | 4 | Multiple countries (1) | Retrospective | 29,153 (152) | 0.723 | 202,675 (426) |
| | Plasencia 2007 for LOPE [6] | Logistic regression | logit(p) = -6.585 + 1.368*(Afro-Caribbean ethnicity) + 1.311*(mixed ethnicity) + 0.091*(BMI) + 0.960*(family history of pre-eclampsia) - 1.663*(parous with no history of pre-eclampsia) | 5 | UK | Prospective | 6,015 (not reported) | 0.801 | 18,839 (377) |
| | Poon 2010 for LOPE [21] | Logistic regression | logit(p) = -7.860 + 0.034*(maternal age) + 0.096*(BMI) + 1.089*(Black ethnicity) + 0.980*(Indian/Pakistani ethnicity) + 1.196*(mixed ethnicity) + 1.070*(family history of pre-eclampsia) - 1.413*(parous with no history of pre-eclampsia) + 0.780*(parous with a history of pre-eclampsia) | 8 | UK | Prospective | 8,366 (128) | 0.796 | 18,839 (377) |
| | Kuc 2013 for LOPE [16] | Logistic regression | logit(p) = -14.374 + 2.300*(ln(weight)) + 1.303*(nulliparous) + 0.068*(maternal age) | 3 | Netherlands | Retrospective | 599 (99) | Not reported | Not available |
| | Scazzocchio 2013 for LOPE [17] | Logistic regression | logit(p) = -6.135 + 2.124*(pre-eclampsia) + 1.571*(chronic hypertension) + 0.958*(diabetes) + 1.416*(thrombophilia) - 0.487*(parous) + 0.093*(BMI) | 6 | Spain | Prospective | 5,170 (110) | Not reported | Not available |

| | | | | | | | | | |
|---|---|---|---|---|---|---|---|---|---|
| | Crovetto 2015 for LOPE [20] | Logistic regression | logit(p) = -5.873 - 0.462*(white) + 0.109*(BMI) - 0.825*(nulliparous) + 2.726*(parous pre-eclampsia) + 1.956*(chronic hypertension) - 0.575*(smoke) | 6 | Spain | Prospective | 9,462 (246) | 0.722 | 29,473 (590) |
| | Allotey 2020 for LOPE (a) [13] [#] | Logistic regression | logit(p) = -6.7280 - 0.0138*(maternal age) + 0.0150*(systolic blood pressure) + 0.0355*(BMI) + 1.0363*(nulliparous) + 1.1159*(previous pre-eclampsia) + 1.2487*(renal disease) + 1.6863*(chronic hypertension) | 7 | Multiple countries (1) | Retrospective | 29,187 (1,039) | 0.677 | 47,490 (950) |
| | Allotey 2020 for LOPE (b) [13] [$] | Logistic regression | logit(p) = -8.0911 - 0.0137*(maternal age) + 0.0104*(systolic blood pressure) + 0.0124*(diastolic blood pressure) + 0.0647*(BMI) + 1.1891*(nulliparous) + 1.1650*(previous pre-eclampsia) + 0.7569*(renal disease) + 1.9032*(chronic hypertension) | 8 | Multiple countries (1) | Retrospective | 29,153 (993) | 0.705 | 34,884 (698) |
| | Zhang 2025 for LOPE [22] | Logistic regression | logit(p) = -4.187 + 2.561*(chronic hypertension) + 0.763*(diabetes) + 0.554*(antiphospholipid syndrome) + 1.340*(renal disease) + 0.099*(assisted conception) + 0.943*(nulliparous) + 0.284*(maternal age z-score) + 0.588*(BMI z-score) + 0.810*(mean arterial pressure z-score) | 9 | China | Prospective | 1,728 (110) | 0.833 | 18,839 (377) |
| | Myers 2013 for Preterm PE [23] | Logistic regression | logit(p) = -8.4093 + 0.9037*(assisted conception) + 0.1030*(mean arterial pressure) + 0.7999*(sister with pre-eclampsia) | 3 | Multiple countries (2) | Prospective | 235 (47) | 0.76 | 77,475 (396) |
| | Rocha 2017 for Preterm PE (a) [12] | Logistic regression | logit(p) = -0.051 + 0.109*(chronic hypertension) + 0.083*(history of pre-eclampsia) + 0.003*(BMI) | 3 | Brazil | Prospective | 733 (21) | 0.773 | 75,002 (383) |
| | Rocha 2017 for Preterm PE (b) [12] | Logistic regression | logit(p) = -0.197 + 0.084*(chronic hypertension) + 0.073*(history of pre-eclampsia) + 0.003*(mean arterial pressure) | 3 | Brazil | Prospective | 733 (21) | 0.842 | 75,002 (383) |
| | Sepúlveda-Martínez 2019 for Preterm PE [24] | Logistic regression | logit(p) = -5.354954 + 0.0218775*(maternal age) + 1.534937*(chronic hypertension) + 2.026845*(systemic lupus erythematosus) - 0.1196494*(parous without previous PE) + 2.754261*(parous with previous PE) | 5 | Chile | Prospective | 1,756 (29) | 0.817 | 75,002 (383) |
| | Rocha 2017 for Term PE [12] | Logistic regression | logit(p) = -0.161 + 0.093*(family history of pre-eclampsia) + 0.007*(BMI) + 0.049*(nulliparous) | 3 | Brazil | Prospective | 733 (34) | 0.754 | 25,887 (438) |

| | | | | | | | | | |
|---|---|---|---|---|---|---|---|---|---|
| | Lai 2013 for Intermediate PE (34-37 weeks) [25] | Logistic regression | logit(p) = -5.23982 + 0.031922*(Weight-75kg) - 0.11212*(Height-165cm) + 0.85620*(Afro-Caribbean ethnicity) + 1.58057*(Mixed ethnicity) - 1.012628*(Parous with no PE) + 1.56240*(Chronic hypertension) | 6 | UK | Prospective | 4,855 (37) | 0.771 | 70,814 (383) |
| | Lai 2013 for Late PE [25] | Logistic regression | logit(p) = -4.00255 + 0.020410*(Weight-75kg) - 0.034916*(Height-165cm) + 0.83073*(Afro-Caribbean ethnicity) - 0.77209*(Parous with no PE) + 2.64967*(Chronic hypertension) | 5 | UK | Prospective | 4,855 (108) | 0.756 | 25,454 (431) |
| Stillbirth | Yerlikaya 2016 for stillbirth ≥ 24 weeks [26] | Logistic regression | logit(p) = -6.02615 + 0.01037*(weight - 69 kg) + 0.70027*(Afro-Caribbean ethnicity) + 0.57994*(assisted conception) + 0.53367*(smoking) + 0.96253*(chronic hypertension) + 1.28416*(antiphospholipid syndrome or systemic lupus erythematosus) + 0.93628*(pre-existing diabetes) + 1.57086*(previous stillbirth) | 8 | UK | Prospective | 113,415 (396) | 0.642 | 298,903 (1,465) |
| | Trudell 2017 for stillbirth ≥ 32 weeks [27] | Logistic regression | logit(p) = -6.8772 - 0.8707*(maternal age <= 18) + 0.2094*(maternal age 35-39) + 0.4377*(maternal age ≥ 40) + 0.8536*(Black ethnicity) + 0.3423*(nulliparous) - 0.0219*(BMI 25-29.9) + 0.5607*(BMI 30-34.9) - 0.5948*(BMI 35-39.9) + 0.1593*(BMI ≥ 40) + 0.2270*(smoking) + 0.6255*(chronic hypertension) + 0.9863*(pre-existing diabetes) | 12 | USA | Retrospective | 57,326 (330) | 0.66 | 307,424 (1,138) |
| Small-for-gestational-age (SGA) | Macdonald-Wallis 2015 [11] | Logistic regression | logit(p) = -1.64 + 0.003*(mean arterial pressure) - 0.625*(normal weight) - 0.791*(overweight) - 1.072*(obese) - 0.058*(height in cm) + 0.182*(maternal age >35) - 0.756*(parity = 1) - 0.824*(parity ≥2) + 0.850*(smoking in pregnancy) + 0.447*(chronic hypertension) + 0.209*(previous gestational hypertension) - 1.445*(previous GDM) + 0.856*(non-White ethnicity) | 13 | UK | Prospective | 12,679 (1,205) | 0.7 | 6,923 (932) |
| | González-González 2017 [28] | Logistic regression | logit(p) = -1.732 + 0.299*(nulliparous) + 0.783*(smoking in pregnancy) - 1.743*(pre-existing diabetes) | 3 | Spain | Prospective | 988 (193) | 0.615 | 20,150 (2,711) |
| | Adjahou 2025 [29] | Logistic regression | logit(p) = -2.07540 - 0.02429*(weight – 80 kg) - 0.03429*(height – 165 cm) + 0.71648*(black ethnicity) + 0.68779*(South Asian ethnicity) + 0.03643*(mixed ethnicity) + 0.97590*(cigarette smoker) + 0.91847*(chronic hypertension) - 0.78688*(diabetes | 10 | UK | Retrospective | 107,875 (12,420) | 0.718 | 5,905 (795) |

| | | | | | | | | | |
|---|---|---|---|---|---|---|---|---|---|
| | | | mellitus Type I) - 0.85317*(parous, no previous SGA) + 0.49876*(parous, previous SGA) | | | | | | |
| | Poon 2011 (log(birthweight)) [30] | Linear regression | Log10(birthweight) = -0.935219 + 0.186853*(gestation length) - 0.002078*(gestation length ²) + 0.003726*(weight) - 0.000030*(weight²) + 0.00000008820640*(weight³) + 0.000965*(height) + 0.001466*(maternal age) - 0.000026*(maternal age²) + 0.016986*(parous) - 0.024867*(smoke) - 0.021769*(African ethnicity) - 0.017824*(south Asian) - 0.005543*(East Asian ethnicity) - 0.009063*(mixed ethnicity) - 0.020995*(chronic hypertension) + 0.03143*(pre-existing diabetes) - 0.004015*(assisted conception) | 17 | UK | Prospective | 33,602 | Not applicable | Not applicable |

**Note:**

**EOPE:** early-onset pre-eclampsia - delivery < 34 gestation weeks with PE
**LOPE**: late-onset pre-eclampsia - delivery ≥ 34 gestation weeks with PE
**Preterm PE:** delivery < 37 gestation weeks with PE
**Term PE:** delivery ≥ 37 gestation weeks with PE
**Intermediate PE:** delivery between 34 to 37 gestation weeks with PE
**Late PE:** delivery ≥ 38 gestation weeks with PE

**Multiple countries (1): UK, New Zealand, Australia, Ireland, USA, Greece, Argentina, Colombia, India, Italy, Kenya, Peru, Switzerland, Thailand, Netherlands, Norway, Denmark**
**Multiple countries (2): UK, Ireland, Australia, New Zealand**

**# Model developed using data from first trimester**
**$ Model developed using data from second trimester**

Additional file 1: Table S3 Existing external validations for eligible models

| Outcome | Model | External validation study | Validation cohort size, N (cases) [median] ¥ | Estimated sample size (cases) required for existing validation study, N # | Validation cohort source | Discrimination reported | Calibration reported |
|---|---|---|---|---|---|---|---|
| Gestational diabetes (GDM) | van Leeuwen 2010 [1] | Kotzaeridi 2021 [31] | 1,132 (239) | 3,035 (641) | Austria | Yes | Yes |
| | | Meertens 2020 [32] | 5,260 (127) | 16,378 (396) | Netherlands | Yes | Yes |
| | | Sweeting 2017 [2] | 980 (248) | 2,750 (696) | Australia | Yes | No |
| | | Thériault 2014 [33] | 5,302 (280) | 8,293 (438) | Canada | Yes | No |
| | | van Hoorn 2021 [34] | 3,723 (181) | 8,888 (433) | Netherlands | Yes | Yes |
| | Sweeting 2017 [2] | Kotzaeridi 2021 [31] | 1,132 (239) | 1,961 (415) | Austria | Yes | Yes |
| | | Meertens 2020 [32] | 5,260 (127) | 15,540 (376) | Netherlands | Yes | Yes |
| | Benhalima 2020 [3] | Kotzaeridi 2021 [31] | 1,132 (239) | 4,267 (901) | Austria | Yes | Yes |
| Stillbirth | Yerlikaya 2016 for stillbirth ≥ 24 weeks [26] | Allotey 2022 [35] | 812 - 379,390 (3 - 1,792) [2,540] | 309,950 (1,465) | Multiple countries (1) | Yes | Yes |
| | Trudell 2017 for stillbirth ≥ 32 weeks [27] | Allotey 2022 [35] | 425 - 379,390 (2 - 895) [3,019] | 480,860 (1,135) | Multiple countries (2) | Yes | Yes |
| Pre-eclampsia (PE) | Plasencia 2007 for Any PE [6] | Allotey 2020 [13] | 658 - 1,554 (14 - 56) [1,045] | 10,285 (371) | UK | Yes | Yes |
| | | Lamain-de Ruiter 2019 [36] | 3,736 (87) | 16,126 (376) | Netherlands | Yes | Yes |
| | | Meertens 2019 [37] | 2,614 (76) | 12,840 (374) | Netherlands | Yes | Yes |
| | | Snell 2020 [38] | 658 - 1,554 (14 - 56) [1,045] | 10,285 (371) | UK | Yes | Yes |
| | Poon 2008 for Any PE (1) [8] | Meertens 2019 [37] | 2,614 (76) | 12,840 (374) | Netherlands | Yes | Yes |
| | Poon 2008 for Any PE (2) [9] | Allotey 2020 [13] | 658 - 1,554 (14 - 56) [1,045] | 10,285 (371) | UK | Yes | Yes |
| | | Allen 2017 [39] | 2,180 (52) | 15,734 (376) | UK | Yes | Yes |
| | | Meertens 2019 [37] | 2,614 (76) | 12,840 (374) | Netherlands | Yes | Yes |
| | | Snell 2020 [38] | 658 - 1,554 (14 - 56) [1,045] | 10,285 (371) | UK | Yes | Yes |
| | Direkvand-Moghadam 2013 for Any PE [10] | Meertens 2019 [37] | 2,614 (76) | 36,145 (1,051) | Netherlands | Yes | Yes |

| | | | | | | | |
|---|---|---|---|---|---|---|---|
| | Macdonald-Wallis 2015 for Any PE [11] | Meertens 2019 [37] | 2,614 (76) | 13,869 (404) | Netherlands | Yes | Yes |
| | Allotey 2020 for Any PE (a) [13] | Allotey 2020 [13] | 230 - 7,273 (14 - 278) [1,704] | 48,934 (949) | Multiple countries (3) | Yes | Yes |
| | Allotey 2020 for Any PE (b) [13] | Allotey 2020 [13] | 230 - 7,273 (32 - 278) [3,097] | 36,687 (712) | Multiple countries (4) | Yes | Yes |
| | Plasencia 2007 for EOPE [6] | Allotey 2020 [13] | 316 - 4,212 (6 - 10) [1,106] | 161,554 (384) | UK | Yes | Yes |
| | | Snell 2020 [38] | 316 - 4,212 (6 - 10) [1,106] | 161,554 (384) | UK | Yes | Yes |
| | Kuc 2013 for EOPE [16] | Allotey 2020 [13] | 658 - 136,635 (6 - 1,237) [9,278] | Not available | UK | Yes | Yes |
| | | Snell 2020 [38] | 658 - 136,635 (6 - 1,237) [9,278] | Not available | UK | Yes | Yes |
| | Scazzocchio 2013 for EOPE [17] | Allotey 2020 [13] | 658 - 4,212 (6 - 10) [1,554] | Not available | UK | Yes | Yes |
| | | Snell 2020 [38] | 658 - 4,212 (6 - 10) [1,554] | Not available | UK | Yes | Yes |
| | Baschat 2014 for EOPE [18] | Allotey 2020 [13] | 658 - 14,344 (4 - 144) [2,422] | 137,486 (384) | UK | Yes | Yes |
| | | Snell 2020 [38] | 658 - 14,344 (4 - 144) [2,422] | 137,486 (384) | UK | Yes | Yes |
| | | Allen 2017 [39] | 2,186 (52) | 15,778 (376) | UK | Yes | No |
| | Crovetto 2015 for EOPE [20] | Allotey 2020 [13] | 658 - 4,212 (6 - 10) [1,554] | 168,429 (400) | UK | Yes | Yes |
| | | Snell 2020 [38] | 658 - 4,212 (6 - 10) [1,554] | 168,429 (400) | UK | Yes | Yes |
| | Allotey 2020 for EOPE (a) [13] | Allotey 2020 [13] | 230 - 7,273 (1 - 44) [1,704] | 232,508 (576) | Multiple countries (3) | Yes | Yes |
| | Allotey 2020 for EOPE (b) [13] | Allotey 2020 [13] | 230 - 7,273 (1 - 44) [3,097] | 223,310 (553) | Multiple countries (4) | Yes | Yes |
| | Plasencia 2007 for LOPE [6] | Allotey 2020 [13] | 658 - 1,554 (13 - 51) [1,045] | 11,331 (372) | UK | Yes | Yes |
| | | Lamain-de Ruiter 2019 [36] | 3,736 (71) | 19,847 (378) | Netherlands | Yes | Yes |
| | | Snell 2020 [38] | 658 - 1,554 (13 - 51) [1,045] | 11,331 (372) | UK | Yes | Yes |
| | Poon 2010 for LOPE [21] | Allotey 2020 [13] | 658 - 1,554 (13 - 51) [1,045] | 11,331 (372) | UK | Yes | Yes |
| | | Lamain-de Ruiter 2019 [36] | 3,736 (71) | 19,847 (378) | Netherlands | Yes | Yes |
| | | Snell 2020 [38] | 658 - 1,554 (13 - 51) [1,045] | 11,331 (372) | UK | Yes | Yes |
| | Kuc 2013 for LOPE [16] | Allotey 2020 [13] | 449 - 136,635 (13 - 3,733) [2,883] | Not available | UK | Yes | Yes |
| | | Lamain-de Ruiter 2019 [36] | 3,736 (71) | Not available | Netherlands | Yes | Yes |

| | | | | | | | |
|---|---|---|---|---|---|---|---|
| | | Snell 2020 [38] | 449 - 136,635 (13 - 3,733) [2,883] | Not available | UK | Yes | Yes |
| | Scazzocchio 2013 for LOPE [17] | Allotey 2020 [13] | 658 (26) | Not available | UK | Yes | Yes |
| | | Lamain-de Ruiter 2019 [36] | 3,736 (71) | Not available | Netherlands | Yes | Yes |
| | | Snell 2020 [38] | 658 (26) | Not available | UK | Yes | Yes |
| | Crovetto 2015 for LOPE [20] | Allotey 2020 [13] | 316 - 4,212 (13 - 263) [1,045] | 10,528 (658) | UK | Yes | Yes |
| | | Lamain-de Ruiter 2019 [36] | 3,736 (71) | 30,932 (588) | Netherlands | Yes | Yes |
| | | Snell 2020 [38] | 316 - 4,212 (13 - 263) [1,045] | 10,528 (658) | UK | Yes | Yes |
| | Allotey 2020 for LOPE (a) [13] | Allotey 2020 [13] | 230 - 7,273 (13 - 263) [1,704] | 55,809 (944) | Multiple countries (3) | Yes | Yes |
| | Allotey 2020 for LOPE (b) [13] | Allotey 2020 [13] | 230 - 7,273 (19 - 263) [3,097] | 40,933 (693) | Multiple countries (4) | Yes | Yes |
| Small-for-gestational-age (SGA) | Macdonald-Wallis 2015 [11] | Meertens 2019 [40] | 2,582 (203) | 10,625 (836) | Netherlands | Yes | Yes |
| | González-González 2017 [28] | Meertens 2019 [40] | 2,582 (203) | 31,874 (2,506) | Netherlands | Yes | Yes |
| | Poon 2011 (log(birthweight)) [30] | Allotey 2024 [41] | 823 - 406,286 [1,877] | Not applicable | Multiple countries (5) | Not applicable | Yes |

**Note:**

**Multiple countries (1):** UK, USA, Japan, and Norway

**Multiple countries (2):** UK, USA, Japan, Norway, Denmark, Indonesia, Netherlands, New Zealand, Australia, Ireland, Greece, Argentina, Colombia, India, Italy, Kenya, Peru, Switzerland, and Thailand

**Multiple countries (3):** UK, New Zealand, Australia, Ireland, USA, Greece, Argentina, Colombia, India, Italy, Kenya, Peru, Switzerland, Thailand, Netherlands, Norway, and Denmark

**Multiple countries (4):** UK, New Zealand, Australia, Ireland, USA, Greece, Argentina, Colombia, India, Italy, Kenya, Peru, Switzerland, Thailand, Netherlands, and Norway

**Multiple countries (5):** UK, USA, Netherlands, Australia, Japan, and Norway

**EOPE:** early-onset pre-eclampsia - delivery < 34 gestation weeks with PE
**LOPE**: late-onset pre-eclampsia - delivery ≥ 34 gestation weeks with PE

**¥** If multiple cohorts used: reported as min-max N (min-max cases) [median N].

**#** Calculated from each external validation study's cohort outcome prevalence and the C-statistic reported in the original model development study. If multiple cohorts were used, the largest cohort was evaluated.

Additional file 1: Table S4 Characteristics of predictors in primary analysis

| Variable | | Full cohort (Pregnancies, N = 1,581,161) | | Cohort for Small-for-gestational-age (Pregnancies, N = 957,496) | |
|---|---|---|---|---|---|
| | | Mean (SD) / Count (%) # | Missing, N (%) # | Mean (SD) / Count (%) # | Missing, N (%) # |
| **Diastolic blood pressure (mmHg)** | | 70.70 (9.36) | 491,331 (31.07%) | 70.62 (9.34) | 278,443 (29.08%) |
| **Systolic blood pressure (mmHg)** | | 114.64 (12.57) | 491,336 (31.07%) | 114.54 (12.51) | 278,462 (29.08%) |
| **Baby sex** | **Male** | 489,302 (30.95%) | 626,127 (39.60%) | 472,751 (49.37%) | 32,143 (3.36%) |
| | **Female** | 465,732 (29.46%) | | 452,602 (47.27%) | |
| **Antiphospholipid syndrome** | | 1,125 (0.07%) | 0 | 687 (0.07%) | 0 |
| **Assisted conception** | | 6,338 (0.40%) | 0 | 2,830 (0.30%) | 0 |
| **Pre-existing diabetes** | | 13,099 (0.83%) | 0 | 7,493 (0.78%) | 0 |
| **Family history of diabetes** | | 224,383 (14.19%) | 0 | 143,027 (14.94%) | 0 |
| **Family history of hypertension** | | 159,302 (10.08%) | 0 | 98,147 (10.25%) | 0 |
| **Family history of pre-eclampsia** | | 121 (0.01%) | 0 | 87 (0.01%) | 0 |
| **Hypertension** | | 18,501 (1.17%) | 0 | 10,944 (1.14%) | 0 |
| **Infertility** | | 93,912 (5.94%) | 0 | 54,202 (5.66%) | 0 |
| **Previous fetal macrosomia** | | 25,687 (1.62%) | 0 | 16,817 (1.76%) | 0 |
| **Previous small-for-gestational-age** | | 55,414 (3.50%) | 0 | 38,796 (4.05%) | 0 |
| **Previous gestational diabetes** | | 13,900 (0.88%) | 0 | 8,782 (0.92%) | 0 |
| **Previous gestational hypertension** | | 34,444 (2.18%) | 0 | 21,424 (2.24%) | 0 |
| **Previous pre-eclampsia** | | 14,416 (0.91%) | 0 | 9,079 (0.95%) | 0 |
| **Previous stillbirth** | | 6,609 (0.42%) | 0 | 3,819 (0.40%) | 0 |
| **Smoking** | | 230,651 (14.59%) | 0 | 143,086 (14.94%) | 0 |
| **Systemic lupus erythematosus** | | 1,308 (0.08%) | 0 | 799 (0.08%) | 0 |
| **Renal disease** | | 2,621 (0.17%) | 0 | 1,577 (0.16%) | 0 |
| **Thrombophilia** | | 2,774 (0.18%) | 0 | 1,701 (0.18%) | 0 |

**Note:**

**SD:** standard deviation
### Percentages may not sum to 100% due to rounding.

Additional file 1: Table S5 Validation results of primary analysis

| Outcome | Model | C-Statistic | Calibration-in-the-large | Calibration slope |
|---|---|---|---|---|
| Gestational diabetes (GDM) | van Leeuwen 2010 [1] | 0.74 (0.73 - 0.74) | -0.44 (-0.45 - -0.44) | 1.26 (1.25 - 1.27) |
| | Sweeting 2017 [2] | 0.74 (0.74 - 0.74) | -1.88 (-1.89 - -1.87) | 0.48 (0.48 - 0.49) |
| | Benhalima 2020 [3] | 0.77 (0.77 - 0.77) | -0.99 (-0.99 - -0.98) | 1.09 (1.08 - 1.10) |
| | Basil 2024 [4] | 0.71 (0.71 - 0.72) | -1.43 (-1.43 - -1.42) | 0.69 (0.68 - 0.69) |
| | Syngelaki 2025 (for nulliparous) [5] | 0.75 (0.75 - 0.76) | -0.78 (-0.79 - -0.76) | 1.07 (1.05 - 1.08) |
| | Syngelaki 2025 (for parous without previous GDM) [5] | 0.76 (0.75 - 0.76) | -0.81 (-0.82 - -0.80) | 1.04 (1.03 - 1.06) |
| | Syngelaki 2025 (for parous with previous GDM) [5] | 0.57 (0.56 - 0.58) | 0.20 (0.17 - 0.24) | 0.49 (0.42 - 0.56) |
| Pre-eclampsia (PE) | Plasencia 2007 for Any PE [6] | 0.69 (0.66 - 0.72) | 0.58 (0.57 - 0.59) | 0.63 (0.53 - 0.74) |
| | Emonts 2008 for Any PE [7] | 0.57 (0.54 - 0.61) | -5.52 (-5.54 - -5.51) | 0.19 (0.10 - 0.27) |
| | Poon 2008 for Any PE (1) [8] | 0.67 (0.65 - 0.69) | 0.75 (0.74 - 0.76) | 0.46 (0.41 - 0.52) |
| | Poon 2008 for Any PE (2) [9] | 0.69 (0.66 - 0.72) | 0.46 (0.44 - 0.47) | 0.67 (0.55 - 0.78) |
| | Direkvand-Moghadam 2013 for Any PE [10] | 0.55 (0.55 - 0.55) | -3.20 (-3.21 - -3.19) | 0.85 (0.83 - 0.87) |
| | Macdonald-Wallis 2015 for Any PE [11] | 0.72 (0.69 - 0.74) | -1.63 (-1.65 - -1.62) | 0.66 (0.60 - 0.73) |
| | Rocha 2017 for Any PE (a) [12] | 0.61 (0.55 - 0.68) | -3.83 (-3.84 - -3.81) | 5.54 (3.05 - 8.03) |
| | Rocha 2017 for Any PE (b) [12] | 0.67 (0.63 - 0.71) | -3.80 (-3.81 - -3.79) | 9.58 (7.58 - 11.58) |
| | Allotey 2020 for Any PE (a) [13] | 0.72 (0.70 - 0.74) | -0.19 (-0.20 - -0.18) | 1.10 (1.01 - 1.20) |
| | Allotey 2020 for Any PE (b) [13] | 0.72 (0.69 - 0.75) | -0.18 (-0.19 - -0.17) | 0.92 (0.82 - 1.02) |
| | Tang 2022 for Any PE [14] | 0.69 (0.66 - 0.73) | 0.92 (0.90 - 0.93) | 0.63 (0.51 - 0.75) |
| | Tarca 2022 for Any PE [15] | 0.70 (0.70 - 0.71) | -0.68 (-0.69 - -0.66) | 1.48 (1.46 - 1.50) |
| | Plasencia 2007 for EOPE [6] | 0.56 (0.56 - 0.57) | -0.13 (-0.16 - -0.09) | 0.64 (0.60 - 0.68) |
| | Kuc 2013 for EOPE [16] | 0.65 (0.64 - 0.66) | -5.14 (-5.17 - -5.10) | 0.34 (0.32 - 0.37) |
| | Scazzocchio 2013 for EOPE [17] | 0.64 (0.63 - 0.66) | -1.15 (-1.19 - -1.12) | 0.63 (0.61 - 0.66) |
| | Baschat 2014 for EOPE [18] | 0.69 (0.68 - 0.70) | -1.20 (-1.24 - -1.17) | 2.94 (2.84 - 3.04) |
| | Teixeira 2014 for EOPE [19] | 0.69 (0.68 - 0.70) | -1.23 (-1.26 - -1.19) | 0.68 (0.65 - 0.71) |
| | Crovetto 2015 for EOPE [20] | 0.50 (0.49 - 0.51) | -1.65 (-1.69 - -1.62) | 0.36 (0.34 - 0.39) |
| | Allotey 2020 for EOPE (a) [13] | 0.67 (0.66 - 0.68) | -0.74 (-0.77 - -0.70) | 0.82 (0.79 - 0.86) |
| | Allotey 2020 for EOPE (b) [13] | 0.67 (0.66 - 0.69) | -0.88 (-0.92 - -0.85) | 0.86 (0.83 - 0.90) |
| | Plasencia 2007 for LOPE [6] | 0.68 (0.65 - 0.72) | 0.68 (0.67 - 0.70) | 0.59 (0.47 - 0.71) |
| | Poon 2010 for LOPE [21] | 0.68 (0.64 - 0.72) | 0.66 (0.65 - 0.68) | 0.63 (0.49 - 0.76) |
| | Kuc 2013 for LOPE [16] | 0.65 (0.60 - 0.69) | -2.14 (-2.15 - -2.13) | 0.62 (0.40 - 0.84) |
| | Scazzocchio 2013 for LOPE [17] | 0.68 (0.62 - 0.73) | -0.19 (-0.21 - -0.18) | 0.74 (0.59 - 0.89) |

| | | | | |
|---|---|---|---|---|
| | Crovetto 2015 for LOPE [20] | 0.54 (0.49 - 0.59) | -0.59 (-0.60 - -0.58) | 0.29 (0.16 - 0.43) |
| | Allotey 2020 for LOPE (a) [13] | 0.71 (0.69 - 0.73) | -0.09 (-0.10 - -0.08) | 1.04 (0.94 - 1.14) |
| | Allotey 2020 for LOPE (b) [13] | 0.71 (0.68 - 0.75) | -0.01 (-0.03 - -0.00) | 0.86 (0.74 - 0.97) |
| | Zhang 2025 for LOPE [22] | 0.68 (0.64 - 0.73) | -2.57 (-2.60 - -2.55) | 0.32 (0.25 - 0.38) |
| | Myers 2013 for Preterm PE [23] | 0.66 (0.65 - 0.67) | -6.14 (-6.16 - -6.12) | 0.61 (0.59 - 0.64) |
| | Rocha 2017 for Preterm PE (a) [12] | 0.63 (0.62 - 0.64) | -5.30 (-5.32 - -5.27) | 16.47 (16.01 - 16.92) |
| | Rocha 2017 for Preterm PE (b) [12] | 0.68 (0.67 - 0.69) | -5.33 (-5.35 - -5.31) | 17.78 (17.35 - 18.21) |
| | Sepúlveda-Martínez 2019 for Preterm PE [24] | 0.62 (0.61 - 0.62) | -0.71 (-0.74 - -0.69) | 0.81 (0.78 - 0.83) |
| | Rocha 2017 for Term PE [12] | 0.66 (0.59 - 0.73) | -4.10 (-4.12 - -4.09) | 10.58 (6.42 - 14.73) |
| | Lai 2013 for Intermediate PE [25] | 0.68 (0.65 - 0.70) | -0.01 (-0.03 - 0.02) | 0.56 (0.49 - 0.63) |
| | Lai 2013 for Late PE [25] | 0.68 (0.68 - 0.68) | -0.03 (-0.05 - -0.02) | 0.70 (0.69 - 0.72) |
| Stillbirth | Yerlikaya 2016 for stillbirth ≥ 24 weeks [26] | 0.58 (0.57 - 0.58) | 0.49 (0.46 - 0.51) | 0.87 (0.83 - 0.91) |
| | Trudell 2017 for stillbirth ≥ 32 weeks [27] | 0.54 (0.53 - 0.55) | 0.89 (0.86 - 0.92) | 0.42 (0.36 - 0.48) |
| Small-for-gestational-age (SGA) | Macdonald-Wallis 2015 for SGA [11] | 0.65 (0.65 - 0.66) | 9.64 (9.63 - 9.64) | 0.69 (0.68 - 0.70) |
| | González-González 2017 for SGA [28] | 0.58 (0.58 - 0.58) | -0.41 (-0.42 - -0.41) | 0.69 (0.67 - 0.71) |
| | Adjahou 2025 for SGA [29] | 0.67 (0.67 - 0.67) | -0.16 (-0.16 - -0.15) | 0.73 (0.72 - 0.73) |
| | Poon 2011 (log(birthweight)) [30] | Not applicable | 104.23 g (103.46 - 105.01) | 0.94 (0.94 - 0.95) |

**Note:**

**PE:** pre-eclampsia
**EOPE:** early-onset pre-eclampsia - delivery < 34 gestation weeks with PE
**LOPE**: late-onset pre-eclampsia - delivery ≥ 34 gestation weeks with PE
**Preterm PE:** delivery < 37 gestation weeks with PE
**Term PE:** delivery ≥ 37 gestation weeks with PE
**Intermediate PE:** delivery between 34 to 37 gestation weeks with PE
**Late PE:** delivery ≥ 38 gestation weeks with PE

Additional file 1: Table S6 Characteristics of the validation cohort when selecting the first of overlapping pregnancies

| | | Full cohort (Pregnancies, N = 1,527,385) | | Sub-cohort for small-for-gestational-age (Pregnancies, N = 936,994) | |
|---|---|---|---|---|---|
| **Variable** | | Mean (SD) / Count (%) # | Missing, N (%) # | Mean (SD) / Count (%) # | Missing, N (%) # |
| **Maternal age (years)** | | 29.86 (5.68) | 0 | 29.77 (5.63) | 0 |
| **Gravidity (total pregnancies)** | | 2.36 (1.54) | 0 | 2.34 (1.51) | 0 |
| **Parity** | Nulliparous | 694,653 (45.48%) | 0 | 428,362 (45.72%) | 0 |
| | Parous | 832,732 (54.52%) | 0 | 508,632 (54.28%) | 0 |
| **Gestational length (weeks)** | | 39.05 (2.64) | 0 | 39.19 (1.89) | 0 |
| **Height (cm)** | | 163.93 (6.94) | 486,683 (31.86%) | 163.89 (6.91) | 280,765 (29.96%) |
| **Weight (kg)** | | 68.82 (16.05) | 310,245 (20.31%) | 68.95 (16.11) | 172,273 (18.39%) |
| **Body Mass Index (kg/m²)** | | 25.55 (5.74) | 419,990 (27.50%) | 25.62 (5.76) | 236,173 (25.21%) |
| **Ethnicity** | **Bangladeshi** | 21,345 (1.40%) | 22,188 (1.45%) | 14,971 (1.60%) | 10,552 (1.13%) |
| | **Black African** | 46,931 (3.07%) | | 29,131 (3.11%) | |
| | **Black Caribbean** | 15,042 (0.98%) | | 9,074 (0.97%) | |
| | **Black Other** | 12,733 (0.83%) | | 7,961 (0.85%) | |
| | **Chinese** | 9,162 (0.60%) | | 5,874 (0.63%) | |
| | **Indian** | 45,018 (2.95%) | | 28,102 (3.00%) | |
| | **Mixed** | 21,833 (1.43%) | | 13,914 (1.48%) | |
| | **Other** | 38,804 (2.54%) | | 24,338 (2.60%) | |
| | **Other Asian** | 31,030 (2.03%) | | 19,656 (2.10%) | |
| | **Pakistani** | 50,682 (3.32%) | | 33,242 (3.55%) | |
| | **White** | 1,212,617 (79.39%) | | 740,179 (79.00%) | |
| **Gestational diabetes** | | 62,947 (4.12%) | 0 | 43,803 (4.67%) | 0 |
| **Pre-eclampsia (Any)** | | 33,929 (2.22%) | 0 | 22,714 (2.42%) | 0 |
| **Pre-eclampsia (early-onset)** | | 3,186 (0.21%) | 0 | 2,357 (0.25%) | 0 |
| **Pre-eclampsia (late-onset)** | | 30,743 (2.01%) | 0 | 20,357 (2.17%) | 0 |
| **Pre-eclampsia (preterm)** | | 7,870 (0.52%) | 0 | 6,073 (0.65%) | 0 |
| **Pre-eclampsia (term)** | | 26,059 (1.71%) | 0 | 16,641 (1.78%) | 0 |
| **Pre-eclampsia (intermediate)** | | 8,251 (0.54%) | 0 | 6,792 (0.72%) | 0 |
| **Pre-eclampsia (late)** | | 22,492 (1.47%) | 0 | 13,565 (1.45%) | 0 |
| **Stillbirth (≥24 weeks)** | | 7,367 (0.48%) | 0 | 3,648 (0.39%) | 0 |
| **Stillbirth (≥32 weeks)** | | 5,582 (0.37%) | 0 | 2,414 (0.26%) | 0 |
| **Birthweight (g)** | | 3,376.99 (542.15) | 590,391 (38.65%) | 3,376.99 (542.15) | 0 |
| **Small-for-gestational-age** | | / | / | 126,143 (13.46%) | 0 |
| **Diastolic blood pressure (mmHg)** | | 70.67 (9.36) | 459,025 (30.05%) | 70.59 (9.34) | 266,705 (28.46%) |
| **Systolic blood pressure (mmHg)** | | 114.61 (12.56) | 459,054 (30.05%) | 114.52 (12.51) | 266,739 (28.47%) |
| **Baby sex** | Male | 478,738 (31.34%) | 592,848 (38.81%) | 462,632 (49.37%) | 31,419 (3.35%) |
| | Female | 455,799 (29.84%) | | 442,943 (47.27%) | |
| **Antiphospholipid syndrome** | | 1,090 (0.07%) | 0 | 676 (0.07%) | 0 |
| **Assisted conception** | | 6,174 (0.40%) | 0 | 2,780 (0.30%) | 0 |
| **Pre-existing diabetes** | | 12,646 (0.83%) | 0 | 7,349 (0.78%) | 0 |
| **Family history of diabetes** | | 217,631 (14.25%) | 0 | 140,231 (14.97%) | 0 |
| **Family history of hypertension** | | 153,436 (10.05%) | 0 | 95,922 (10.24%) | 0 |

| | | | | |
|---|---|---|---|---|
| **Family history of pre-eclampsia** | 119 (0.01%) | 0 | 86 (0.01%) | 0 |
| **Hypertension** | 17,776 (1.16%) | 0 | 10,715 (1.14%) | 0 |
| **Infertility** | 90,642 (5.93%) | 0 | 53,001 (5.66%) | 0 |
| **Previous fetal macrosomia** | 24,358 (1.59%) | 0 | 16,486 (1.76%) | 0 |
| **Previous small-for-gestational-age** | 52,865 (3.46%) | 0 | 38,069 (4.06%) | 0 |
| **Previous gestational diabetes** | 12,718 (0.83%) | 0 | 8,327 (0.89%) | 0 |
| **Previous gestational hypertension** | 32,388 (2.12%) | 0 | 20,804 (2.22%) | 0 |
| **Previous pre-eclampsia** | 13,511 (0.88%) | 0 | 8,824 (0.94%) | 0 |
| **Previous stillbirth** | 6,014 (0.39%) | 0 | 3,660 (0.39%) | 0 |
| **Smoking** | 222,745 (14.58%) | 0 | 139,999 (14.94%) | 0 |
| **Systemic lupus erythematosus** | 1,253 (0.08%) | 0 | 780 (0.08%) | 0 |
| **Renal disease** | 2,511 (0.16%) | 0 | 1,526 (0.16%) | 0 |
| **Thrombophilia** | 2,686 (0.18%) | 0 | 1,671 (0.18%) | 0 |

**Note:**

**SD:** standard deviation
**#** Percentages may not sum to 100% due to rounding.

Additional file 1: Table S7 Validation results of the non-overlapping analysis

| Outcome | Model | C-Statistic | Calibration-in-the-large | Calibration slope |
|---|---|---|---|---|
| Gestational diabetes (GDM) | van Leeuwen 2010 [1] | 0.73 (0.73 - 0.73) | -0.44 (-0.45 - -0.44) | 1.24 (1.22 - 1.25) |
| | Sweeting 2017 [2] | 0.74 (0.74 - 0.74) | -1.87 (-1.88 - -1.86) | 0.48 (0.47 - 0.48) |
| | Benhalima 2020 [3] | 0.77 (0.77 - 0.77) | -0.97 (-0.98 - -0.97) | 1.08 (1.07 - 1.09) |
| | Basil 2024 [4] | 0.71 (0.71 - 0.72) | -1.42 (-1.42 - -1.41) | 0.68 (0.68 - 0.69) |
| | Syngelaki 2025 (for nulliparous) [5] | 0.75 (0.75 - 0.76) | -0.78 (-0.79 - -0.76) | 1.07 (1.05 - 1.08) |
| | Syngelaki 2025 (for parous without previous GDM) [5] | 0.75 (0.75 - 0.76) | -0.77 (-0.79 - -0.76) | 1.04 (1.03 - 1.06) |
| | Syngelaki 2025 (for parous with previous GDM) [5] | 0.58 (0.57 - 0.59) | 0.14 (0.10 - 0.18) | 0.53 (0.46 - 0.61) |
| Pre-eclampsia (PE) | Plasencia 2007 for Any PE [6] | 0.69 (0.65 - 0.72) | 0.57 (0.56 - 0.58) | 0.63 (0.53 - 0.73) |
| | Emonts 2008 for Any PE [7] | 0.57 (0.53 - 0.61) | -5.53 (-5.54 - -5.51) | 0.18 (0.10 - 0.27) |
| | Poon 2008 for Any PE (1) [8] | 0.67 (0.65 - 0.69) | 0.73 (0.72 - 0.74) | 0.46 (0.40 - 0.52) |
| | Poon 2008 for Any PE (2) [9] | 0.69 (0.65 - 0.72) | 0.45 (0.43 - 0.46) | 0.66 (0.55 - 0.78) |
| | Direkvand-Moghadam 2013 for Any PE [10] | 0.55 (0.54 - 0.55) | -3.19 (-3.20 - -3.18) | 0.82 (0.80 - 0.84) |
| | Macdonald-Wallis 2015 for Any PE [11] | 0.72 (0.70 - 0.74) | -1.64 (-1.66 - -1.63) | 0.67 (0.60 - 0.74) |
| | Rocha 2017 for Any PE (a) [12] | 0.61 (0.55 - 0.67) | -3.82 (-3.83 - -3.81) | 5.49 (3.03 - 7.95) |
| | Rocha 2017 for Any PE (b) [12] | 0.67 (0.63 - 0.71) | -3.80 (-3.81 - -3.79) | 9.52 (7.55 - 11.49) |
| | Allotey 2020 for Any PE (a) [13] | 0.72 (0.69 - 0.74) | -0.20 (-0.21 - -0.19) | 1.10 (1.01 - 1.20) |
| | Allotey 2020 for Any PE (b) [13] | 0.72 (0.69 - 0.75) | -0.18 (-0.20 - -0.17) | 0.92 (0.82 - 1.02) |
| | Tang 2022 for Any PE [14] | 0.69 (0.66 - 0.73) | 0.91 (0.90 - 0.93) | 0.63 (0.52 - 0.75) |
| | Tarca 2022 for Any PE [15] | 0.70 (0.70 - 0.71) | -0.67 (-0.68 - -0.66) | 1.48 (1.46 - 1.50) |
| | Plasencia 2007 for EOPE [6] | 0.56 (0.55 - 0.57) | -0.12 (-0.16 - -0.09) | 0.61 (0.57 - 0.65) |
| | Kuc 2013 for EOPE [16] | 0.65 (0.64 - 0.66) | -5.15 (-5.19 - -5.12) | 0.35 (0.33 - 0.37) |
| | Scazzocchio 2013 for EOPE [17] | 0.64 (0.63 - 0.65) | -1.14 (-1.18 - -1.10) | 0.63 (0.60 - 0.65) |
| | Baschat 2014 for EOPE [18] | 0.69 (0.67 - 0.70) | -1.20 (-1.23 - -1.16) | 2.94 (2.84 - 3.04) |
| | Teixeira 2014 for EOPE [19] | 0.69 (0.68 - 0.70) | -1.23 (-1.27 - -1.20) | 0.67 (0.64 - 0.70) |
| | Crovetto 2015 for EOPE [20] | 0.50 (0.49 - 0.51) | -1.63 (-1.66 - -1.59) | 0.34 (0.32 - 0.37) |
| | Allotey 2020 for EOPE (a) [13] | 0.67 (0.65 - 0.68) | -0.73 (-0.76 - -0.69) | 0.81 (0.78 - 0.85) |
| | Allotey 2020 for EOPE (b) [13] | 0.67 (0.66 - 0.68) | -0.87 (-0.91 - -0.84) | 0.85 (0.81 - 0.88) |
| | Plasencia 2007 for LOPE [6] | 0.68 (0.65 - 0.72) | 0.67 (0.66 - 0.68) | 0.60 (0.48 - 0.71) |
| | Poon 2010 for LOPE [21] | 0.68 (0.64 - 0.72) | 0.66 (0.64 - 0.67) | 0.63 (0.49 - 0.76) |
| | Kuc 2013 for LOPE [16] | 0.65 (0.60 - 0.70) | -2.15 (-2.16 - -2.14) | 0.63 (0.40 - 0.85) |
| | Scazzocchio 2013 for LOPE [17] | 0.68 (0.62 - 0.73) | -0.19 (-0.20 - -0.18) | 0.74 (0.59 - 0.88) |

| | | | | |
|---|---|---|---|---|
| | Crovetto 2015 for LOPE [20] | 0.54 (0.49 - 0.59) | -0.57 (-0.58 - -0.55) | 0.28 (0.15 - 0.42) |
| | Allotey 2020 for LOPE (a) [13] | 0.71 (0.69 - 0.73) | -0.09 (-0.11 - -0.08) | 1.04 (0.94 - 1.14) |
| | Allotey 2020 for LOPE (b) [13] | 0.71 (0.68 - 0.75) | -0.02 (-0.03 - -0.01) | 0.86 (0.74 - 0.97) |
| | Zhang 2025 for LOPE [22] | 0.68 (0.64 - 0.73) | -2.57 (-2.59 - -2.55) | 0.32 (0.25 - 0.38) |
| | Myers 2013 for Preterm PE [23] | 0.66 (0.65 - 0.67) | -6.13 (-6.15 - -6.11) | 0.61 (0.59 - 0.64) |
| | Rocha 2017 for Preterm PE (a) [12] | 0.63 (0.62 - 0.64) | -5.29 (-5.31 - -5.27) | 16.29 (15.83 - 16.76) |
| | Rocha 2017 for Preterm PE (b) [12] | 0.68 (0.67 - 0.68) | -5.32 (-5.35 - -5.30) | 17.65 (17.21 - 18.09) |
| | Sepúlveda-Martínez 2019 for Preterm PE [24] | 0.61 (0.61 - 0.62) | -0.71 (-0.73 - -0.68) | 0.79 (0.77 - 0.82) |
| | Rocha 2017 for Term PE [12] | 0.66 (0.59 - 0.73) | -4.10 (-4.11 - -4.08) | 10.61 (6.52 - 14.69) |
| | Lai 2013 for Intermediate PE [25] | 0.67 (0.65 - 0.70) | -0.01 (-0.04 - 0.01) | 0.56 (0.49 - 0.63) |
| | Lai 2013 for Late PE [25] | 0.68 (0.65 - 0.70) | -0.04 (-0.05 - -0.02) | 0.71 (0.69 - 0.72) |
| Stillbirth | Yerlikaya 2016 for stillbirth ≥ 24 weeks [26] | 0.56 (0.56 - 0.57) | 0.47 (0.46 - 0.49) | 0.71 (0.66 - 0.76) |
| | Trudell 2017 for stillbirth ≥ 32 weeks [27] | 0.54 (0.53 - 0.55) | 0.86 (0.83 - 0.89) | 0.44 (0.38 - 0.50) |
| Small-for-gestational-age (SGA) | Macdonald-Wallis 2015 for SGA [11] | 0.65 (0.65 - 0.66) | 9.63 (9.62 - 9.64) | 0.69 (0.68 - 0.70) |
| | González-González 2017 for SGA [28] | 0.58 (0.58 - 0.58) | -0.41 (-0.42 - -0.41) | 0.69 (0.68 - 0.71) |
| | Adjahou 2025 for SGA [29] | 0.67 (0.67 - 0.67) | -0.17 (-0.17 - -0.16) | 0.73 (0.72 - 0.74) |
| | Poon 2011 (log(birthweight)) [30] | Not applicable | 105.25 g (104.46 - 106.04) | 0.95 (0.94 - 0.95) |

**Note:**

**PE:** pre-eclampsia
**EOPE:** early-onset pre-eclampsia - delivery < 34 gestation weeks with PE
**LOPE**: late-onset pre-eclampsia - delivery ≥ 34 gestation weeks with PE
**Preterm PE:** delivery < 37 gestation weeks with PE
**Term PE:** delivery ≥ 37 gestation weeks with PE
**Intermediate PE:** delivery between 34 to 37 gestation weeks with PE
**Late PE:** delivery ≥ 38 gestation weeks with PE

Additional file 1: Table S8 Characteristics of the complete-case validation cohort

| Variable | | Gestational diabetes<br>N = 793,031<br>Mean (SD) / Count (%) # | Pre-eclampsia<br>N = 792,401<br>Mean (SD) / Count (%) # | Stillbirth<br>N = 1,178,044<br>Mean (SD) / Count (%) # | Small-for-gestational-age<br>N = 502,967<br>Mean (SD) / Count (%) # |
|---|---|---|---|---|---|
| **Outcome** | | 36,532 (4.61%) | 18,884 (2.38%) | 5,763 (0.49%) | 66,767 (13.27%) |
| **Maternal age (years)** | | 29.71 (5.54) | 29.71 (5.54) | 29.99 (5.51) | 29.62 (5.50) |
| **Gravidity (total pregnancies)** | | 2.32 (1.50) | 2.32 (1.50) | 2.38 (1.55) | 2.29 (1.48) |
| **Parity** | Nulliparous | 368,111 (46.42%) | 367,831 (46.42%) | 524,954 (44.56%) | 235,417 (46.81%) |
| | Parous | 424,920 (53.58%) | 424,570 (53.58%) | 653,090 (55.44%) | 267,550 (53.19%) |
| **Gestational length (weeks)** | | 38.95 (2.77) | 38.95 (2.77) | 39.00 (2.70) | 39.17 (1.89) |
| **Ethnicity** | **Bangladeshi** | 12,162 (1.53%) | 12,154 (1.53%) | 16,487 (1.40%) | 8,795 (1.75%) |
| | **Black African** | 28,238 (3.56%) | 28,215 (3.56%) | 38,202 (3.24%) | 17,749 (3.53%) |
| | **Black Caribbean** | 8,065 (1.02%) | 8,061 (1.02%) | 11,447 (0.97%) | 5,052 (1.00%) |
| | **Black Other** | 7,201 (0.91%) | 7,197 (0.91%) | 9,892 (0.84%) | 4,614 (0.92%) |
| | **Chinese** | 4,876 (0.61%) | 4,871 (0.61%) | 7,291 (0.62%) | 3,209 (0.64%) |
| | **Indian** | 25,897 (3.27%) | 25,873 (3.27%) | 36,451 (3.09%) | 16,649 (3.31%) |
| | **Mixed** | 11,773 (1.48%) | 11,766 (1.48%) | 16,915 (1.44%) | 7,738 (1.54%) |
| | **Other** | 22,994 (2.90%) | 22,973 (2.90%) | 31,373 (2.66%) | 14,631 (2.91%) |
| | **Other Asian** | 18,513 (2.33%) | 18,502 (2.33%) | 25,329 (2.15%) | 11,969 (2.38%) |
| | **Pakistani** | 26,495 (3.34%) | 26,471 (3.34%) | 37,942 (3.22%) | 17,860 (3.55%) |
| | **White** | 617,211 (77.83%) | 616,717 (77.83%) | 931,266 (79.05%) | 389,828 (77.51%) |
| | **Unknown** | 9,606 (1.21%) | 9,601 (1.21%) | 15,449 (1.31%) | 4,873 (0.97%) |

**Note:**

**SD:** standard deviation
**#** Percentages may not sum to 100% due to rounding.

Additional file 1: Table S9 Validation results of complete-case analysis

| Outcome | Model | C-Statistic | Calibration-in-the-large | Calibration slope |
|---|---|---|---|---|
| Gestational diabetes (GDM) | van Leeuwen 2010 [1] | 0.74 (0.73 - 0.74) | -0.37 (-0.38 - -0.36) | 1.23 (1.21 - 1.24) |
| | Sweeting 2017 [2] | 0.74 (0.73 - 0.74) | -1.96 (-1.98 - -1.95) | 0.45 (0.45 - 0.46) |
| | Benhalima 2020 [3] | 0.77 (0.77 - 0.78) | -0.89 (-0.90 - -0.88) | 1.07 (1.06 - 1.08) |
| | Basil 2024 [4] | 0.71 (0.71 - 0.72) | -1.39 (-1.40 - -1.38) | 0.66 (0.65 - 0.67) |
| | Syngelaki 2025 (for nulliparous) [5] | 0.76 (0.75 - 0.76) | -0.70 (-0.72 - -0.69) | 1.07 (1.05 - 1.09) |
| | Syngelaki 2025 (for parous without previous GDM) [5] | 0.76 (0.76 - 0.76) | -0.73 (-0.75 - -0.72) | 1.04 (1.02 - 1.06) |
| | Syngelaki 2025 (for parous with previous GDM) [5] | 0.56 (0.54 - 0.57) | 0.17 (0.12 - 0.21) | 0.40 (0.30 - 0.49) |
| Pre-eclampsia (PE) | Plasencia 2007 for Any PE [6] | 0.68 (0.68 - 0.69) | 0.61 (0.59 - 0.62) | 0.61 (0.60 - 0.62) |
| | Emonts 2008 for Any PE [7] | 0.57 (0.57 - 0.57) | -5.53 (-5.55 - -5.52) | 0.17 (0.16 - 0.18) |
| | Poon 2008 for Any PE (1) [8] | 0.67 (0.66 - 0.67) | 0.74 (0.73 - 0.76) | 0.44 (0.43 - 0.45) |
| | Poon 2008 for Any PE (2) [9] | 0.68 (0.68 - 0.69) | 0.49 (0.47 - 0.50) | 0.64 (0.63 - 0.65) |
| | Direkvand-Moghadam 2013 for Any PE [10] | 0.55 (0.55 - 0.56) | -3.12 (-3.13 - -3.11) | 0.88 (0.85 - 0.91) |
| | Macdonald-Wallis 2015 for Any PE [11] | 0.71 (0.71 - 0.72) | -1.63 (-1.65 - -1.62) | 0.62 (0.61 - 0.64) |
| | Rocha 2017 for Any PE (a) [12] | 0.61 (0.61 - 0.62) | -3.75 (-3.76 - -3.73) | 5.25 (5.07 - 5.43) |
| | Rocha 2017 for Any PE (b) [12] | 0.67 (0.67 - 0.67) | -3.72 (-3.74 - -3.71) | 9.39 (9.18 - 9.60) |
| | Allotey 2020 for Any PE (a) [13] | 0.72 (0.71 - 0.72) | -0.16 (-0.17 - -0.14) | 1.06 (1.04 - 1.08) |
| | Allotey 2020 for Any PE (b) [13] | 0.72 (0.72 - 0.72) | -0.15 (-0.17 - -0.14) | 0.88 (0.86 - 0.89) |
| | Tang 2022 for Any PE [14] | 0.69 (0.69 - 0.70) | 0.95 (0.94 - 0.97) | 0.61 (0.60 - 0.62) |
| | Tarca 2022 for Any PE [15] | 0.69 (0.69 - 0.70) | -0.62 (-0.64 - -0.61) | 1.37 (1.34 - 1.39) |
| | Plasencia 2007 for EOPE [6] | 0.57 (0.56 - 0.58) | -0.03 (-0.07 - 0.02) | 0.64 (0.59 - 0.69) |
| | Kuc 2013 for EOPE [16] | 0.66 (0.64 - 0.67) | -5.09 (-5.14 - -5.05) | 0.35 (0.32 - 0.38) |
| | Scazzocchio 2013 for EOPE [17] | 0.66 (0.65 - 0.68) | -1.09 (-1.14 - -1.04) | 0.64 (0.61 - 0.67) |
| | Baschat 2014 for EOPE [18] | 0.70 (0.69 - 0.71) | -1.08 (-1.13 - -1.04) | 2.88 (2.76 - 3.00) |
| | Teixeira 2014 for EOPE [19] | 0.70 (0.69 - 0.71) | -1.21 (-1.26 - -1.17) | 0.70 (0.66 - 0.73) |
| | Crovetto 2015 for EOPE [20] | 0.53 (0.51 - 0.54) | -1.63 (-1.68 - -1.58) | 0.39 (0.36 - 0.42) |
| | Allotey 2020 for EOPE (a) [13] | 0.68 (0.67 - 0.70) | -0.65 (-0.70 - -0.61) | 0.84 (0.80 - 0.87) |
| | Allotey 2020 for EOPE (b) [13] | 0.68 (0.67 - 0.70) | -0.76 (-0.81 - -0.71) | 0.88 (0.84 - 0.92) |
| | Plasencia 2007 for LOPE [6] | 0.68 (0.67 - 0.68) | 0.70 (0.69 - 0.72) | 0.57 (0.55 - 0.58) |
| | Poon 2010 for LOPE [21] | 0.67 (0.67 - 0.68) | 0.68 (0.67 - 0.70) | 0.59 (0.58 - 0.61) |
| | Kuc 2013 for LOPE [16] | 0.64 (0.64 - 0.65) | -2.09 (-2.11 - -2.08) | 0.60 (0.58 - 0.62) |
| | Scazzocchio 2013 for LOPE [17] | 0.68 (0.67 - 0.68) | -0.17 (-0.19 - -0.16) | 0.70 (0.68 - 0.71) |

| | | | | |
|---|---|---|---|---|
| | Crovetto 2015 for LOPE [20] | 0.55 (0.54 - 0.55) | -0.54 (-0.56 - -0.53) | 0.29 (0.28 - 0.31) |
| | Allotey 2020 for LOPE (a) [13] | 0.71 (0.70 - 0.71) | -0.06 (-0.08 - -0.05) | 0.98 (0.97 - 1.00) |
| | Allotey 2020 for LOPE (b) [13] | 0.71 (0.71 - 0.72) | 0.00 (-0.01 - 0.02) | 0.81 (0.79 - 0.83) |
| | Zhang 2025 for LOPE [22] | 0.68 (0.68 - 0.69) | -2.64 (-2.66 - -2.63) | 0.30 (0.30 - 0.31) |
| | Myers 2013 for Preterm PE [23] | 0.67 (0.66 - 0.67) | -6.01 (-6.04 - -5.98) | 0.63 (0.61 - 0.66) |
| | Rocha 2017 for Preterm PE (a) [12] | 0.65 (0.64 - 0.66) | -5.17 (-5.20 - -5.14) | 16.41 (15.87 - 16.95) |
| | Rocha 2017 for Preterm PE (b) [12] | 0.69 (0.68 - 0.69) | -5.20 (-5.23 - -5.17) | 17.58 (17.06 - 18.10) |
| | Sepúlveda-Martínez 2019 for Preterm PE [24] | 0.63 (0.62 - 0.64) | -0.61 (-0.64 - -0.58) | 0.82 (0.79 - 0.84) |
| | Rocha 2017 for Term PE [12] | 0.65 (0.65 - 0.66) | -4.04 (-4.06 - -4.02) | 9.75 (9.45 - 10.05) |
| | Lai 2013 for Intermediate PE [25] | 0.68 (0.67 - 0.68) | 0.09 (0.06 - 0.12) | 0.58 (0.56 - 0.60) |
| | Lai 2013 for Late PE [25] | 0.68 (0.65 - 0.70) | -0.07 (-0.09 - -0.05) | 0.71 (0.69 - 0.72) |
| Stillbirth | Yerlikaya 2016 for stillbirth ≥ 24 weeks [26] | 0.58 (0.57 - 0.58) | 0.47 (0.44 - 0.49) | 0.82 (0.77 - 0.87) |
| | Trudell 2017 for stillbirth ≥ 32 weeks [27] | 0.55 (0.54 - 0.55) | 0.86 (0.83 - 0.89) | 0.46 (0.39 - 0.53) |
| Small-for-gestational-age (SGA) | Macdonald-Wallis 2015 for SGA [11] | 0.66 (0.66 - 0.66) | 9.57 (9.56 - 9.58) | 0.71 (0.70 - 0.72) |
| | González-González 2017 for SGA [28] | 0.58 (0.58 - 0.58) | -0.46 (-0.47 - -0.45) | 0.65 (0.63 - 0.68) |
| | Adjahou 2025 for SGA [29] | 0.67 (0.67 - 0.67) | -0.23 (-0.23 - -0.22) | 0.73 (0.72 - 0.74) |
| | Poon 2011 (log(birthweight)) [30] | Not applicable | 113.63 g (112.55 - 114.71) | 0.93 (0.93 - 0.94) |

**Note:**

**PE:** pre-eclampsia
**EOPE:** early-onset pre-eclampsia - delivery < 34 gestation weeks with PE
**LOPE**: late-onset pre-eclampsia - delivery ≥ 34 gestation weeks with PE
**Preterm PE:** delivery < 37 gestation weeks with PE
**Term PE:** delivery ≥ 37 gestation weeks with PE
**Intermediate PE:** delivery between 34 to 37 gestation weeks with PE
**Late PE:** delivery ≥ 38 gestation weeks with PE

Additional file 1: Table S10 External validation results of models without intercept

| Model | Outcome | Model type | Equation | Number of predictors, N | Population Source | Reported C-Statistics | Validated C-Statistics |
|---|---|---|---|---|---|---|---|
| Naylor 1997 [42] | GDM | Logistic regression | logit(p) = α + 0.4700*(Age ≥35) + 0.5878*(BMI 22.1-25.0) + 1.1632*(BMI ≥25.1) - 0.3567*(Black race) + 1.5686*(Asian race) + 0.4700*(Ethnicity: other) | 6 | Canada | 0.68 | 0.70 (0.70 - 0.70) |
| Shirazian 2009 [43] | GDM | Logistic regression | logit(p) = α + 0.5128*(Age 25-29) + 1.5151*(Age ≥30) + 0.5128*(BMI 25.0-29.9) + 0.8920*(BMI ≥30.0) + 0.8416*(Family history of DM2) | 5 | Iran | Not reported | 0.67 (0.67 - 0.67) |
| Nanda 2011 (a) [44] | GDM | Logistic regression | logit(p) = α + 0.0583*(Maternal age) + 0.1133*(BMI) + 1.0043*(South Asian ethnicity) + 0.8879*(East Asian ethnicity) + 3.7226*(Parous with previous GDM) + 0.6780*(Parous with previous LGA) | 6 | UK | 0.788 | 0.75 (0.75 - 0.76) |
| Nanda 2011 (b, No previous GDM) [44] | GDM | Logistic regression | logit(p) = α + 0.0583*(Maternal age) + 0.1133*(BMI) + 0.8920*(South Asian ethnicity) + 0.9670*(East Asian ethnicity) - 0.3285*(Parous with no previous GDM) + 0.9969*(Parous with previous LGA) | 6 | UK | 0.738 | 0.73 (0.72 - 0.73) |
| Shen 2022 (a) [45] | GDM | Logistic regression | logit(p) = α + 0.1160*(Maternal age) - 0.0212*(Height) + 0.1266*(BMI) + 0.3846*(IVF) + 1.1684*(Chronic hypertension) | 5 | China | 0.708 | 0.72 (0.72 - 0.72) |
| Shen 2022 (b, parous) [45] | GDM | Logistic regression | logit(p) = α + 0.1089*(Maternal age) - 0.0325*(Height) + 0.0980*(BMI) + 0.0305*(Inter-pregnancy interval) + 0.1997*(Birth weight Z-score of last pregnancy) + 1.8413*(Previous GDM) | 6 | China | 0.735 | 0.77 (0.77 - 0.77) |
| Tranidou 2023 (a) [46] | GDM | Logistic regression | logit(p) = α + 0.0770*(Maternal age) + 0.0953*(Pre-pregnancy BMI) + 0.2070*(Smoking during pregnancy) | 3 | Greece | 0.672 | 0.71 (0.71 - 0.71) |
| Tranidou 2023 (b) [46] | GDM | Logistic regression | logit(p) = α + 0.0677*(Maternal age) + 0.0953*(Pre-pregnancy BMI) + 0.3646*(Conception ART) + 0.2231*(Thyroid disease) + 0.4947*(Chronic hypertension) + 0.1823*(SLE/APS) | 6 | Greece | 0.672 | 0.72 (0.71 - 0.72) |
| Akolekar 2011 for EOPE [47] | EOPE | Logistic regression | logit(p) = α + 0.0208*(Weight (kg)) - 0.0523*(Height (cm)) + 1.2931*(African ethnicity) + 0.9458*(South Asian ethnicity) + 0.7998*(Assisted conception) + 0.6471*(Family history of PE) + 1.7267*(Chronic hypertension) + 0.8042*(Parous with previous PE) - 1.0996*(Parous without previous PE) (shrinkage factor 0.95 reported) | 9 | UK | Not reported | 0.73 (0.72 - 0.73) |

| | | | | | | | |
|---|---|---|---|---|---|---|---|
| Akolekar 2011 for Intermediate PE [47] | Intermediate PE | Logistic regression | logit(p) = α + 0.0218*(Weight (kg)) - 0.0440*(Height (cm)) + 0.9791*(African ethnicity) + 0.8800*(South Asian ethnicity) + 0.7566*(Assisted conception) + 1.4346*(Chronic hypertension) + 1.2167*(Pre-existing diabetes) + 0.8800*(Parous with previous PE) - 0.8393*(Parous without previous PE) | 9 | UK | Not reported | 0.71 (0.69 - 0.72) |
| Akolekar 2011 for Late PE [47] | Late PE | Logistic regression | logit(p) = α + 0.0276*(Weight (kg)) - 0.0367*(Height (cm)) + 0.7528*(African ethnicity) + 1.1049*(Chronic hypertension) + 0.5961*(Parous with previous PE) - 1.2413*(Parous without previous PE) | 6 | UK | Not reported | 0.69 (0.69 - 0.69) |
| Suksai 2022 for Any PE [48] | Any PE | Logistic regression | logit(p) = α + 0.6081*(Maternal age ≥30) + 0.4606*(BMI ≥25) + 1.7347*(Multifetal pregnancy) + 1.7071*(Previous pre-eclampsia) + 1.2881*(Previous adverse perinatal outcomes) + 1.0123*(Pregnancy interval >10 years) + 1.0842*(Nulliparous) + 2.0620*(Renal disease) + 1.1976*(Chronic hypertension) + 0.9877*(SLE or APS) + 0.3702*(Diabetes mellitus) + 0.9651*(MAP ≥95 mmHg) | 12 | Thailand | 0.771 | 0.69 (0.68 - 0.70) |

**Note:**

**GDM**: gestational diabetes
**PE:** pre-eclampsia
**EOPE:** early-onset pre-eclampsia - delivery < 34 gestation weeks with PE
**Intermediate PE:** delivery between 34 to 37 gestation weeks with PE
**Late PE:** delivery ≥ 38 gestation weeks with PE

References


1. van Leeuwen M, Opmeer BC, Zweers EJ, van Ballegooie E, ter Brugge HG, de Valk HW, Visser GH, Mol BW: **Estimating the risk of gestational diabetes mellitus: a clinical prediction model based on patient characteristics and medical history**. *Bjog* 2010, **117**(1):69-75.
2. Sweeting AN, Appelblom H, Ross GP, Wong J, Kouru H, Williams PF, Sairanen M, Hyett JA: **First trimester prediction of gestational diabetes mellitus: A clinical model based on maternal demographic parameters**. *Diabetes Res Clin Pract* 2017, **127**:44-50.
3. Benhalima K, Van Crombrugge P, Moyson C, Verhaeghe J, Vandeginste S, Verlaenen H, Vercammen C, Maes T, Dufraimont E, De Block C *et al*: **Estimating the risk of gestational diabetes mellitus based on the 2013 WHO criteria: a prediction model based on clinical and biochemical variables in early pregnancy**. *Acta Diabetol* 2020, **57**(6):661-671.
4. Basil B, Mba IN, Myke-Mbata BK, Adebisi SA, Oghagbon EK: **A first trimester prediction model and nomogram for gestational diabetes mellitus based on maternal clinical risk factors in a resource-poor setting**. *BMC Pregnancy Childbirth* 2024, **24**(1):346.
5. Syngelaki A, Wright A, Gomez Fernandez C, Mitsigiorgi R, Nicolaides KH: **First-Trimester Prediction of Gestational Diabetes Mellitus Based on Maternal Risk Factors**. *Bjog* 2025, **132**(7):972-982.
6. Plasencia W, Maiz N, Bonino S, Kaihura C, Nicolaides KH: **Uterine artery Doppler at 11 + 0 to 13 + 6 weeks in the prediction of pre-eclampsia**. *Ultrasound in Obstetrics & Gynecology* 2007, **30**(5):742-749.
7. Emonts P, Seaksan S, Seidel L, Thoumsin H, Gaspard U, Albert A, Foidart JM: **Prediction of maternal predisposition to preeclampsia**. *Hypertens Pregnancy* 2008, **27**(3):237-245.
8. Poon LC, Kametas N, Bonino S, Vercellotti E, Nicolaides KH: **Urine albumin concentration and albumin-to-creatinine ratio at 11(+0) to 13(+6) weeks in the prediction of pre-eclampsia**. *Bjog* 2008, **115**(7):866-873.
9. Poon LC, Kametas NA, Pandeva I, Valencia C, Nicolaides KH: **Mean arterial pressure at 11(+0) to 13(+6) weeks in the prediction of preeclampsia**. *Hypertension* 2008, **51**(4):1027-1033.
10. Direkvand-Moghadam A, Khosravi A, Sayehmiri K: **Predictive factors for preeclampsia in pregnant women: a Receiver Operation Character approach**. *Arch Med Sci* 2013, **9**(4):684-689.
11. Macdonald-Wallis C, Silverwood RJ, de Stavola BL, Inskip H, Cooper C, Godfrey KM, Crozier S, Fraser A, Nelson SM, Lawlor DA, Tilling K: **Antenatal blood pressure for prediction of pre-eclampsia, preterm birth, and small for gestational age babies: development and validation in two general population cohorts**. *Bmj* 2015, **351**:h5948.
12. Rocha RS, Alves JAG, Maia EHMSB, Araujo Júnior E, Peixoto AB, Santana EFM, Martins WP, Vasconcelos CTM, Da Silva Costa F, Oriá MOB: **Simple approach based on maternal characteristics and mean arterial pressure for the prediction of preeclampsia in the first trimester of pregnancy**. *J Perinat Med* 2017, **45**(7):843-849.
13. Allotey J, Snell KI, Smuk M, Hooper R, Chan CL, Ahmed A, Chappell LC, von Dadelszen P, Dodds J, Green M *et al*: **Validation and development of models using clinical, biochemical and ultrasound markers for predicting pre-eclampsia: an individual participant data meta-analysis**. *Health Technol Assess* 2020, **24**(72):1-252.
14. Tang Z, Ji Y, Zhou S, Su T, Yuan Z, Han N, Jia J, Wang H: **Development and Validation of Multi-Stage Prediction Models for Pre-eclampsia: A Retrospective Cohort Study on Chinese Women**. *Front Public Health* 2022, **10**:911975.
15. Tarca AL, Taran A, Romero R, Jung E, Paredes C, Bhatti G, Ghita C, Chaiworapongsa T, Than NG, Hsu CD: **Prediction of preeclampsia throughout gestation with maternal characteristics and biophysical and biochemical markers: a longitudinal study**. *Am J Obstet Gynecol* 2022, **226**(1):126.e121-126.e122.
16. Kuc S, Koster MP, Franx A, Schielen PC, Visser GH: **Maternal characteristics, mean arterial pressure and serum markers in early prediction of preeclampsia**. *PLoS One* 2013, **8**(5):e63546.
17. Scazzocchio E, Figueras F, Crispi F, Meler E, Masoller N, Mula R, Gratacos E: **Performance of a first-trimester screening of preeclampsia in a routine care low-risk setting**. *Am J Obstet Gynecol* 2013, **208**(3):203.e201-203.e210.

18. Baschat AA, Magder LS, Doyle LE, Atlas RO, Jenkins CB, Blitzer MG: **Prediction of preeclampsia utilizing the first trimester screening examination**. *Am J Obstet Gynecol* 2014, **211**(5):514.e511-517.
19. Teixeira C, Tejera E, Martins H, Pereira AT, Costa-Pereira A, Rebelo I: **First trimester aneuploidy screening program for preeclampsia prediction in a portuguese obstetric population**. *Obstet Gynecol Int* 2014, **2014**:435037.
20. Crovetto F, Figueras F, Triunfo S, Crispi F, Rodriguez-Sureda V, Dominguez C, Llurba E, Gratacós E: **First trimester screening for early and late preeclampsia based on maternal characteristics, biophysical parameters, and angiogenic factors**. *Prenat Diagn* 2015, **35**(2):183-191.
21. Poon LC, Kametas NA, Chelemen T, Leal A, Nicolaides KH: **Maternal risk factors for hypertensive disorders in pregnancy: a multivariate approach**. *J Hum Hypertens* 2010, **24**(2):104-110.
22. Zhang Y, Gu X, Yang N, Xue Y, Ma L, Wang Y, Zhang H, Jia K: **Prediction Models for Late-Onset Preeclampsia: A Study Based on Logistic Regression, Support Vector Machine, and Extreme Gradient Boosting Models**. *Biomedicines* 2025, **13**(2).
23. Myers JE, Kenny LC, McCowan LM, Chan EH, Dekker GA, Poston L, Simpson NA, North RA: **Angiogenic factors combined with clinical risk factors to predict preterm pre-eclampsia in nulliparous women: a predictive test accuracy study**. *Bjog* 2013, **120**(10):1215-1223.
24. Sepúlveda-Martínez A, Rencoret G, Silva MC, Ahumada P, Pedraza D, Muñoz H, Valdés E, Parra-Cordero M: **First trimester screening for preterm and term pre-eclampsia by maternal characteristics and biophysical markers in a low-risk population**. *J Obstet Gynaecol Res* 2019, **45**(1):104-112.
25. Lai J, Poon LC, Pinas A, Bakalis S, Nicolaides KH: **Uterine artery Doppler at 30-33 weeks' gestation in the prediction of preeclampsia**. *Fetal Diagn Ther* 2013, **33**(3):156-163.
26. Yerlikaya G, Akolekar R, McPherson K, Syngelaki A, Nicolaides KH: **Prediction of stillbirth from maternal demographic and pregnancy characteristics**. *Ultrasound Obstet Gynecol* 2016, **48**(5):607-612.
27. Trudell AS, Tuuli MG, Colditz GA, Macones GA, Odibo AO: **A stillbirth calculator: Development and internal validation of a clinical prediction model to quantify stillbirth risk**. *PLoS One* 2017, **12**(3):e0173461.
28. González-González NL, González-Dávila E, González Marrero L, Padrón E, Conde JR, Plasencia W: **Value of placental volume and vascular flow indices as predictors of intrauterine growth retardation**. *Eur J Obstet Gynecol Reprod Biol* 2017, **212**:13-19.
29. Adjahou S, Syngelaki A, Nanda M, Papavasileiou D, Akolekar R, Nicolaides KH: **Routine 36-week scan: prediction of small-for-gestational-age neonate**. *Ultrasound Obstet Gynecol* 2025, **65**(1):20-29.
30. Poon LC, Karagiannis G, Staboulidou I, Shafiei A, Nicolaides KH: **Reference range of birth weight with gestation and first-trimester prediction of small-for-gestation neonates**. *Prenat Diagn* 2011, **31**(1):58-65.
31. Kotzaeridi G, Blätter J, Eppel D, Rosicky I, Mittlböck M, Yerlikaya-Schatten G, Schatten C, Husslein P, Eppel W, Huhn EA *et al*: **Performance of early risk assessment tools to predict the later development of gestational diabetes**. *Eur J Clin Invest* 2021, **51**(12):e13630.
32. Meertens LJE, Scheepers HCJ, van Kuijk SMJ, Roeleveld N, Aardenburg R, van Dooren IMA, Langenveld J, Zwaan IM, Spaanderman MEA, van Gelder M, Smits LJM: **External validation and clinical utility of prognostic prediction models for gestational diabetes mellitus: A prospective cohort study**. *Acta Obstet Gynecol Scand* 2020, **99**(7):891-900.
33. Thériault S, Forest JC, Massé J, Giguère Y: **Validation of early risk-prediction models for gestational diabetes based on clinical characteristics**. *Diabetes Res Clin Pract* 2014, **103**(3):419-425.
34. van Hoorn F, Koster M, Naaktgeboren CA, Groenendaal F, Kwee A, Lamain-de Ruiter M, Franx A, Bekker MN: **Prognostic models versus single risk factor approach in first-trimester selective screening for gestational diabetes mellitus: a prospective population-based multicentre cohort study**. *Bjog* 2021, **128**(4):645-654.
35. Allotey J, Whittle R, Snell KIE, Smuk M, Townsend R, von Dadelszen P, Heazell AEP, Magee L, Smith GCS, Sandall J *et al*: **External validation of prognostic models to predict stillbirth using International Prediction of Pregnancy Complications (IPPIC) Network database: individual participant data meta-analysis**. *Ultrasound Obstet Gynecol* 2022, **59**(2):209-219.

36. Lamain-de Ruiter M, Kwee A, Naaktgeboren CA, Louhanepessy RD, De Groot I, Evers IM, Groenendaal F, Hering YR, Huisjes AJM, Kirpestein C *et al*: **External validation of prognostic models for preeclampsia in a Dutch multicenter prospective cohort**. *Hypertens Pregnancy* 2019, **38**(2):78-88.
37. Meertens LJE, Scheepers HCJ, van Kuijk SMJ, Aardenburg R, van Dooren IMA, Langenveld J, van Wijck AM, Zwaan I, Spaanderman MEA, Smits LJM: **External Validation and Clinical Usefulness of First Trimester Prediction Models for the Risk of Preeclampsia: A Prospective Cohort Study**. *Fetal Diagn Ther* 2019, **45**(6):381-393.
38. Snell KIE, Allotey J, Smuk M, Hooper R, Chan C, Ahmed A, Chappell LC, Von Dadelszen P, Green M, Kenny L *et al*: **External validation of prognostic models predicting pre-eclampsia: individual participant data meta-analysis**. *BMC Med* 2020, **18**(1):302.
39. Allen RE, Zamora J, Arroyo-Manzano D, Velauthar L, Allotey J, Thangaratinam S, Aquilina J: **External validation of preexisting first trimester preeclampsia prediction models**. *Eur J Obstet Gynecol Reprod Biol* 2017, **217**:119-125.
40. Meertens L, Smits L, van Kuijk S, Aardenburg R, van Dooren I, Langenveld J, Zwaan IM, Spaanderman M, Scheepers H: **External validation and clinical usefulness of first-trimester prediction models for small- and large-for-gestational-age infants: a prospective cohort study**. *BJOG* 2019, **126**(4):472-484.
41. Allotey J, Archer L, Coomar D, Snell KI, Smuk M, Oakey L, Haqnawaz S, Betrán AP, Chappell LC, Ganzevoort W *et al*: **Development and validation of prediction models for fetal growth restriction and birthweight: an individual participant data meta-analysis**. *Health Technol Assess* 2024, **28**(47):1-119.
42. Naylor CD, Sermer M, Chen E, Farine D: **Selective screening for gestational diabetes mellitus. Toronto Trihospital Gestational Diabetes Project Investigators**. *N Engl J Med* 1997, **337**(22):1591-1596.
43. Shirazian N, Emdadi R, Mahboubi M, Motevallian A, Fazel-Sarjuei Z, Sedighpour N, Fadaki SF, Shahmoradi N: **Screening for gestational diabetes: usefulness of clinical risk factors**. *Arch Gynecol Obstet* 2009, **280**(6):933-937.
44. Nanda S, Savvidou M, Syngelaki A, Akolekar R, Nicolaides KH: **Prediction of gestational diabetes mellitus by maternal factors and biomarkers at 11 to 13 weeks**. *Prenat Diagn* 2011, **31**(2):135-141.
45. Shen L, Sahota DS, Chaemsaithong P, Tse WT, Chung MY, Ip JKH, Leung TY, Poon LCY: **First Trimester Screening for Gestational Diabetes Mellitus with Maternal Factors and Biomarkers**. *Fetal Diagn Ther* 2022, **49**(5-6):256-264.
46. Tranidou A, Tsakiridis I, Apostolopoulou A, Xenidis T, Pazaras N, Mamopoulos A, Athanasiadis A, Chourdakis M, Dagklis T: **Prediction of Gestational Diabetes Mellitus in the First Trimester of Pregnancy Based on Maternal Variables and Pregnancy Biomarkers**. *Nutrients* 2023, **16**(1).
47. Akolekar R, Syngelaki A, Sarquis R, Zvanca M, Nicolaides KH: **Prediction of early, intermediate and late pre-eclampsia from maternal factors, biophysical and biochemical markers at 11-13 weeks**. *Prenat Diagn* 2011, **31**(1):66-74.
48. Suksai M, Geater A, Phumsiripaiboon P, Suntharasaj T: **A new risk score model to predict preeclampsia using maternal factors and mean arterial pressure in early pregnancy**. *J Obstet Gynaecol* 2022, **42**(3):437-442.
