## Additional file 2 for "Systematic Review and External Validation of Clinical Prediction Models for Adverse Pregnancy Outcomes Using Routinely Collected Pre-Conception and Early Pregnancy Data"

**Note: References in this document correspond to the reference list at the end of this file.**

Additional file 2: Figure S1 Flow chart of building baseline cohort for primary analysis

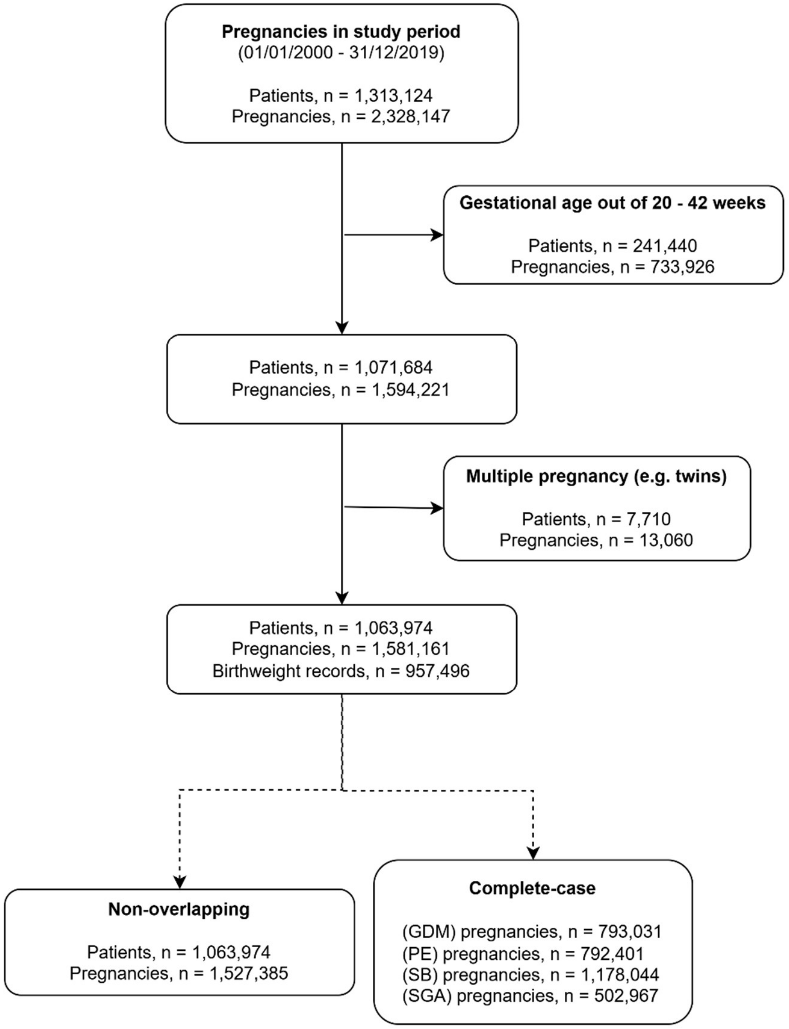

Figure S1. Flow chart of cohort selection for the primary and sensitivity analyses.

Sequential exclusions for gestational length outside 20–42 weeks and multiple pregnancy derived the primary cohort (1,581,161 pregnancies); dashed arrows indicate the non-overlapping and complete-case sensitivity cohorts. Abbreviations: GDM, gestational diabetes mellitus; PE, pre-eclampsia; SB, stillbirth; SGA, small-for-gestational-age.

Additional file 2: Figure S2 Calibration plots of models in primary analysis

### Gestational diabetes

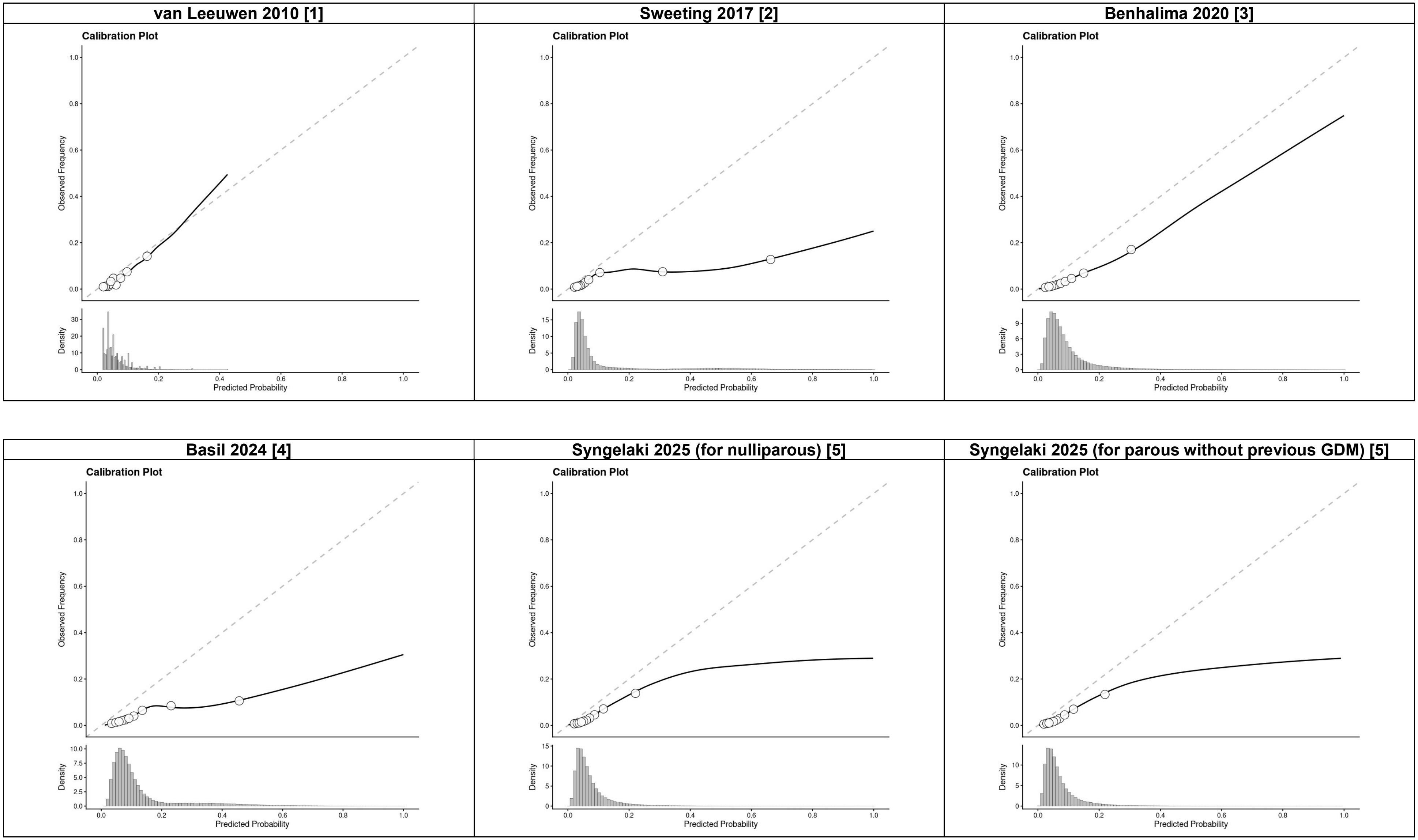

| Syngelaki 2025 (for parous with previous GDM) [5] |
|---|
| 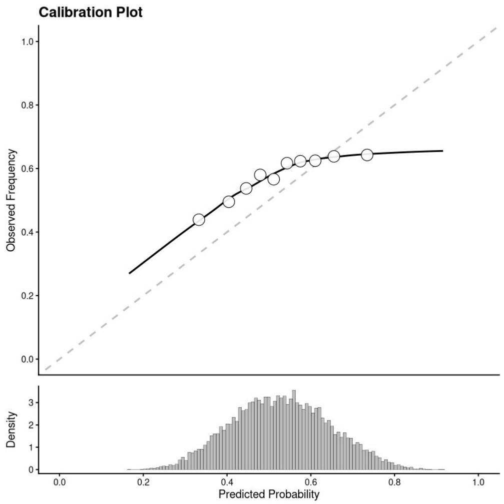 |

### Pre-eclampsia

#### Plasencia 2007 for Any PE [6]

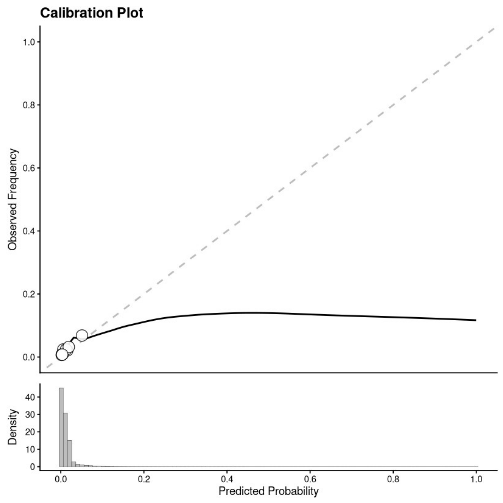

#### Emonts 2008 for Any PE [7]

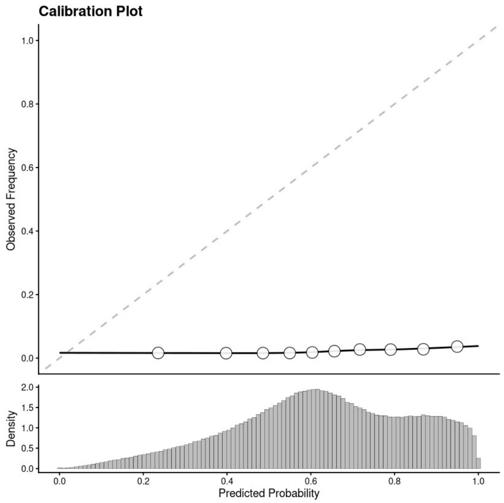

#### Poon 2008 for Any PE (1) [8]

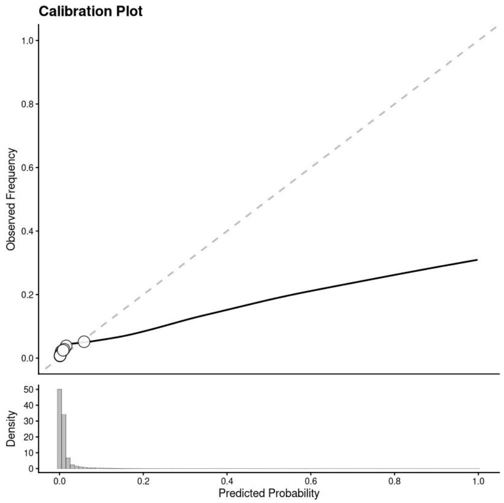

#### Poon 2008 for Any PE (2) [9]

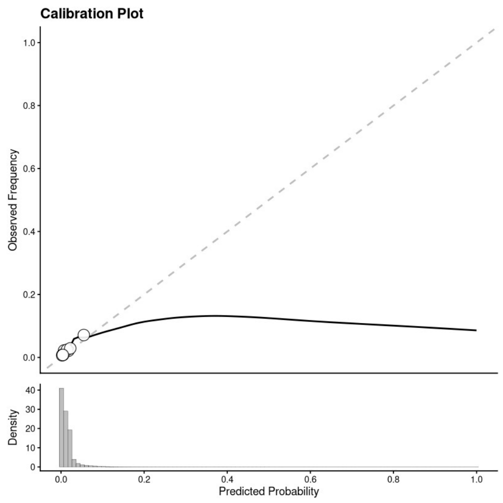

#### Macdonald-Wallis 2015 for Any PE [10]

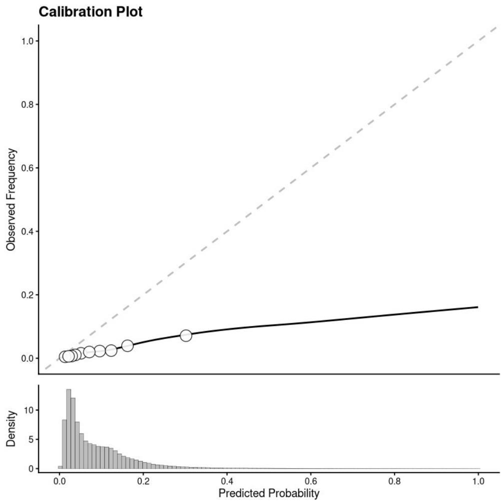

#### Rocha 2017 for Any PE (a) [11]

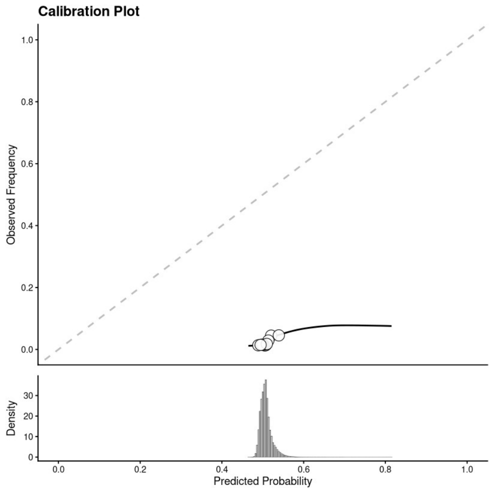

**Rocha 2017 for Any PE (b) [11]**

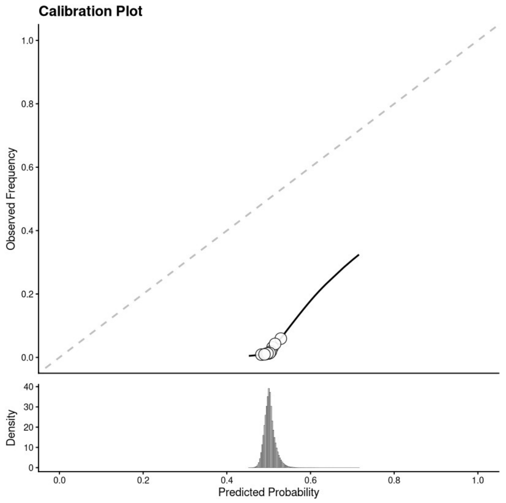

**Allotey 2020 for Any PE (a) [12]**

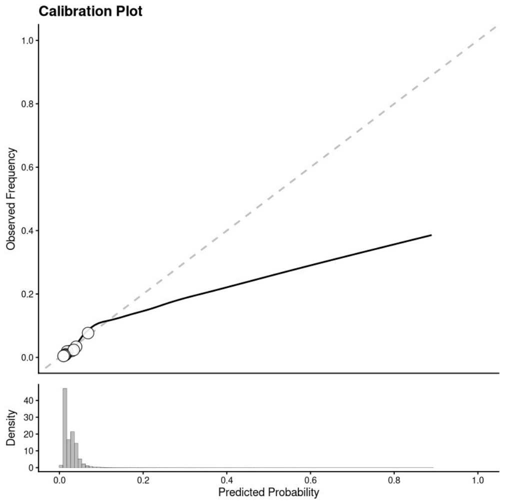

**Allotey 2020 for Any PE (b) [12]**

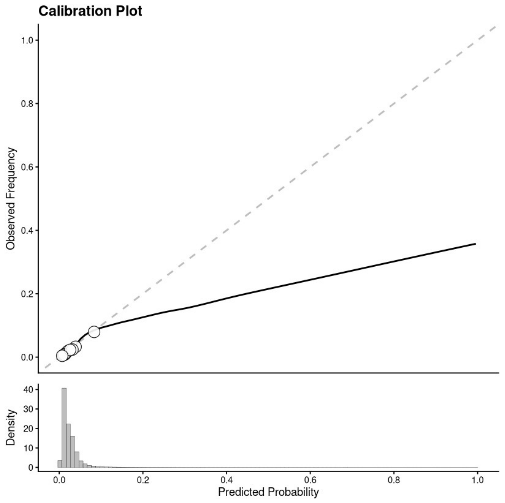

**Tang 2022 for Any PE [13]**

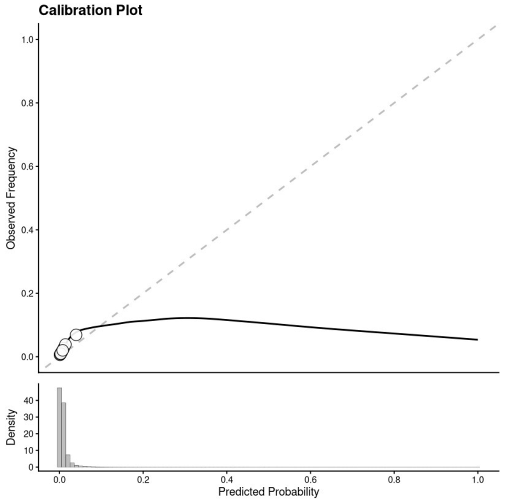

**Tarca 2022 for Any PE [14]**

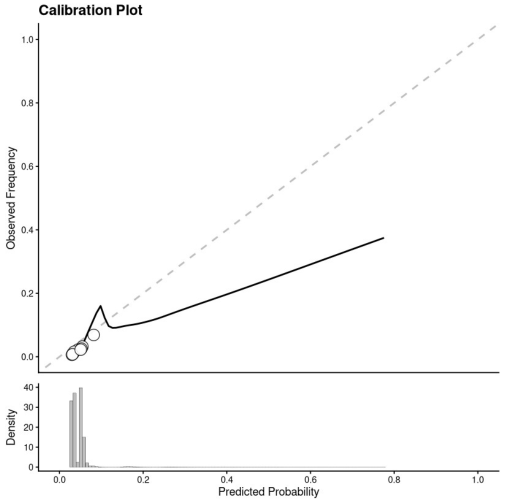

**Note:**
Calibration plots of Direkvand-Moghadam 2013 for Any PE [15] cannot be meaningfully generated due to insufficient spread of predicted probabilities.

Pre-eclampsia (early-onset)

**Kuc 2013 for EOPE [16]**

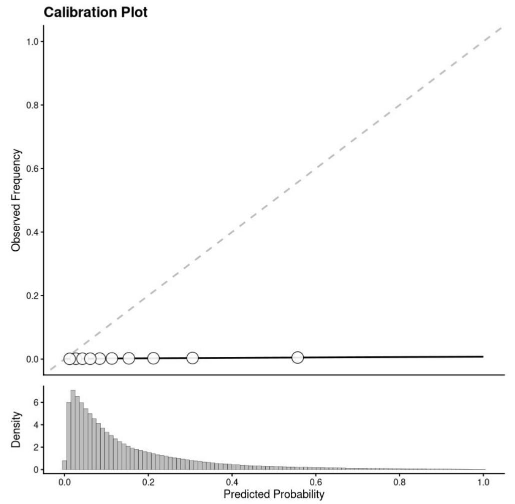

**Scazzocchio 2013 for EOPE [17]**

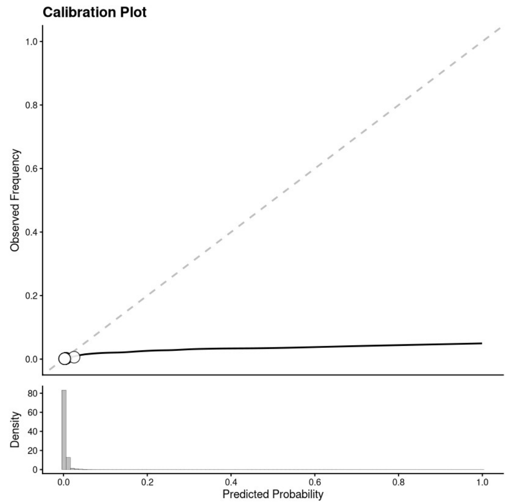

**Baschat 2014 for EOPE [18]**

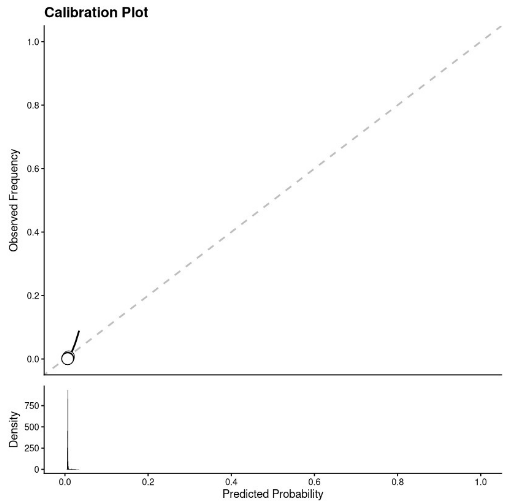

**Crovetto 2015 for EOPE [19]**

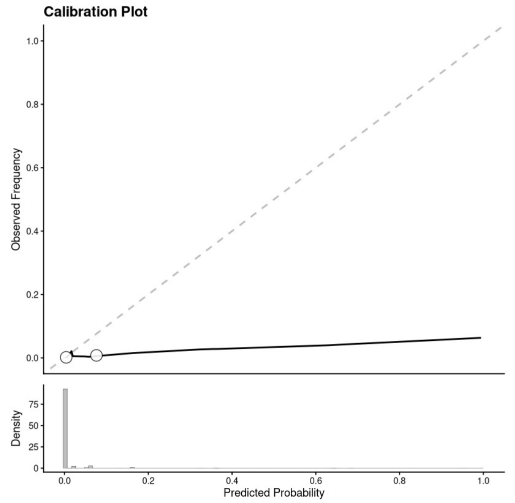

**Teixeira 2014 for EOPE [20]**

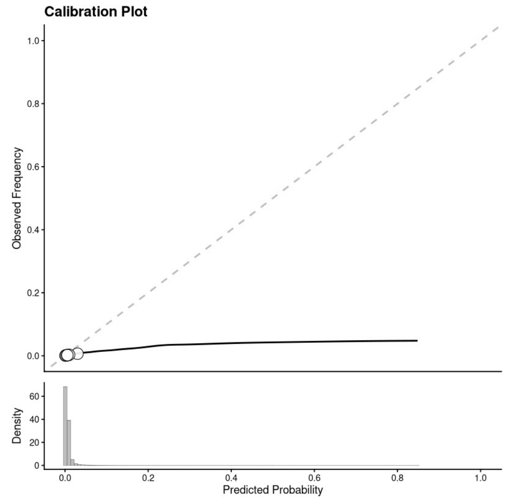

**Allotey 2020 for EOPE (a) [12]**

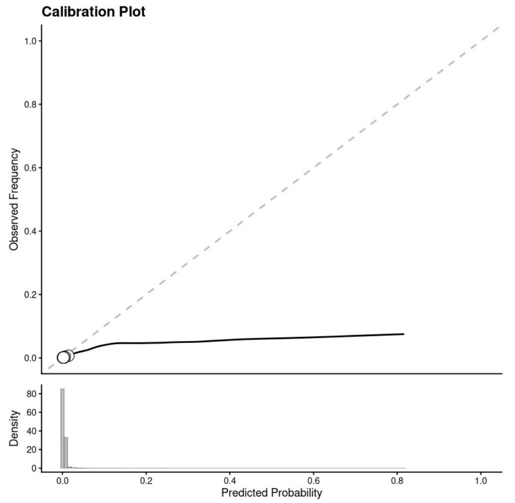

| Allotey 2020 for EOPE (b) [12] | **Note:** |
|---|---|
| 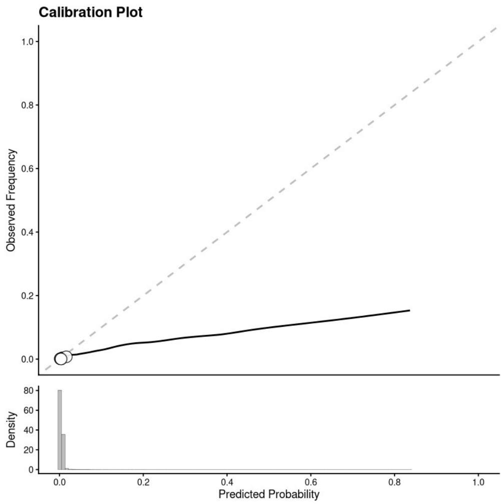 | Calibration plots of Plasencia 2007 for EOPE **[6]** cannot be meaningfully generated due to insufficient spread of predicted probabilities. |

### Pre-eclampsia (late-onset)

#### Plasencia 2007 for LOPE [6]

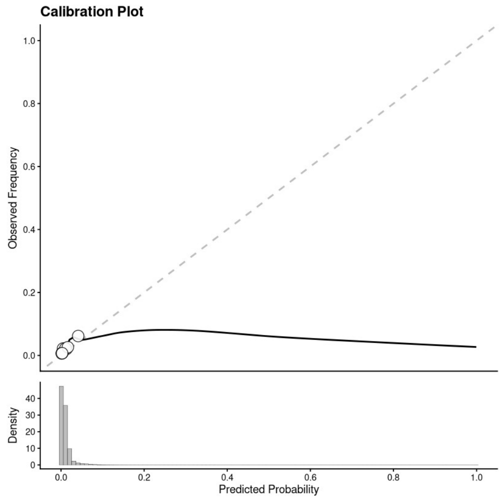

#### Poon 2010 for LOPE [21]

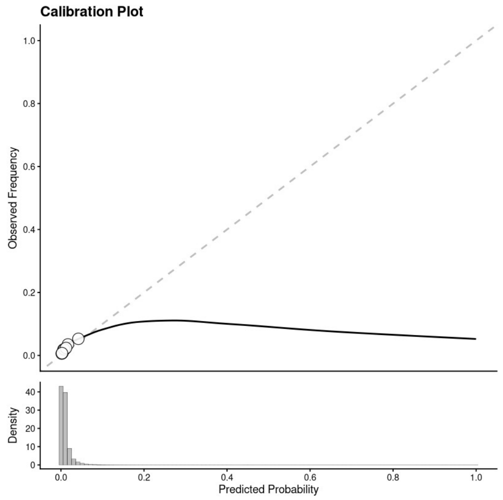

#### Kuc 2013 for LOPE [16]

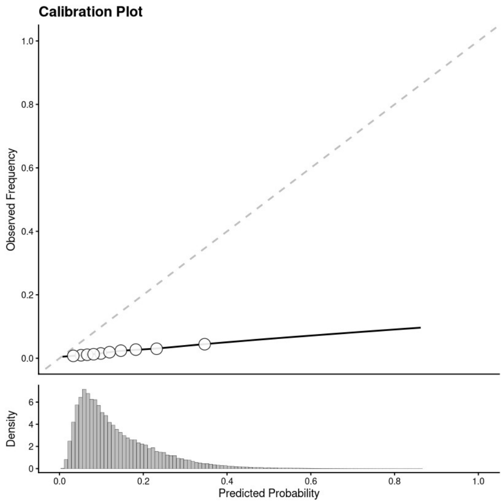

#### Scazzocchio 2013 for LOPE [17]

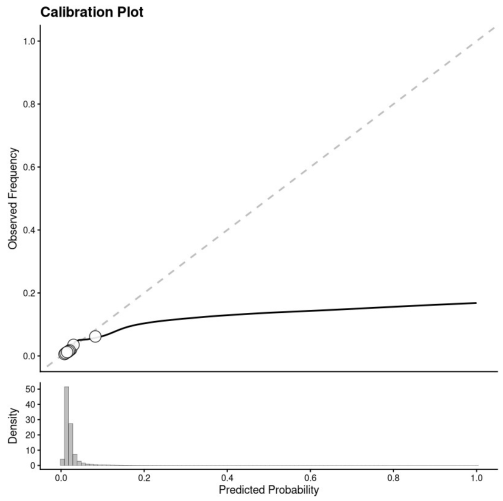

#### Crovetto 2015 for LOPE [19]

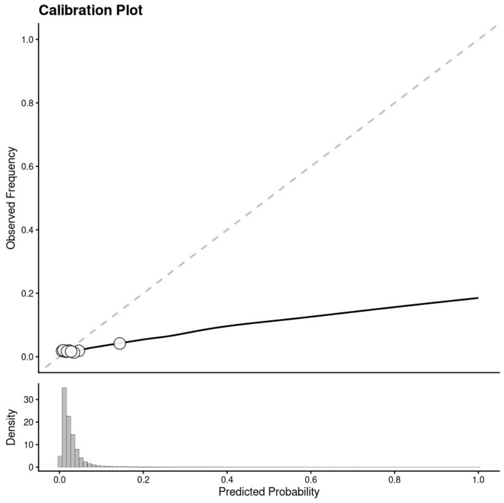

#### Allotey 2020 for LOPE (a) [12]

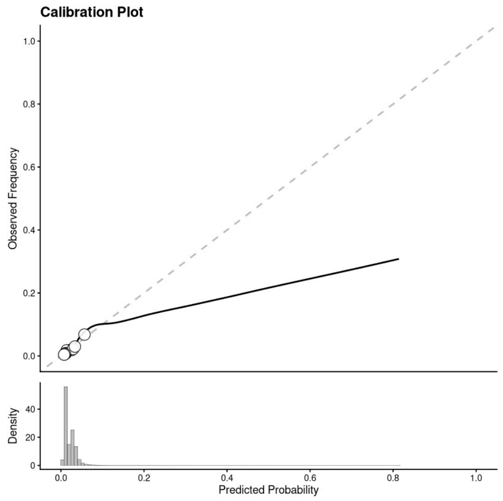

### Allotey 2020 for LOPE (b) [12]

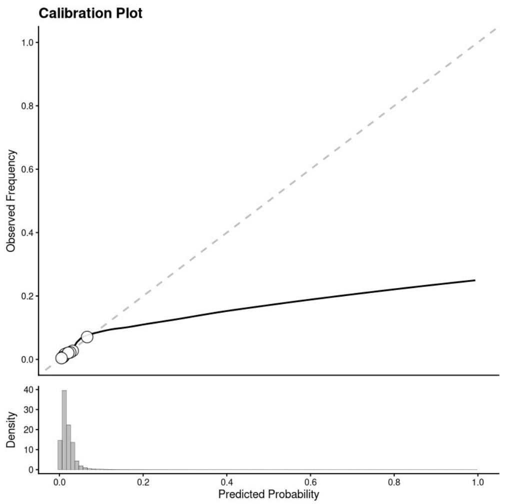

### Zhang 2025 for LOPE [22]

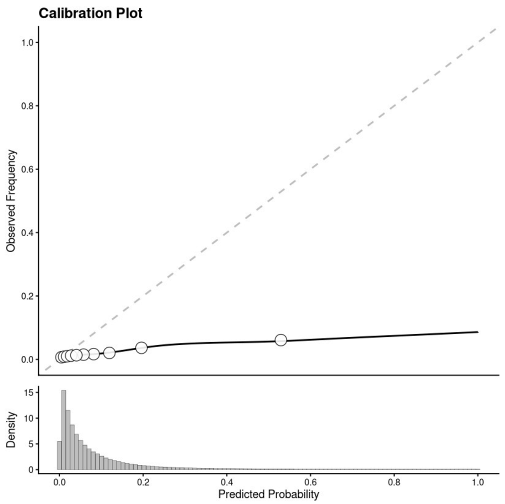

### Pre-eclampsia (other)

#### Rocha 2017 for Preterm PE (a) [11]

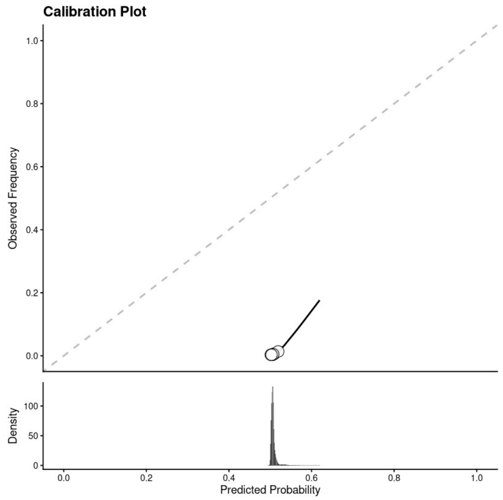

#### Rocha 2017 for Preterm PE (b) [11]

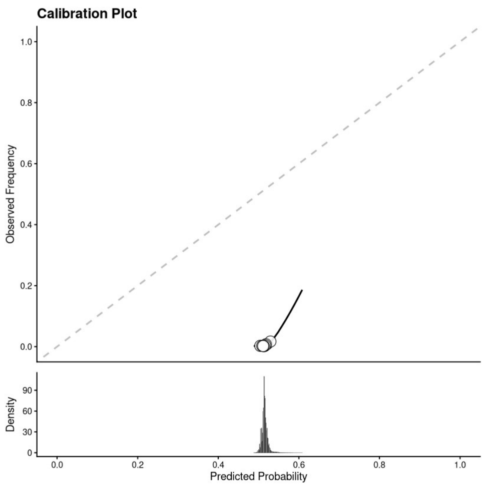

#### Sepúlveda-Martínez 2019 for Preterm PE [23]

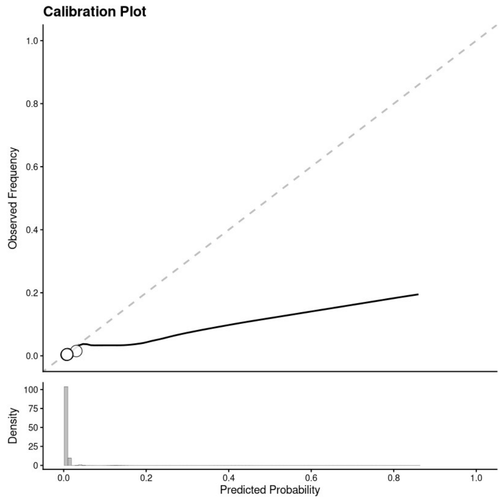

#### Myers 2013 for Preterm PE [24]

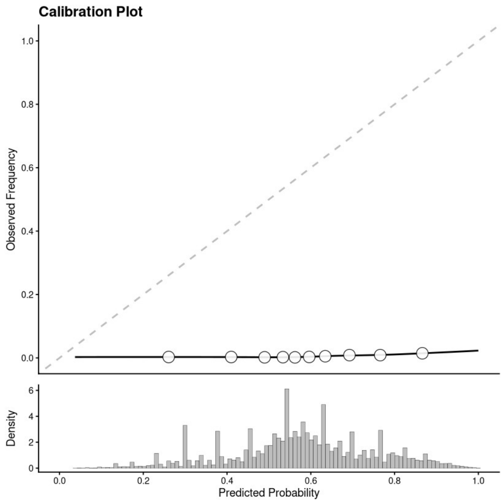

#### Rocha 2017 for Term PE [11]

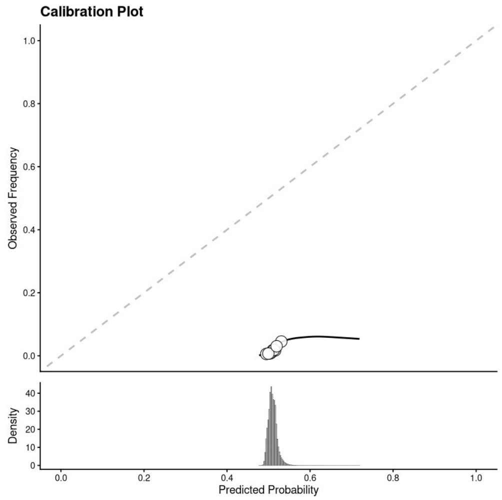

#### Lai 2013 for Intermediate PE [25]

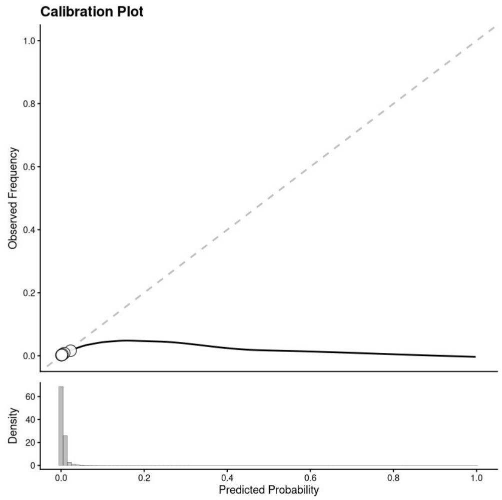

| Lai 2013 for Late PE [25] |
|---|
| 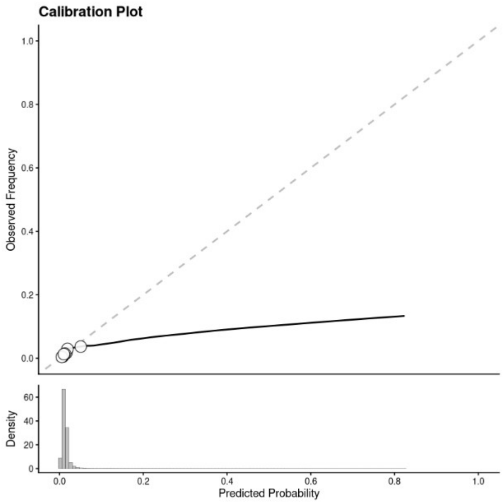 |

### Stillbirth

| Yerlikaya 2016 for stillbirth ≥ 24 weeks [26] | Trudell 2017 for stillbirth ≥ 32 weeks [27] |
| --- | --- |
| 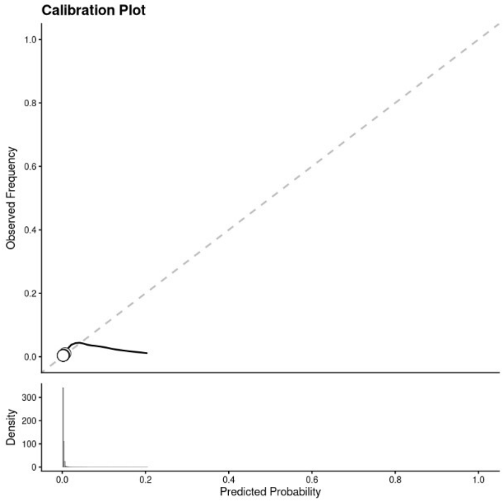  | 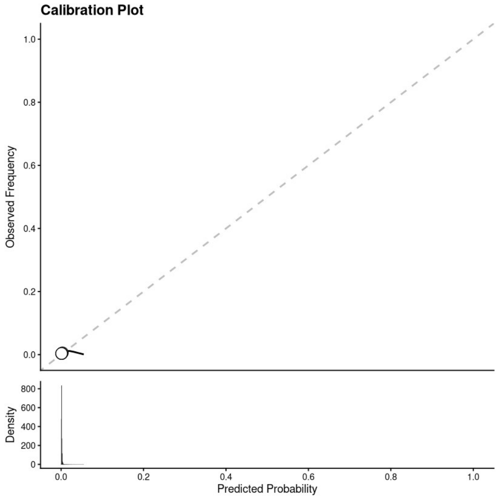  |

Small-for-gestational-age

**Macdonald-Wallis 2015 for SGA [10]**

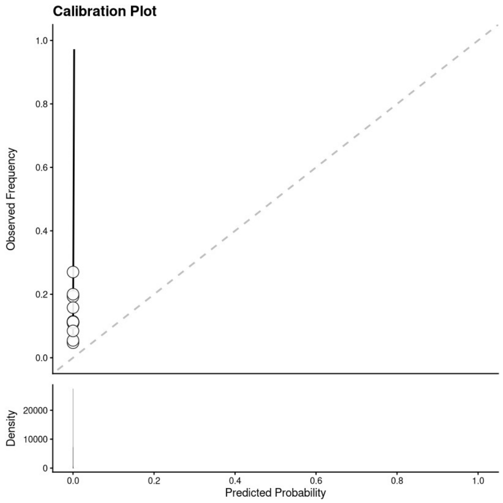

**Adjahou 2025 for SGA [28]**

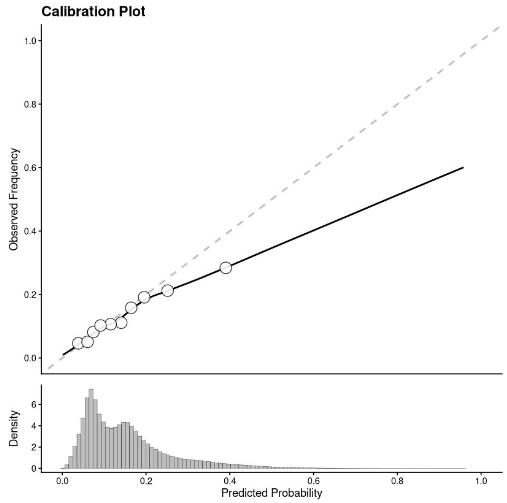

**Poon 2011 (log(birthweight)) [29]**

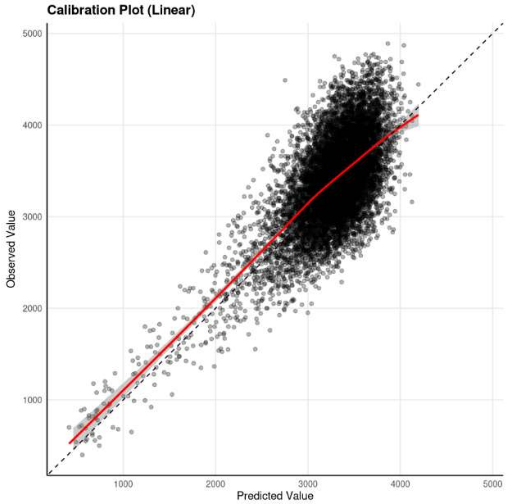

**Note:**
Calibration plots of González-González 2017 for SGA [30] cannot be meaningfully generated due to insufficient spread of predicted probabilities.

Figure S2. Calibration plots of models in the primary analysis.
Pooled calibration plots are displayed for 45 of 48 models: 44 binary models (smoothed curve, quantile circles, dashed ideal line, and density histogram) and 1 continuous model (Poon 2011: observed vs predicted birthweight in grams, with 95% confidence band). Three binary models were excluded for insufficient predicted probability spread.

Additional file 2: Figure S3 Validation metrics distribution of sensitivity analysis

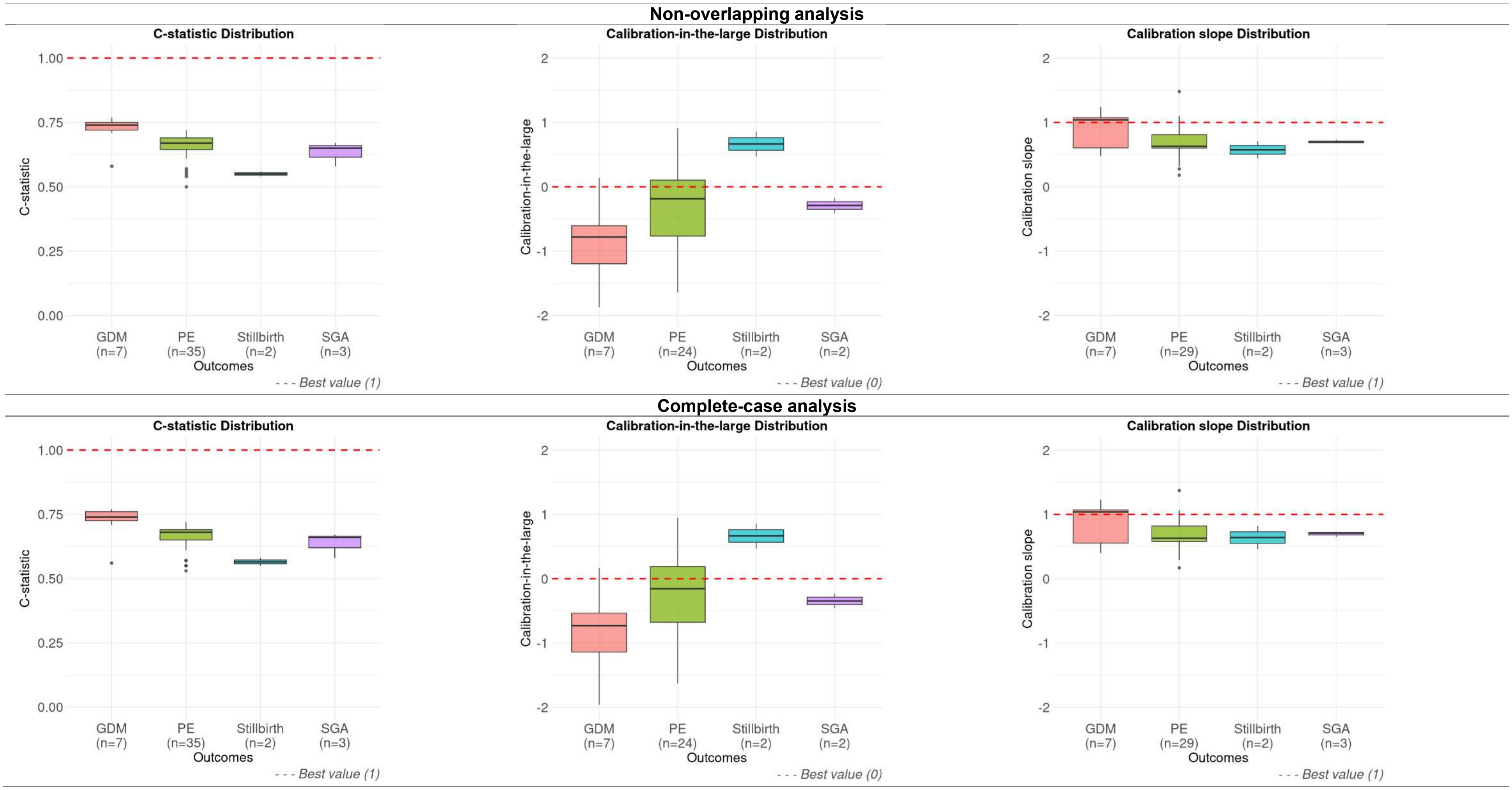

Figure S3. Validation performance by outcome in the sensitivity analyses.

Boxplots show the C-statistic, calibration-in-the-large, and calibration slope for the non-overlapping (top) and complete-case (bottom) analyses, with red dashed lines indicating ideal values (1.00 for C-statistic and slope; 0 for calibration-in-the-large). The continuous model (all panels) and values outside -2 to 2 (respective calibration panels) were excluded. Abbreviations: GDM, gestational diabetes mellitus; PE, pre-eclampsia; SGA, small-for-gestational-age.
