## Additional file 3 for "Systematic Review and External Validation of Clinical Prediction Models for Adverse Pregnancy Outcomes Using Routinely Collected Pre-Conception and Early Pregnancy Data"

Additional file 3: Table S11: Excluded models and reasons (a) - gestational diabetes

| First Author | Publish Year | Reason for exclusion | Development paper link / PMID |
|---|---|---|---|
| Naylor | 1997 | No intercept reported. | 9371855 |
| Shirazian | 2009 | No intercept reported. | 19301026 |
| Nanda (2 models) | 2011 | No intercept reported. | 21268030 |
| Shen L (a, 2 models) | 2022 | No intercept reported. | 35671743 |
| Shen L (b, 2 models) | 2022 | Biomarker or genetic predictors. | 35671743 |
| Tranidou (a, 2 models) | 2023 | No intercept reported. | 38201950 |
| Tranidou (b, 3 models) | 2023 | Biomarker or genetic predictors. | 38201950 |
| Tran | 2013 | Less than 3 predictors.<br>No intercept reported. | 23160727 |
| Donovan | 2019 | Missing coefficients of some predictors. | 30978258 |
| Gabbay-Benziv | 2014 | Some predictors not available. | 25153547 |
| Phaloprakarn | 2009 | Some predictors not available. | 19446948 |
| Teede | 2011 | Some predictors not available. | 21951203 |
| Syngelaki | 2011 | Some predictors not available. | 22067258 |
| Pintaudi | 2014 | Some predictors not available. | 24114434 |
| Syngelaki | 2015 | Some predictors not available. | 25531073 |
| Snyder (characteristic only) | 2020 | Some predictors not available. | 32272192 |
| Wu YQ (2 models) | 2024 | Some predictors not available. | 38622619 |
| Cooray (4 models) | 2023 | Some predictors not available. | 37659584 |
| Caliskan | 2004 | Some predictors not available. | 15144332 |
| Schaefer | 2018 | Biomarker or genetic predictors. | 30030928 |
| Savvidou | 2010 | Biomarker or genetic predictors. | 20876721 |
| Göbl | 2012 | Biomarker or genetic predictors. | 23001377 |
| Savona-Ventura | 2013 | Biomarker or genetic predictors. | 23279935 |
| Lovati | 2013 | Biomarker or genetic predictors. | 23642968 |
| Eleftheriades (characteristic only) | 2014 | Biomarker or genetic predictors. | 25173717 |
| Thériault | 2015 | Biomarker or genetic predictors. | 26351946 |
| Capula | 2016 | Biomarker or genetic predictors. | 27268754 |
| Zhao M | 2016 | Biomarker or genetic predictors. | 27916067 |
| Nombo | 2018 | Biomarker or genetic predictors. | 29852237 |
| Sweeting | 2018 | Biomarker or genetic predictors. | 29898442 |
| Zheng T | 2019 | Biomarker or genetic predictors. | 31324151 |
| Benhalima (b) | 2020 | Biomarker or genetic predictors. | 31915927 |
| Gao S | 2020 | Biomarker or genetic predictors. | 32327440 |
| Liu H | 2020 | Biomarker or genetic predictors. | 32845061 |
| Wu YT | 2020 | Biomarker or genetic predictors. | 33351102 |
| Xiong Y | 2020 | Biomarker or genetic predictors. | 32762275 |
| Zhang Y | 2020 | Biomarker or genetic predictors. | 32536997 |
| Guo F | 2020 | Biomarker or genetic predictors. | 31959134 |
| Garmendia | 2020 | Biomarker or genetic predictors. | 31671205 |
| Cremona | 2021 | Biomarker or genetic predictors. | 34620334 |
| Kang M | 2021 | Biomarker or genetic predictors. | 34956091 |
| Liu C | 2021 | Biomarker or genetic predictors. | 34970221 |
| Wang X | 2021 | Biomarker or genetic predictors. | 34803906 |
| Wang YM | 2021 | Biomarker or genetic predictors. | 33993435 |
| Zhang XM | 2021 | Biomarker or genetic predictors. | 33277541 |
| Zhao M | 2021 | Biomarker or genetic predictors. | 33795771 |
| Wang J (2 models) | 2021 | Biomarker or genetic predictors. | 34879850 |

| | | | |
|---|---|---|---|
| Zheng Y | 2022 | Biomarker or genetic predictors. | 35387184 |
| Du | 2022 | Biomarker or genetic predictors. | 38711005 |
| Kumar (3 models) | 2022 | Biomarker or genetic predictors. | 35124096 |
| Belsti (4 models) | 2023 | Biomarker or genetic predictors. | 37774429 |
| Yang | 2024 | Biomarker or genetic predictors. | 39285345 |
| Kaya | 2024 | Biomarker or genetic predictors. | 39217284 |
| Liang | 2024 | Biomarker or genetic predictors. | 38625888 |
| Huang | 2024 | Biomarker or genetic predictors. | 39103437 |
| Xing | 2024 | Biomarker or genetic predictors. | 38460091 |
| Li Y | 2024 | Biomarker or genetic predictors. | 38963181 |
| Cheng | 2024 | Biomarker or genetic predictors. | 39165511 |
| Wang H | 2024 | Biomarker or genetic predictors. | 39462412 |
| Susarla | 2024 | Biomarker or genetic predictors. | 39394552 |
| Ding T | 2024 | Biomarker or genetic predictors. | 39393404 |
| Parsaei | 2024 | Biomarker or genetic predictors. | 39716122 |
| Hromadnikova | 2024 | Biomarker or genetic predictors. | 39296937 |
| Liang Q | 2024 | Biomarker or genetic predictors. | 39777224 |
| Zhou F | 2024 | Biomarker or genetic predictors. | 38975148 |
| Wu X | 2024 | Biomarker or genetic predictors. | 39619690 |
| Zhu B | 2024 | Biomarker or genetic predictors. | 39307357 |
| Mogos | 2024 | Biomarker or genetic predictors. | 39202579 |
| Shetty | 2024 | Biomarker or genetic predictors. | 39662422 |
| Naz | 2024 | Biomarker or genetic predictors. | 40018203 |
| Mathew | 2024 | Biomarker or genetic predictors. | 39545096 |
| Gu Y | 2025 | Biomarker or genetic predictors. | 39059385 |
| Tian | 2025 | Biomarker or genetic predictors. | 40221404 |
| Zhao M | 2025 | Biomarker or genetic predictors. | 39992231 |
| Yang Z | 2025 | Biomarker or genetic predictors. | 40035324 |
| Zheng C | 2025 | Biomarker or genetic predictors. | 40273822 |
| Borges | 2025 | Biomarker or genetic predictors. | 39694165 |
| Tang | 2025 | Biomarker or genetic predictors. | 39825726 |
| Zhang J | 2025 | Biomarker or genetic predictors. | 40175970 |
| Zhu T | 2025 | Biomarker or genetic predictors. | 40102801 |
| Lyu J | 2025 | Biomarker or genetic predictors. | 40274429 |
| Ni H | 2025 | Biomarker or genetic predictors. | 40314023 |
| Moon | 2025 | Biomarker or genetic predictors. | 40367989 |
| Bigdeli | 2025 | Biomarker or genetic predictors. | 39754258 |
| Liu K | 2025 | No access. | 39630585 |
| Shi Z | 2018 | No access. | No link. See Huang 2022 (36218132) references. |
| Xiao X | 2018 | No access. | No link. See Zhang 2022 (35294369) references. |
| Wang Y | 2019 | No access. | No link. See Huang 2022 (36218132) references. |
| Cen L | 2019 | No access. | No link. See Huang 2022 (36218132) references. |
| Chen M | 2019 | No access. | No link. See Huang 2022 (36218132) references. |
| Li K | 2019 | No access. | No link. See Huang 2022 (36218132) references. |
| Ma S | 2019 | No access. | No link. See Huang 2022 (36218132) references. |
| Miao Z | 2020 | No access. | No link. See Huang 2022 (36218132) references. |
| Ye J | 2020 | No access. | No link. See Huang 2022 (36218132) references. |

| | | | |
|---|---|---|---|
| Tan | 2020 | No access. | No link. See Zhang 2022 (35294369) references. |
| Cui | 2020 | No access. | No link. See Zhang 2022 (35294369) references. |
| Xu M | 2021 | No access. | No link. See Huang 2022 (36218132) references. |
| Yang T (2 models) | 2021 | No access. | No link. See Huang 2022 (36218132) references. |
| Deng F | 2021 | No access. | No link. See Huang 2022 (36218132) references. |
| Guo J | 2021 | No access. | No link. See Huang 2022 (36218132) references. |
| Meng Y | 2021 | No access. | No link. See Huang 2022 (36218132) references. |
| Zhang LQ (4 models) | 2021 | No access. | No link. See Huang 2022 (36218132) references. |
| Zhang XQ | 2021 | No access. | No link. See Huang 2022 (36218132) references. |
| Chen Q | 2022 | No access. | No link. See Huang 2022 (36218132) references. |
| Rao J | 2022 | No access. | No link. See Huang 2022 (36218132) references. |
| Huang Y | 2017 | No access. | No link. See Zhang 2022 (35294369) references. |
| Wu B | 2017 | No access. | No link. See Zhang 2022 (35294369) references. |

Additional file 3: Table S11: Excluded models and reasons (b) - pre-eclampsia (any onset)

| First Author | Publish Year | Reason for exclusion | Development paper link / PMID |
|---|---|---|---|
| Wright (a, maternal characteristic only) | 2012 | Outcome ineligible. | 22846473 |
| Wright (b, 3 models) | 2012 | Biomarker or genetic predictors.<br>Outcome ineligible. | 22846473 |
| Wright (2 models) | 2015 | Outcome ineligible. | 25724400 |
| Massé (4 models) | 1993 | Biomarker or genetic predictors. | 8372852 |
| Harrington | 1997 | Biomarker or genetic predictors. | 9197870 |
| Mello | 2002 | Biomarker or genetic predictors. | 12375550 |
| August | 2004 | Biomarker or genetic predictors. | 15547540 |
| Yu K | 2005 | Biomarker or genetic predictors. | 16098866 |
| Khaw | 2008 | Biomarker or genetic predictors. | 18190374 |
| Leal | 2009 | Biomarker or genetic predictors. | 19173240 |
| Phaloprakarn | 2009 | Biomarker or genetic predictors. | 19591558 |
| Poon (1) | 2009 | Biomarker or genetic predictors. | 19273739 |
| Farina | 2010 | Biomarker or genetic predictors. | 20934680 |
| Goetzinger | 2010 | Biomarker or genetic predictors. | 20936638 |
| Sekizawa | 2010 | Biomarker or genetic predictors. | 20121832 |
| North (a) | 2011 | Some predictors not available. | 21474517 |
| North (b) | 2011 | Biomarker or genetic predictors. | 21474517 |
| Odibo (1) (5 models) | 2011 | Biomarker or genetic predictors. | 21652068 |
| Odibo (2) (2 models) | 2011 | Biomarker or genetic predictors. | 21744367 |
| Myatt | 2012 | Biomarker or genetic predictors. | 22617589 |
| Ghojazadeh | 2013 | Biomarker or genetic predictors. | 24403715 |
| Kleinrouweler (6 models) | 2013 | Biomarker or genetic predictors. | 23417857 |
| Scazzocchio | 2013 | Biomarker or genetic predictors. | 23246313 |
| Gurgel Alves | 2014 | Biomarker or genetic predictors. | 24585555 |
| Kenny | 2014 | Biomarker or genetic predictors. | 25122928 |
| Skråstad | 2014 | Biomarker or genetic predictors. | 25146367 |
| Teixeira | 2014 | Biomarker or genetic predictors. | 24991215 |
| Garcés | 2015 | Biomarker or genetic predictors. | 26121675 |
| Han Q (a) | 2020 | Biomarker or genetic predictors. | 32706060 |
| Han Q (b) | 2020 | Machine learning models.<br>No reproducible information reported. | 32706060 |
| Yue CY | 2021 | Biomarker or genetic predictors. | 33060833 |
| Tarca (6 models) | 2022 | Biomarker or genetic predictors. | 34998477 |
| Chen (2 models) | 2023 | Biomarker or genetic predictors. | 37337643 |
| Chen J (a) | 2024 | No reproducible information reported. | 38568284 |
| Chen J (b, 2 models) | 2024 | Biomarker or genetic predictors. | 38568284 |
| Deeba | 2024 | Biomarker or genetic predictors. | 39549432 |
| Chen SM | 2025 | Biomarker or genetic predictors. | 40425699 |
| Ling Y | 2025 | Biomarker or genetic predictors. | 40181312 |
| Lu YJ | 2025 | Biomarker or genetic predictors. | 40980178 |
| Wang C | 2025 | Biomarker or genetic predictors. | 40450650 |
| Wu X | 2025 | Biomarker or genetic predictors. | 40385044 |
| Wu Y | 2025 | Biomarker or genetic predictors. | 40188283 |
| Xiao F | 2025 | Biomarker or genetic predictors. | 39926686 |
| Xu Q | 2025 | Biomarker or genetic predictors. | 40426100 |
| Kenny | 2010 | Biomarker or genetic predictors.<br>No model information. | 20837882 |

| | | | |
|---|---|---|---|
| Antonios (3 models) | 2013 | Biomarker or genetic predictors.<br>Less than 3 predictors (some models). | 23757401 |
| Pilalis | 2007 | Biomarker or genetic predictors.<br>No intercept and coefficient reported. | 17221926 |
| Thilaganathan (2 models) | 2010 | Biomarker or genetic predictors.<br>No intercept and coefficient reported. | 20502295 |
| Di Lorenzo | 2012 | Biomarker or genetic predictors.<br>No intercept and coefficient reported. | 22459245 |
| Khalil (4 models) | 2012 | Biomarker or genetic predictors.<br>No intercept and coefficient reported. | 22565361 |
| Cohen | 2014 | Biomarker or genetic predictors.<br>No intercept and coefficient reported. | 25129153 |
| Goetzinger | 2014 | Biomarker or genetic predictors.<br>No intercept and coefficient reported. | 24705967 |
| Moon | 2014 | Biomarker or genetic predictors.<br>No intercept and coefficient reported. | 25330843 |
| Mostafa Fouad Gomaa | 2015 | Biomarker or genetic predictors.<br>No intercept and coefficient reported. | 25881914 |
| Kumar | 2016 | Biomarker or genetic predictors.<br>No intercept and coefficient reported. | 27136371 |
| Diguisto | 2017 | Biomarker or genetic predictors.<br>No intercept and coefficient reported. | 27151901 |
| Kim | 2019 | Biomarker or genetic predictors.<br>No intercept and coefficient reported. | 31220966 |
| Liu Y | 2022 | Biomarker or genetic predictors.<br>No intercept and coefficient reported. | 35774506 |
| Ratnik (11 models) | 2022 | Biomarker or genetic predictors.<br>No intercept and coefficient reported. | 35966513 |
| Huang Z | 2024 | Biomarker or genetic predictors.<br>No intercept and coefficient reported. | 39148025 |
| Ma Y (4 models) | 2024 | Biomarker or genetic predictors.<br>No intercept and coefficient reported. | 38750217 |
| Qin H | 2024 | Biomarker or genetic predictors.<br>No intercept and coefficient reported. | 38941397 |
| Agarwal (18 models) | 2025 | Biomarker or genetic predictors.<br>No intercept and coefficient reported. | 41180159 |
| Han X | 2025 | Biomarker or genetic predictors.<br>No intercept and coefficient reported. | 39871211 |
| Suksai | 2025 | Biomarker or genetic predictors.<br>No intercept and coefficient reported. | 40035337 |
| Svirsky (4 models) | 2025 | Biomarker or genetic predictors.<br>No intercept and coefficient reported. | 40396999 |
| Wang X (5 models) | 2025 | Biomarker or genetic predictors.<br>No intercept and coefficient reported. | 39825806 |
| Wang Z (7 models) | 2025 | Biomarker or genetic predictors.<br>No intercept and coefficient reported. | 39815029 |
| Ye X | 2025 | Biomarker or genetic predictors.<br>No intercept and coefficient reported. | 40203595 |
| Zhang Han | 2025 | Biomarker or genetic predictors.<br>No intercept and coefficient reported. | 40837671 |
| Zhang Hui (6 models) | 2025 | Biomarker or genetic predictors.<br>No intercept and coefficient reported. | 41468471 |

| | | | |
|---|---|---|---|
| Zhang J | 2025 | Biomarker or genetic predictors.<br>No intercept and coefficient reported. | 40949738 |
| Akolekar | 2013 | Biomarker or genetic predictors.<br>Outcome ineligible. | 22906914 |
| Allen | 2017 | Less than 3 predictors. | 28714317 |
| Cheng Y (3 models) | 2018 | Less than 3 predictors. | 29032578 |
| Eberhard BW (2 models) | 2024 | Machine learning models and Cox models.<br>No reproducible information reported. | 39002866 |
| Nair (3 models) | 2018 | Machine learning models.<br>No reproducible information reported. | 29859381 |
| Sandström | 2019 | Machine learning models.<br>No reproducible information reported. | 31774875 |
| Sufriyana (6 models) | 2020 | Machine learning models.<br>No reproducible information reported. | 32283530 |
| Manoochehri (5 models) | 2021 | Machine learning models.<br>No reproducible information reported. | 34977453 |
| Ansbacher-Feldman (2 models) | 2022 | Machine learning models.<br>No reproducible information reported. | 36454636 |
| Bennett (16 models) | 2022 | Machine learning models.<br>No reproducible information reported. | 35385525 |
| Gómez-Jemes | 2022 | Machine learning models.<br>No reproducible information reported. | https://doi.org/10.3390/electronics11193240 |
| Li S (19 models) | 2022 | Machine learning models.<br>No reproducible information reported. | 35668134 |
| Melinte-Popescu (4 models) | 2023 | Machine learning models.<br>No reproducible information reported. | 36675347 |
| Torres-Torres (4 models) | 2023 | Machine learning models.<br>No reproducible information reported. | 37774112 |
| Wang H (5 models) | 2023 | Machine learning models.<br>No reproducible information reported. | 36849879 |
| Baetens (6 models) | 2024 | Machine learning models.<br>No reproducible information reported. | 39695764 |
| He A | 2024 | Machine learning models.<br>No reproducible information reported. | 38914704 |
| Jung YM (2 models) | 2024 | Machine learning models.<br>No reproducible information reported. | 39403751 |
| Kovacheva (12 models) | 2024 | Machine learning models.<br>No reproducible information reported. | 37901968 |
| Li T (25 models) | 2024 | Machine learning models.<br>No reproducible information reported. | 38919479 |
| Liu X | 2024 | Machine learning models.<br>No reproducible information reported. | 39707003 |
| Starodubtseva (2 models) | 2024 | Machine learning models.<br>No reproducible information reported. | 39408980 |
| Tiruneh (10 models) | 2024 | Machine learning models.<br>No reproducible information reported. | 39393122 |
| Wang (6 models) | 2024 | Machine learning models.<br>No reproducible information reported. | 38373676 |
| Wang L (10 models) | 2024 | Machine learning models.<br>No reproducible information reported. | 38326453 |
| Zhao Z (4 models) | 2024 | Machine learning models.<br>No reproducible information reported. | 39409013 |

| | | | |
|---|---|---|---|
| Zhou TF | 2024 | Machine learning models.<br>No reproducible information reported. | 38230614 |
| Adil M | 2025 | Machine learning models.<br>No reproducible information reported. | 39939524 |
| Chen J (113 models) | 2025 | Machine learning models.<br>No reproducible information reported. | 40552285 |
| Chen SC (127 models) | 2025 | Machine learning models.<br>No reproducible information reported. | 40312361 |
| Eberhard (96 models) | 2025 | Machine learning models.<br>No reproducible information reported. | 40493626 |
| Lin R | 2025 | Machine learning models.<br>No reproducible information reported. | 39916955 |
| Lv B | 2025 | Machine learning models.<br>No reproducible information reported. | 39889366 |
| Nakano (4 models) | 2025 | Machine learning models.<br>No reproducible information reported. | 40427999 |
| Yu T (6 models) | 2025 | Machine learning models.<br>No reproducible information reported. | 39853851 |
| Zhao Z (2 models) | 2025 | Machine learning models.<br>No reproducible information reported. | 40229739 |
| Li YX (4 models) | 2021 | Machine learning models.<br>No reproducible information reported.<br>Biomarker or genetic predictors. | 34739939 |
| Liu MY | 2022 | Machine learning models.<br>No reproducible information reported.<br>Biomarker or genetic predictors. | 36035487 |
| Wang Q (4 models) | 2022 | Machine learning models.<br>No reproducible information reported.<br>Retracted paper. | 35035520 |
| Boutin (2 models) | 2018 | No access. | 29079078 |
| Ghesquière L | 2024 | No access. | 38490251 |
| Yuan F | 2024 | No access. | 38965198 |
| Kawakita | 2025 | No access. | 39631775 |
| Long D | 2025 | No access. | 39467580 |
| Kern-Goldberger AR | 2025 | No access. | 40174872 |
| Praciano de Souza | 2018 | No access.<br>Not English. | 27300275 |
| Suksai | 2022 | No intercept reported. | 34151676 |
| Syngelaki | 2011 | Some predictors not available. | 22067258 |
| Audibert (4 models) | 2010 | No reproducible information reported. | 20691410 |
| Goetzinger (8 models) | 2013 | No reproducible information reported. | 23980220 |
| Austdal (4 models) | 2015 | No reproducible information reported. | 26370975 |
| Rocha | 2017 | No reproducible information reported. | 29153662 |
| Murtoniemi (a) | 2018 | No reproducible information reported. | 29970026 |
| Kaya Y (10 models) | 2024 | No reproducible information reported.<br>Machine learning models. | 39797241 |
| Li Q (2 models) | 2024 | No reproducible information reported. | 39544937 |
| Gonen (6 models) | 2008 | No reproducible information reported.<br>Biomarker or genetic predictors. | 19035985 |
| Yu J (7 models) | 2011 | No reproducible information reported.<br>Biomarker or genetic predictors. | 20737451 |

| | | | |
|---|---|---|---|
| Schneuer | 2012 | No reproducible information reported.<br>Biomarker or genetic predictors. | 22748852 |
| Zhou | 2012 | No reproducible information reported.<br>Biomarker or genetic predictors. | 22734455 |
| Diguisto (4 models) | 2013 | No reproducible information reported.<br>Biomarker or genetic predictors. | 23847040 |
| Schneuer (c, 2 models) | 2013 | No reproducible information reported.<br>Biomarker or genetic predictors. | 26103799 |
| Giguère | 2014 | No reproducible information reported.<br>Biomarker or genetic predictors. | 25175335 |
| Hannaford (15 models) | 2015 | No reproducible information reported.<br>Biomarker or genetic predictors. | 26014314 |
| Gabbay-Benziv | 2016 | No reproducible information reported.<br>Biomarker or genetic predictors. | 26448637 |
| Guy GP (6 models) | 2016 | No reproducible information reported.<br>Biomarker or genetic predictors. | 27619066 |
| Agarwal | 2017 | No reproducible information reported.<br>Biomarker or genetic predictors. | 28569565 |
| Asiltas (3 models) | 2018 | No reproducible information reported.<br>Biomarker or genetic predictors. | 29510888 |
| Murtoniemi (b, 2 models) | 2018 | No reproducible information reported.<br>Biomarker or genetic predictors. | 29970026 |
| Guo Z | 2020 | No reproducible information reported.<br>Biomarker or genetic predictors. | 32274292 |
| Wang W | 2020 | No reproducible information reported.<br>Biomarker or genetic predictors. | 32526695 |
| Chen G (4 models) | 2024 | No reproducible information reported.<br>Biomarker or genetic predictors. | 38926668 |
| Hromadnikova I | 2024 | No reproducible information reported.<br>Biomarker or genetic predictors. | 39296937 |
| Hodžić (3 models) | 2025 | No reproducible information reported.<br>Biomarker or genetic predictors. | 40129334 |
| Jørgensen (7 models) | 2025 | No reproducible information reported.<br>Biomarker or genetic predictors. | 39607297 |
| Spencer (4 models) | 2006 | No reproducible information reported.<br>Biomarker or genetic predictors.<br>Less than 3 predictors. | 16493628 |
| Spencer (1) (6 models) | 2007 | No reproducible information reported.<br>Biomarker or genetic predictors.<br>Less than 3 predictors. | 17278173 |
| Spencer (2) (4 models) | 2007 | No reproducible information reported.<br>Biomarker or genetic predictors.<br>Less than 3 predictors. | 17149788 |
| Spencer (2 models) | 2008 | No reproducible information reported.<br>Biomarker or genetic predictors.<br>Less than 3 predictors. | 18816493 |
| Spencer | 2005 | No reproducible information reported.<br>Biomarker or genetic predictors.<br>Multivariate Gaussian Model.<br>Less than 3 predictors. | 16086443 |

| | | | |
|---|---|---|---|
| Gallo | 2016 | No reproducible information reported.<br>Biomarker or genetic predictors.<br>Outcome ineligible. | 26627730 |
| O'Gorman | 2016 | No reproducible information reported.<br>Biomarker or genetic predictors.<br>Outcome ineligible. | 26297382 |
| Rolle V | 2024 | No reproducible information reported.<br>Biomarker or genetic predictors.<br>Outcome ineligible. | 39768791 |
| Schneuer (a) | 2013 | No reproducible information reported.<br>Less than 3 predictors. | 26103799 |
| Schneuer (b) | 2013 | No reproducible information reported.<br>Some predictors not available. | 26103799 |
| Gasse | 2019 | No reproducible information reported.<br>Machine learning models. | 29082781 |
| Marić (3 models) | 2020 | No reproducible information reported.<br>Machine learning models.<br>Biomarker or genetic predictors. | 33345966 |
| Ohseto (a, 4 models) | 2025 | Outcome ineligible.<br>No intercept reported. | 40258933 |
| Ohseto (b, 12 models) | 2025 | No reproducible information reported.<br>Biomarker or genetic predictors.<br>Outcome ineligible. | 40258933 |
| Ruan FY (3 models) | 2025 | No reproducible information reported.<br>Outcome ineligible. | 40016711 |
| Kindschuh WF (5 models) | 2024 | No reproducible information reported.<br>Preprint paper. | 39677801 |
| Mousavi SS | 2024 | Outcome ineligible. | 39677418 |
| Ma R | 2025 | Outcome ineligible. | 40821034 |
| Ballard (2 models) | 2024 | Outcome ineligible.<br>Some predictors not available.<br>No intercept reported. | 39141914 |
| Zain H | 2025 | Review article | 40511070 |
| Papageorghiou | 2005 | Some predictors not available. | 15924523 |
| De Paco (2 models) | 2008 | Some predictors not available. | 18238965 |
| Seed | 2011 | Some predictors not available. | 20795821 |
| Macdonald-Wallis (b, 6 models) | 2015 | Some predictors not available. | 26578347 |
| Al-Rubaie | 2020 | Some predictors not available. | 31906891 |
| Buciu VB | 2025 | Some predictors not available.<br>No intercept reported. | 40429393 |
| Capdeville (2 models) | 2025 | Some predictors not available.<br>No intercept reported. | 40584298 |

Additional file 3: Table S11: Excluded models and reasons (c) - pre-eclampsia (early onset)

| First Author | Publish Year | Reason for exclusion | Development paper link / PMID |
|---|---|---|---|
| Akolekar (1 model) | 2011 | No intercept reported. | 21210481 |
| Yu K | 2005 | Biomarker or genetic predictors. | 16098866 |
| Akolekar | 2008 | Biomarker or genetic predictors. | 18956425 |
| Onwudiwe | 2008 | Biomarker or genetic predictors. | 18991324 |
| Plasencia (2 models) | 2008 | Biomarker or genetic predictors. | 18634131 |
| Poon (2 models) | 2008 | Biomarker or genetic predictors. | 19090499 |
| Poon (1) | 2009 | Biomarker or genetic predictors. | 19273739 |
| Poon (2) | 2009 | Biomarker or genetic predictors. | 19827052 |
| Poon (3) | 2009 | Biomarker or genetic predictors. | 19242924 |
| Poon (4) (3 models) | 2009 | Biomarker or genetic predictors. | 19644947 |
| Foidart | 2010 | Biomarker or genetic predictors. | 20205159 |
| Poon (2) (4 models) | 2010 | Biomarker or genetic predictors. | 20108221 |
| Akolekar (1) | 2011 | Biomarker or genetic predictors. | 20205626 |
| Poon (2 models) | 2011 | Biomarker or genetic predictors. | 20232288 |
| Abdelaziz | 2012 | Biomarker or genetic predictors. | 22689569 |
| Bahado-Singh | 2012 | Biomarker or genetic predictors. | 22494326 |
| Caradeux | 2013 | Biomarker or genetic predictors. | 23584890 |
| Keikkala | 2013 | Biomarker or genetic predictors. | 23993394 |
| Parra-Cordero | 2013 | Biomarker or genetic predictors. | 22807133 |
| Crovetto (1) (b) | 2014 | Biomarker or genetic predictors. | 25346181 |
| Kenny | 2014 | Biomarker or genetic predictors. | 25122928 |
| Parra-Cordero | 2014 | Biomarker or genetic predictors. | 24903217 |
| Arakaki | 2015 | Biomarker or genetic predictors. | 25042564 |
| Tan MY (a) | 2018 | No reproducible information reported.<br>Outcome ineligible. | 29896812 |
| Tan MY (b, 15 models) | 2018 | No reproducible information reported.<br>Biomarker or genetic predictors.<br>Outcome ineligible. | 29896812 |
| Serra | 2020 | Biomarker or genetic predictors. | 31972161 |
| Xu SH | 2025 | Biomarker or genetic predictors. | 40211215 |
| Kuc | 2014 | Biomarker or genetic predictors.<br>Less than 3 predictors. | 24873829 |
| Di Lorenzo | 2012 | Biomarker or genetic predictors.<br>No intercept reported. | 22459245 |
| Akolekar | 2013 | Biomarker or genetic predictors.<br>Outcome ineligible. | 22906914 |
| Sandström | 2019 | Machine learning models.<br>No reproducible information reported. | 31774875 |
| Ansbacher-Feldman | 2022 | Machine learning models.<br>No reproducible information reported. | 36454636 |
| Melinte-Popescu (4 models) | 2023 | Machine learning models.<br>No reproducible information reported. | 36675347 |
| Saadaty (2 models) | 2023 | Machine learning models.<br>No reproducible information reported. | 37238928 |
| Torres-Torres (5 models) | 2023 | Machine learning models.<br>No reproducible information reported. | 37774112 |
| Xue (6 models) | 2023 | Machine learning models.<br>No reproducible information reported. | 37916364 |
| Khalil (2 models) | 2024 | Machine learning models.<br>No reproducible information reported. | 38432413 |

| | | | |
|---|---|---|---|
| Lin YC (12 models) | 2024 | Machine learning models.<br>No reproducible information reported. | 39716098 |
| Chen SC (127 models) | 2025 | Machine learning models.<br>No reproducible information reported. | 40312361 |
| Lv (5 models) | 2025 | Machine learning models.<br>No reproducible information reported. | 39889366 |
| Nakano (8 models) | 2025 | Machine learning models.<br>No reproducible information reported. | 40427999 |
| Nie (2 models) | 2025 | Machine learning models.<br>No reproducible information reported. | 40049809 |
| Marić (3 models) | 2020 | Machine learning models.<br>No reproducible information reported.<br>Biomarker or genetic predictors. | 33345966 |
| Li Z | 2022 | Machine learning models.<br>No reproducible information reported.<br>Outcome ineligible. | 36388117 |
| Audibert (4 models) | 2010 | No reproducible information reported. | 20691410 |
| Wortelboer | 2010 | No reproducible information reported. | 20840693 |
| Goetzinger (8 models) | 2013 | No reproducible information reported. | 23980220 |
| Murtoniemi (a) | 2018 | No reproducible information reported. | 29970026 |
| Murtoniemi (b, 2 models) | 2018 | No reproducible information reported.<br>Biomarker or genetic predictors. | 29970026 |
| Akolekar (1) | 2009 | No reproducible information reported.<br>Biomarker or genetic predictors. | 19777530 |
| Akolekar (2) | 2009 | No reproducible information reported.<br>Biomarker or genetic predictors. | 19412915 |
| Akolekar (3) | 2009 | No reproducible information reported.<br>Biomarker or genetic predictors. | 19776595 |
| Schneuer | 2012 | No reproducible information reported.<br>Biomarker or genetic predictors. | 22748852 |
| Crovetto (2) (a) | 2014 | No reproducible information reported.<br>Biomarker or genetic predictors. | 24714555 |
| Hannaford (15 models) | 2015 | No reproducible information reported.<br>Biomarker or genetic predictors. | 26014314 |
| Chang Y (3 models) | 2017 | No reproducible information reported.<br>Biomarker or genetic predictors. | 27678097 |
| Perales (5 models) | 2017 | No reproducible information reported.<br>Biomarker or genetic predictors. | 27883242 |
| Boutin (3 models) | 2019 | No reproducible information reported.<br>Biomarker or genetic predictors. | 30304731 |
| Zhao Q | 2025 | No reproducible information reported.<br>Biomarker or genetic predictors. | 40325391 |
| Nicolaides | 2006 | No reproducible information reported.<br>Biomarker or genetic predictors.<br>Less than 3 predictors. | 16374755 |
| Spencer (1) (6 models) | 2007 | No reproducible information reported.<br>Biomarker or genetic predictors.<br>Less than 3 predictors. | 17278173 |
| Spencer (2) (4 models) | 2007 | No reproducible information reported.<br>Biomarker or genetic predictors.<br>Less than 3 predictors. | 17149788 |

| | | | |
|---|---|---|---|
| He YJ (7 models) | 2025 | No reproducible information reported.<br>Less than 3 predictors (some models). | 39072715 |
| Poon (1) | 2010 | Some predictors not available. | 19516271 |
| Seed | 2011 | Some predictors not available. | 20795821 |

Additional file 3: Table S11: Excluded models and reasons (d) - pre-eclampsia (late onset)

| First Author | Publish Year | Reason for exclusion | Development paper link / PMID |
|---|---|---|---|
| Yu K | 2005 | Biomarker or genetic predictors. | 16098866 |
| Akolekar | 2008 | Biomarker or genetic predictors. | 18956425 |
| Onwudiwe | 2008 | Biomarker or genetic predictors. | 18991324 |
| Plasencia (2 models) | 2008 | Biomarker or genetic predictors. | 18634131 |
| Poon | 2008 | Biomarker or genetic predictors. | 19090499 |
| Poon (1) | 2009 | Biomarker or genetic predictors. | 19273739 |
| Poon (2) | 2009 | Biomarker or genetic predictors. | 19827052 |
| Poon (3) | 2009 | Biomarker or genetic predictors. | 19242924 |
| Poon (4) (3 models) | 2009 | Biomarker or genetic predictors. | 19644947 |
| Ashoor | 2010 | Biomarker or genetic predictors. | 20865794 |
| Poon (2) (2 models) | 2010 | Biomarker or genetic predictors. | 20108221 |
| Akolekar (1) | 2011 | Biomarker or genetic predictors. | 20205626 |
| Poon (2 models) | 2011 | Biomarker or genetic predictors. | 20232288 |
| Abdelaziz | 2012 | Biomarker or genetic predictors. | 22689569 |
| Parra-Cordero | 2013 | Biomarker or genetic predictors. | 22807133 |
| Crovetto (1) (b) | 2014 | Biomarker or genetic predictors. | 25346181 |
| Teixeira | 2014 | Biomarker or genetic predictors. | 24991215 |
| Kuc | 2014 | Biomarker or genetic predictors.<br>Less than 3 predictors. | 24873829 |
| Di Lorenzo | 2012 | Biomarker or genetic predictors.<br>No intercept reported. | 22459245 |
| Akolekar | 2013 | Biomarker or genetic predictors.<br>Outcome ineligible. | 22906914 |
| Jhee (5 models) | 2019 | Machine learning models.<br>No reproducible information reported.<br>Biomarker or genetic predictors. | 31442238 |
| Sandström | 2019 | Machine learning models.<br>No reproducible information reported. | 31774875 |
| Melinte-Popescu (4 models) | 2023 | Machine learning models.<br>No reproducible information reported. | 36675347 |
| Andresen (24 models) | 2024 | Machine learning models.<br>No reproducible information reported. | 39730472 |
| Chen SC (127 models) | 2025 | Machine learning models.<br>No reproducible information reported. | 40312361 |
| Nakano (6 models) | 2025 | Machine learning models.<br>No reproducible information reported. | 40427999 |
| Li Z | 2022 | Machine learning models.<br>No reproducible information reported.<br>Outcome ineligible (1 model). | 36388117 |
| Goetzinger (8 models) | 2013 | No intercept and coefficient reported. | 23980220 |
| Crovetto (2) (b) | 2014 | No intercept and coefficient reported.<br>Biomarker or genetic predictors. | 24714555 |
| Zhao Q | 2025 | No intercept and coefficient reported.<br>Biomarker or genetic predictors. | 40325391 |
| Audibert (4 models) | 2010 | No reproducible information reported. | 20691410 |
| Murtoniemi (a) | 2018 | No reproducible information reported. | 29970026 |
| Murtoniemi (b, 2 models) | 2018 | No reproducible information reported.<br>Biomarker or genetic predictors. | 29970026 |
| He YJ (7 models) | 2025 | No reproducible information reported. | 39072715 |

| | | | |
|---|---|---|---|
| Akolekar (2) | 2009 | No reproducible information reported.<br>Biomarker or genetic predictors. | 19412915 |
| Akolekar (3) | 2009 | No reproducible information reported.<br>Biomarker or genetic predictors. | 19776595 |
| Youssef | 2011 | No reproducible information reported.<br>Biomarker or genetic predictors. | 22034048 |
| Park | 2014 | No reproducible information reported.<br>Biomarker or genetic predictors. | 23919594 |
| Luo Q | 2017 | No reproducible information reported.<br>Biomarker or genetic predictors. | 27935854 |
| Boutin (3 models) | 2019 | No reproducible information reported.<br>Biomarker or genetic predictors. | 30304731 |
| Spencer (1) (6 models) | 2007 | No reproducible information reported.<br>Biomarker or genetic predictors.<br>Less than 3 predictors. | 17278173 |
| Spencer (2) (4 models) | 2007 | No reproducible information reported.<br>Biomarker or genetic predictors.<br>Less than 3 predictors. | 17149788 |

Additional file 3: Table S11: Excluded models and reasons (e) - pre-eclampsia (other)

| First Author | Publish Year | Reason for exclusion | Development paper link / PMID |
|---|---|---|---|
| Pihl (6 models) | 2019 | Less than 3 predictors. | 31622970 |
| Myers (b, 4 models) | 2013 | Biomarker or genetic predictors. | 23906160 |
| Kenny (2 models) | 2014 | Biomarker or genetic predictors. | 25122928 |
| Parra-Cordero (2 models) | 2014 | Biomarker or genetic predictors. | 24903217 |
| Skråstad | 2014 | Biomarker or genetic predictors. | 25146367 |
| Arakaki | 2017 | Biomarker or genetic predictors. | 27151901 |
| Sepúlveda-Martínez (a) | 2018 | Biomarker or genetic predictors. | 30230132 |
| Sepúlveda-Martínez (b) | 2018 | Less than 3 predictors. | 30230132 |
| Tan MY (a, 2 models) | 2018 | No reproducible information reported.<br>Outcome ineligible. | 29896812 |
| Tan MY (b, 30 models) | 2018 | No reproducible information reported.<br>Biomarker or genetic predictors.<br>Outcome ineligible. | 29896812 |
| Tarca (12 models) | 2022 | Biomarker or genetic predictors. | 34998477 |
| Murtoniemi (6 models) | 2018 | Less than 3 predictors.<br>No reproducible information reported. | 29970026 |
| Ansbacher-Feldman (3 models) | 2022 | Machine learning models.<br>No reproducible information reported. | 36454636 |
| Melinte-Popescu (8 models) | 2023 | Machine learning models.<br>No reproducible information reported. | 36675347 |
| Torres-Torres (5 models) | 2023 | Machine learning models.<br>No reproducible information reported. | 37774112 |
| Khalil (4 models) | 2024 | Machine learning models.<br>No reproducible information reported. | 38432413 |
| Li T (50 models) | 2024 | Machine learning models.<br>No reproducible information reported. | 38919479 |
| Lin YC (12 models) | 2024 | Machine learning models.<br>No reproducible information reported. | 39716098 |
| Zhang XY (2 models) | 2022 | Machine learning models.<br>No reproducible information reported. | https://doi.org/10.1016/j.medntd.2022.100158 |
| Giguère (2 models) | 2014 | No intercept and coefficient reported.<br>Biomarker or genetic predictors. | 25175335 |
| Wang X (5 models) | 2025 | No intercept and coefficient reported.<br>Biomarker or genetic predictors. | 39825806 |
| Akolekar (2 models) | 2011 | No intercept reported. | 21210481 |
| Valiño (2 models) | 2016 | No intercept reported.<br>Biomarker or genetic predictors. | 26094952/26224608 |
| Stamilio (2 models) | 2000 | No reproducible information reported. | 10739512 |
| Gasse (2 models) | 2019 | No reproducible information reported. | 29082781 |
| Chaiworapongsa (2 models) | 2013 | No reproducible information reported.<br>Biomarker or genetic predictors. | 23333542 |
| Skråstad (4 models) | 2014 | No reproducible information reported.<br>Biomarker or genetic predictors. | 25471057 |
| O'Gorman | 2016 | No reproducible information reported.<br>Biomarker or genetic predictors. | 26297382 |

Additional file 3: Table S11: Excluded models and reasons (f) - stillbirth

| First Author | Publish Year | Reason for exclusion | Development paper link / PMID |
|---|---|---|---|
| Smith (2) (2 models) | 2007 | Biomarker or genetic predictors. | 17197600 |
| Dugoff (2 models) | 2008 | Biomarker or genetic predictors. | 18771987 |
| Syngelaki | 2011 | Some predictors not available. | 22067258 |
| Payne | 2015 | Biomarker or genetic predictors. | 26597747 |
| Novillo-Del Álamo (a) | 2024 | Less than 3 predictors. | 39452566 |
| Novillo-Del Álamo (b, 3 models) | 2024 | Biomarker or genetic predictors. | 39452566 |
| Meng ZL | 2021 | Biomarker or genetic predictors.<br>Less than 3 predictors. | 34422871 |
| Koivu (12 models) | 2020 | Machine learning models.<br>No reproducible information reported.<br>Biomarker or genetic predictors. | 32226625 |
| Malacova (30 models) | 2020 | Machine learning models.<br>No reproducible information reported.<br>Biomarker or genetic predictors. | 32210300 |
| Cersonsky (20 models) | 2022 | Machine learning models.<br>No reproducible information reported.<br>Biomarker or genetic predictors. | 37315754 |
| Bosschieter | 2023 | Machine learning models.<br>No reproducible information reported.<br>Biomarker or genetic predictors. | 38273984 |
| Alzakari (14 models) | 2024 | Machine learning models.<br>No reproducible information reported.<br>Biomarker or genetic predictors. | 39424101 |
| Akolekar (1) (4 models) | 2016 | No intercept and coefficient reported.<br>Biomarker or genetic predictors. | 27854387 |
| Akolekar (2) (4 models) | 2016 | No intercept and coefficient reported.<br>Biomarker or genetic predictors. | 27854388 |
| Aupont (3 models) | 2016 | No intercept and coefficient reported.<br>Biomarker or genetic predictors. | 27854395 |
| Mastrodima | 2016 | No intercept and coefficient reported.<br>Biomarker or genetic predictors. | 27561595 |
| Åmark (4 models) | 2018 | No intercept and coefficient reported.<br>Biomarker or genetic predictors. | 30452441 |
| Hromadnikova (2 models) | 2023 | No intercept and coefficient reported.<br>Biomarker or genetic predictors. | 37373283 |
| Al-Fattah | 2024 | No intercept and coefficient reported.<br>Biomarker or genetic predictors. | 38961831 |
| Hromadnikova | 2024 | No intercept and coefficient reported.<br>Biomarker or genetic predictors. | 39296937 |
| Khalil (2 models) | 2015 | No intercept reported.<br>Biomarker or genetic predictors. | 26327300 |
| Familiari (4 models) | 2016 | No intercept reported.<br>Biomarker or genetic predictors. | 27588413 |
| Gunenc | 2025 | No intercept reported.<br>Biomarker or genetic predictors. | 40142283 |
| Torres-Torres | 2025 | No intercept reported.<br>Biomarker or genetic predictors. | 40247285 |
| Smith (1) (3 models) | 2007 | No intercept reported.<br>Biomarker or genetic predictors. | 17516962 |

| | | | |
|---|---|---|---|
| Akolekar | 2011 | No intercept reported.<br>Biomarker or genetic predictors. | 21210479 |
| Wu Jiayue | 2019 | No intercept reported.<br>Biomarker or genetic predictors. | 30755448 |
| Bahado-Singh | 2019 | No reproducible information reported.<br>Biomarker or genetic predictors. | 29712497 |
| Shukla (12 models) | 2020 | No reproducible information reported.<br>Some predictors not available. | 33206194 |
| Khatibi (24 models) | 2021 | No reproducible information reported.<br>Some predictors not available. | 33706701 |
| Lee SJ | 2023 | Outcome ineligible. | 37454734 |
| Kubahoniyesu | 2024 | Outcome ineligible. | 39637200 |
| Ngusie | 2024 | Outcome ineligible. | 39075434 |
| Giorgione | 2025 | Outcome ineligible. | 40051381 |
| Gothwal | 2025 | Outcome ineligible. | 41169841 |
| Prasad | 2025 | Outcome ineligible. | 39788362 |
| Yehuala | 2025 | Outcome ineligible. | 39995747 |
| Zhan Y | 2025 | Outcome ineligible. | 39934709 |
| Romero-Gutiérrez | 2005 | Some predictors not available. | 15603559 |
| Kayode (2 models) | 2016 | Some predictors not available. | 27649795 |
| Kumar | 2022 | Some predictors not available. | 35928077 |
| Goyal | 2015 | Biomarker or genetic predictors.<br>Outcome ineligible. | 26102375 |

Additional file 3: Table S11: Excluded models and reasons (g) - small-for-gestational-age

| First Author | Publish Year | Reason for exclusion | Development paper link / PMID |
|---|---|---|---|
| Doherty (2 models) | 2002 | Biomarker or genetic predictors. | 12467852 |
| Onwudiwe | 2008 | Biomarker or genetic predictors. | 18991324 |
| Karagiannis (3 models) | 2011 | Biomarker or genetic predictors. | 21079385 |
| Poon (2 models) | 2011 | Biomarker or genetic predictors. | 20799245 |
| Sulek (3 models) | 2014 | Biomarker or genetic predictors. | 25057319 |
| Sharp | 2019 | Biomarker or genetic predictors. | 31499415 |
| Xu C (a) | 2021 | Biomarker or genetic predictors. | 32364311 |
| Xu C (b) | 2021 | Machine learning models.<br>No reproducible information reported.<br>Biomarker or genetic predictors. | 32364311 |
| Iwama | 2022 | Biomarker or genetic predictors. | 35618764 |
| Adjahou | 2024 | Biomarker or genetic predictors. | 39586023 |
| Deeba | 2024 | Biomarker or genetic predictors. | 39549432 |
| Schaak (2 models) | 2024 | Biomarker or genetic predictors. | 39002400 |
| Hernández-Castro | 2021 | Biomarker or genetic predictors.<br>Less than 3 predictors. | 34008784 |
| Kumar (2 models) | 2024 | Biomarker or genetic predictors.<br>Less than 3 predictors. | 39013215 |
| Ryu | 2019 | Biomarker or genetic predictors.<br>Outcome ineligible. | 31045822 |
| Hassen | 2020 | Biomarker or genetic predictors.<br>Outcome ineligible. | 32456155 |
| Allotey | 2024 | Duplicated model. | 39184566 |
| Anggraini (4 models) | 2018 | Less than 3 predictors.<br>Biomarker or genetic predictors. | 30400880 |
| Street | 2008 | Machine learning models.<br>Biomarker or genetic predictors. | 18559101 |
| Kuhle (24 models) | 2018 | Machine learning models.<br>No reproducible information reported. | 30111303 |
| Saw (52 models) | 2021 | Machine learning models.<br>No reproducible information reported. | 33462877 |
| Chen XY | 2024 | Machine learning models.<br>No reproducible information reported. | 39642647 |
| Chen ZH | 2024 | Machine learning models.<br>No reproducible information reported. | 39365033 |
| Lee | 2024 | Machine learning models.<br>No reproducible information reported. | 39598319 |
| Mohana | 2024 | Machine learning models.<br>No reproducible information reported. | 39723659 |
| Ulusoy (3 models) | 2024 | Machine learning models.<br>No reproducible information reported. | 39422068 |
| Andresen (12 models) | 2025 | Machine learning models.<br>No reproducible information reported. | 40323295 |
| Dong (7 models) | 2025 | Machine learning models.<br>No reproducible information reported. | 40600032 |
| Hua Q (14 models) | 2025 | Machine learning models.<br>No reproducible information reported. | 40424611 |
| Lopian (6 models) | 2025 | Machine learning models.<br>No reproducible information reported. | 39864484 |

| | | | |
|---|---|---|---|
| Mikołaj (3 models) | 2025 | Machine learning models.<br>No reproducible information reported. | 40437236 |
| Pavanya (6 models) | 2025 | Machine learning models.<br>No reproducible information reported. | 40963798 |
| Spairani | 2025 | Machine learning models.<br>No reproducible information reported. | 40184939 |
| Sufriyana | 2025 | Machine learning models.<br>No reproducible information reported. | 40065124 |
| Yu QY (84 models) | 2025 | Machine learning models.<br>No reproducible information reported. | 41602314 |
| Magenes (3 models) | 2004 | Machine learning models.<br>No reproducible information reported.<br>Biomarker or genetic predictors. | 17271713 |
| Lunghi (4 models) | 2005 | Machine learning models.<br>No reproducible information reported.<br>Biomarker or genetic predictors. | https://ieeexplore.ieee.org/abstract/document/1588083 |
| Horgan | 2014 | Machine learning models.<br>No reproducible information reported.<br>Biomarker or genetic predictors. | 21671558 |
| Magenes (15 models) | 2016 | Machine learning models.<br>No reproducible information reported.<br>Biomarker or genetic predictors. | 28268473 |
| Dahdouh (12 models) | 2018 | Machine learning models.<br>No reproducible information reported.<br>Biomarker or genetic predictors. | 28734056 |
| Bahado-Singh (7 models) | 2019 | Machine learning models.<br>No reproducible information reported.<br>Biomarker or genetic predictors. | 30998683 |
| Signorini (15 models) | 2020 | Machine learning models.<br>No reproducible information reported.<br>Biomarker or genetic predictors. | 31678794 |
| Sufriyana (7 models) | 2020 | Machine learning models.<br>No reproducible information reported.<br>Biomarker or genetic predictors. | 32348266 |
| Van | 2020 | Machine learning models.<br>No reproducible information reported.<br>Biomarker or genetic predictors. | 33257111 |
| Crockart (4 models) | 2021 | Machine learning models.<br>No reproducible information reported.<br>Biomarker or genetic predictors. | 34007875 |
| Pini (2 models) | 2021 | Machine learning models.<br>No reproducible information reported.<br>Biomarker or genetic predictors. | 33889841 |
| Aslam (8 models) | 2022 | Machine learning models.<br>No reproducible information reported.<br>Biomarker or genetic predictors. | https://doi.org/10.3390/electronics11040593 |
| Deval | 2022 | Machine learning models.<br>No reproducible information reported.<br>Biomarker or genetic predictors. | 35941474 |
| Gómez-Jemes | 2022 | Machine learning models.<br>No reproducible information reported.<br>Biomarker or genetic predictors. | https://doi.org/10.3390/electronics11193240 |

| | | | |
|---|---|---|---|
| Weiner | 1985 | No access. | 3889747 |
| Snidvongs | 1989 | No access. | 2794823 |
| de Caunes | 1990 | No access. | 1977639 |
| Foltran | 2011 | No access. | 21880216 |
| Boucoiran | 2013 | No access. | 23283804 |
| Morillon | 2018 | No access. | No link. See Leite 2019 (31401613) references. |
| Poon | 2011 | No intercept reported.<br>No reproducible information reported. | 20799245 |
| Griffin (7 models) | 2018 | No intercept reported.<br>Biomarker or genetic predictors. | 28401605 |
| Tang | 2025 | No intercept reported.<br>Biomarker or genetic predictors. | 40771397 |
| Yang YX | 2025 | No intercept reported.<br>Biomarker or genetic predictors. | 40909444 |
| You SY | 2025 | No intercept reported.<br>Biomarker or genetic predictors. | 41098503 |
| Schwartz (2 models) | 2022 | No reproducible information reported. | 34553780 |
| Huang HF | 2025 | No reproducible information reported.<br>Less than 3 predictors. | 40927559 |
| Schneuer (6 models) | 2014 | No reproducible information reported.<br>Biomarker or genetic predictors. | 24215861 |
| Hromadnikova (6 models) | 2024 | No reproducible information reported.<br>Biomarker or genetic predictors. | 39296937 |
| Zhang B (2 models) | 2024 | No reproducible information reported.<br>Biomarker or genetic predictors. | 39497103 |
| Aktemur | 2025 | No reproducible information reported.<br>Biomarker or genetic predictors. | 40174268 |
| Lopian (14 models) | 2025 | No reproducible information reported.<br>Biomarker or genetic predictors. | 40286315 |
| Luo | 2025 | No reproducible information reported.<br>Biomarker or genetic predictors. | 39934694 |
| Wang JD | 2025 | No reproducible information reported.<br>Biomarker or genetic predictors. | 40114121 |
| Zhang B (2 models) | 2025 | No reproducible information reported.<br>Biomarker or genetic predictors. | 39849974 |
| Zhu HM (3 models) | 2025 | No reproducible information reported.<br>Biomarker or genetic predictors. | 40211115 |
| Bai XY (11 models) | 2025 | No reproducible information reported.<br>Biomarker or genetic predictors.<br>Less than 3 predictors. | 40349027 |
| Bachmann | 2003 | Outcome ineligible. | 12704746 |
| Buscema (8 models) | 2015 | Outcome ineligible. | 26158499 |
| Papastefanou (3 models) | 2020 | Outcome ineligible. | 32573831 |
| Nowacka | 2021 | Outcome ineligible. | 34501234 |
| Papastefanou | 2021 | Outcome ineligible. | 32936500 |
| Papastefanou (1) | 2021 | Outcome ineligible. | 33901487 |
| Papastefanou (2) | 2021 | Outcome ineligible. | 33464642 |
| Nicolaides | 2022 | Outcome ineligible. | 34919332 |
| Nowacka | 2022 | Outcome ineligible. | 34214232 |
| Papastefanou | 2022 | Outcome ineligible. | 36056735 |
| Bartnicki | 1996 | Biomarker or genetic predictors.<br>Outcome ineligible. | 8633678 |

| | | | |
|---|---|---|---|
| Liu CM (2 models) | 2008 | Some predictors not available. | 18937700 |
| Cohen | 2014 | Biomarker or genetic predictors.<br>Outcome ineligible. | 25129153 |
| Singh | 2014 | Biomarker or genetic predictors.<br>Outcome ineligible. | 23949869 |
| Goyal | 2015 | Biomarker or genetic predictors.<br>Outcome ineligible. | 26102375 |
| Sinding | 2017 | Biomarker or genetic predictors.<br>Outcome ineligible. | 28012454 |
| Ngusie (10 models) | 2024 | Biomarker or genetic predictors.<br>Outcome ineligible. | 39075434 |
| Wu WT | 2022 | Outcome ineligible.<br>Some predictors not available. | 34304668 |
| Reeb | 1987 | Outcome ineligible.<br>Some predictors not available. | 3324359 |
| Zahirzada (10 models) | 2021 | Outcome ineligible.<br>Some predictors not available. | https://ieeexplore.ieee.org/document/9415792 |
| Seed (2 models) | 2011 | Some predictors not available. | 20795821 |
| Syngelaki | 2011 | Some predictors not available. | 22067258 |
| Lesmes | 2015 | Some predictors not available. | 25704207 |
| Allotey (2 models) | 2024 | Some predictors not available. | 39252507 |
| Mamelle | 2001 | Variable printing error.<br>Variable definitions unclear. | 11641551 |
