## Supplementary material for "Systematic Review and External Validation of Clinical Prediction Models for Adverse Pregnancy Outcomes Using Routinely Collected Pre-Conception and Early Pregnancy Data": PRISMA checklist

### PRISMA 2020 Checklist

| Section and Topic | Item # | Checklist item | Location where item is reported |
|---|---|---|---|
| **TITLE** | | | |
| Title | 1 | Identify the report as a systematic review. | Title, P1, L1–3 |
| **ABSTRACT** | | | |
| Abstract | 2 | See the PRISMA 2020 for Abstracts checklist. | Abstract, P2–3, L24–51 |
| **INTRODUCTION** | | | |
| Rationale | 3 | Describe the rationale for the review in the context of existing knowledge. | Background, P4–5, L65–98 |
| Objectives | 4 | Provide an explicit statement of the objective(s) or question(s) the review addresses. | Background, P5, L99–106<br>Abstract (Background), P2, L27–31 |
| **METHODS** | | | |
| Eligibility criteria | 5 | Specify the inclusion and exclusion criteria for the review and how studies were grouped for the syntheses. | Methods (Literature search and selection), P6–7, L120–132, L139–154<br>Methods (Data Source and Study cohort), P8, L165–166<br>Table S1 |
| Information sources | 6 | Specify all databases, registers, websites, organisations, reference lists and other sources searched or consulted to identify studies. Specify the date when each source was last searched or consulted. | Methods (Literature search and selection), P6–7, L120–122, L133–138<br>Results (Eligible Models for External Validation), P12, L255–257<br>Table S1 |
| Search strategy | 7 | Present the full search strategies for all databases, registers and websites, including any filters and limits used. | Methods (Literature search and selection), P6–7, L120–122, L132, L136–138<br>Table S1 |
| Selection process | 8 | Specify the methods used to decide whether a study met the inclusion criteria of the review, including how many reviewers screened each record and each report retrieved, whether they worked independently, and if applicable, details of automation tools used in the process. | Methods (Literature search and selection), P7, L133–139<br>Declarations (Authors' contributions), P23, L498–500<br>Figure 1, P30, L653–656 |
| Data collection process | 9 | Specify the methods used to collect data from reports, including how many reviewers collected data from each report, whether they worked independently, any processes for obtaining or confirming data from study investigators, and if applicable, details of automation tools used in the process. | Methods (Literature search and selection), P7, L134–154<br>Declarations (Authors' contributions), P23, L498–500 |
| Data items | 10a | List and define all outcomes for which data were sought. Specify whether all results that were compatible with each outcome domain in each study were sought (e.g. for all measures, time points, analyses), and if not, the methods used to decide which results to collect. | Methods (Literature search and selection), P6–7, L120–132, L139–144<br>Methods (Definition of outcomes and predictors), P8–9, L169–182<br>Results (Eligible Models for External Validation), P12, L263–269 |

### PRISMA 2020 Checklist

| Section and Topic | Item # | Checklist item | Location where item is reported |
|---|---|---|---|
| | 10b | List and define all other variables for which data were sought (e.g. participant and intervention characteristics, funding sources). Describe any assumptions made about any missing or unclear information. | Methods (Literature search and selection), P6, L123–125<br>Methods (Definition of outcomes and predictors), P8–9, L169–174, L182–190<br>Table S2<br>Table S3 |
| Study risk of bias assessment | 11 | Specify the methods used to assess risk of bias in the included studies, including details of the tool(s) used, how many reviewers assessed each study and whether they worked independently, and if applicable, details of automation tools used in the process. | Not applicable |
| Effect measures | 12 | Specify for each outcome the effect measure(s) (e.g. risk ratio, mean difference) used in the synthesis or presentation of results. | Methods (Validation of existing models), P10–11, L213–231 |
| Synthesis methods | 13a | Describe the processes used to decide which studies were eligible for each synthesis (e.g. tabulating the study intervention characteristics and comparing against the planned groups for each synthesis (item #5)). | Methods (Literature search and selection), P6–7, L120–132, L139–154<br>Methods (Data Source and Study cohort), P8, L165–166<br>Methods (Validation of existing models), P10, L213–218<br>Table 1, P27, L644<br>Table S2 |
| | 13b | Describe any methods required to prepare the data for presentation or synthesis, such as handling of missing summary statistics, or data conversions. | Methods (Definition of outcomes and predictors), P8–9, L169–190<br>Methods (Missing data and multiple imputation), P9–10, L203–211<br>Methods (Validation of existing models), P10, L213–218 |
| | 13c | Describe any methods used to tabulate or visually display results of individual studies and syntheses. | Methods (Validation of existing models), P10–11, L219–231<br>Results (External Validation of Models), P14–15, L307–335<br>Table 1, P27, L644<br>Table 2, P28, L647<br>Table 3, P29, L650<br>Figure 2, P30, L657–664<br>Tables S2–S10<br>Figures S2–S3 |
| | 13d | Describe any methods used to synthesize results and provide a rationale for the choice(s). If meta-analysis was performed, describe the model(s), method(s) to identify the presence and extent of statistical heterogeneity, and software package(s) used. | Methods (Validation of existing models), P10–11, L213–231 (performance metrics pooled across 20 imputations using Rubin's rules; no meta-analysis performed) |

### PRISMA 2020 Checklist

| Section and Topic | Item # | Checklist item | Location where item is reported |
|---|---|---|---|
| | 13e | Describe any methods used to explore possible causes of heterogeneity among study results (e.g. subgroup analysis, meta-regression). | Methods (Data Source and Study cohort), P8, L165–166<br>Methods (Sensitivity Analysis), P11, L233–242<br>Discussion (Main findings and Interpretation), P16, L352–358<br>Discussion (Strengths and Limitations), P16–17, L360–372 |
| | 13f | Describe any sensitivity analyses conducted to assess robustness of the synthesized results. | Methods (Sensitivity Analysis), P11, L233–242 |
| Reporting bias assessment | 14 | Describe any methods used to assess risk of bias due to missing results in a synthesis (arising from reporting biases). | Not applicable |
| Certainty assessment | 15 | Describe any methods used to assess certainty (or confidence) in the body of evidence for an outcome. | Not applicable |
| **RESULTS** | | | |
| Study selection | 16a | Describe the results of the search and selection process, from the number of records identified in the search to the number of studies included in the review, ideally using a flow diagram. | Results (Eligible Models for External Validation), P12, L252–260<br>Figure 1, P30, L653–656 |
| | 16b | Cite studies that might appear to meet the inclusion criteria, but which were excluded, and explain why they were excluded. | Results (Eligible Models for External Validation), P12, L260–262<br>Additional file 3: Table S11 |
| Study characteristics | 17 | Cite each included study and present its characteristics. | Results (Eligible Models for External Validation), P12–13, L263–281<br>Table 1, P27, L644<br>Table 2, P28, L647<br>Results (Study Cohort Characteristics), P13–14, L294–305<br>Table 3, P29, L650<br>Tables S2–S4 |
| Risk of bias in studies | 18 | Present assessments of risk of bias for each included study. | Not applicable |
| Results of individual studies | 19 | For all outcomes, present, for each study: (a) summary statistics for each group (where appropriate) and (b) an effect estimates and its precision (e.g. confidence/credible interval), ideally using structured tables or plots. | Results (External Validation of Models), P14–15, L307–335<br>Table 2, P28, L647<br>Table 3, P29, L650<br>Table S5<br>Figure S2 |

### PRISMA 2020 Checklist

| Section and Topic | Item # | Checklist item | Location where item is reported |
|---|---|---|---|
| Results of syntheses | 20a | For each synthesis, briefly summarise the characteristics and risk of bias among contributing studies. | Results (Eligible Models for External Validation), P12–13, L263–281<br>Table 1, P27, L644<br>Results (Study Cohort Characteristics), P13–14, L294–305<br>Table 3, P29, L650<br>Tables S2–S4 |
| | 20b | Present results of all statistical syntheses conducted. If meta-analysis was done, present for each the summary estimate and its precision (e.g. confidence/credible interval) and measures of statistical heterogeneity. If comparing groups, describe the direction of the effect. | Results (External Validation of Models), P14–15, L307–335<br>Table 2, P28, L647<br>Figure 2, P30, L657–664<br>Table S5<br>Figure S2 |
| | 20c | Present results of all investigations of possible causes of heterogeneity among study results. | Results (Eligible Models for External Validation), P12–13, L263–281<br>Results (External Validation of Models), P14–15, L307–335<br>Results (Sensitivity analysis), P15, L337–344<br>Tables 1–2, P27–28, L644, L647<br>Figure 2, P30, L657–664<br>Tables S2, S5–S10<br>Figure S3 |
| | 20d | Present results of all sensitivity analyses conducted to assess the robustness of the synthesized results. | Results (Sensitivity analysis), P15, L337–344<br>Tables S6–S10<br>Figure S3 |
| Reporting biases | 21 | Present assessments of risk of bias due to missing results (arising from reporting biases) for each synthesis assessed. | Not applicable |
| Certainty of evidence | 22 | Present assessments of certainty (or confidence) in the body of evidence for each outcome assessed. | Not applicable |
| **DISCUSSION** | | | |
| Discussion | 23a | Provide a general interpretation of the results in the context of other evidence. | Discussion (Main findings and Interpretation), P16, L348–358<br>Conclusions, P19, L415–419 |
| | 23b | Discuss any limitations of the evidence included in the review. | Discussion (Main findings and Interpretation), P16, L352–358<br>Discussion (Implications for Clinical Practice and Future Research), P17, L380–386 |

### PRISMA 2020 Checklist

| Section and Topic | Item # | Checklist item | Location where item is reported |
|---|---|---|---|
| | 23c | Discuss any limitations of the review processes used. | Discussion (Strengths and Limitations), P16–17, L360–372 |
| | 23d | Discuss implications of the results for practice, policy, and future research. | Discussion (Implications for Clinical Practice and Future Research), P17–18, L374–402<br>Conclusions, P19, L415–419 |
| **OTHER INFORMATION** | | | |
| Registration and protocol | 24a | Provide registration information for the review, including register name and registration number, or state that the review was not registered. | Methods, P6, L111–118 (systematic review not registered) |
| | 24b | Indicate where the review protocol can be accessed, or state that a protocol was not prepared. | Methods, P6, L116–118 (CPRD ISAC protocol number 22_002169)<br>Declarations (Ethics approval and consent to participate), P22, L474–476<br>Declarations (Acknowledgements), P23, L509–510 |
| | 24c | Describe and explain any amendments to information provided at registration or in the protocol. | Not applicable |
| Support | 25 | Describe sources of financial or non-financial support for the review, and the role of the funders or sponsors in the review. | Declarations (Funding), P23, L492–494<br>Declarations (Acknowledgements), P23, L502–511 |
| Competing interests | 26 | Declare any competing interests of review authors. | Declarations (Competing interests), P22, L490–491 |
| Availability of data, code and other materials | 27 | Report which of the following are publicly available and where they can be found: template data collection forms; data extracted from included studies; data used for all analyses; analytic code; any other materials used in the review. | Declarations (Availability of data and materials), P22, L480–489<br>Methods (Literature search and selection), P6, L132<br>Results (Eligible Models for External Validation), P12, L260–262<br>Table S1<br>Tables S2, S5, S10<br>Additional file 3: Table S11 |

*From:* Page MJ, McKenzie JE, Bossuyt PM, Boutron I, Hoffmann TC, Mulrow CD, et al. The PRISMA 2020 statement: an updated guideline for reporting systematic reviews. BMJ 2021;372:n71. doi: 10.1136/bmj.n71
